# Interferon-driven T-cell activation and a senescence-associated signature characterize clinically active adult localized scleroderma

**DOI:** 10.64898/2026.11.23.26368779

**Authors:** Lauren Khoury, Adrien Osakwe, Elvis Martinez-Jaramillo, Don Vu, Ajay Rajaram, Abdulmalik Alqahtani, May Chergui, Mohammed Osman, Philippe Lefrançois, Liqin Xu, Yue Li, Elena Netchiporouk

## Abstract

**Background:** Localized scleroderma (LS, morphea) is an immune-mediated fibrosing skin disease whose molecular basis in adults, and its variation with clinical activity and histological stage, are poorly defined.

**Objectives:** To characterize transcriptional, cellular and senescence-associated programs in adult LS by clinical activity, histological stage and subtype, and to compare them with those of other inflammatory skin diseases.

**Methods:** We performed bulk RNA sequencing of archival lesional skin from 60 adults with LS and 22 healthy controls (HC), with enrichment analyses, cell-type deconvolution, immune-module scoring, cross-disease comparison and p16^INK4a^ immunohistochemistry.

**Results:** LS lesions showed 704 differentially expressed genes versus HC, enriched for interferon signaling, antigen presentation and T-helper differentiation pathways; changes were extensive in clinically active (n = 49) and histologically inflammatory (n = 21) lesions but limited in fibrotic lesions (n = 33). Deconvolution inferred expansion of fibroblasts, macrophages and regulatory, cytotoxic and helper T cells, with co-expression of effector and checkpoint genes in T-cell profiles (*IFNG*, *GZMK*, *LAG3*, *TIGIT*). Th1 and Th17 activity was higher in inflammatory than in fibrotic lesions, whereas Th2 activity did not differ between them. A senescence-associated gene set was enriched in LS (normalized enrichment score 1.64), including fibrotic lesions (nominal P <0.05), and dermal p16INK4a-positive cell density was higher than in HC (median 113.9 vs 36.0 cells/mm²; P = 0.003). LS signatures most closely resembled those of cutaneous lupus erythematosus and lichen planus.

**Conclusions:** Adult LS shows interferon-associated T-cell activation, most pronounced in clinically active and histologically inflammatory disease, and a senescence-associated signature. Type-2 and senescence-associated programs persist in fibrotic lesions. These findings support Janus kinase–signal transducer and activator of transcription (JAK–STAT) inhibition in active disease and trials of senescence-modulating therapies across disease stages.

**Plain language summary:** *What is happening in the skin of adults with morphea?:* Morphea, also called localized scleroderma, causes patches of skin to become inflamed and then hard and tight. It can leave lasting marks and limit movement. Most research on how morphea develops has been done in children, so less is known about adults, and treatment for adults is often chosen by trial and error. We studied skin samples from 60 adults with morphea and 22 people without skin disease. We measured which genes were switched on in the skin and used computer methods to estimate which types of cells were present. We also stained the samples for a marker of aged cells that have stopped dividing, called senescent cells. Skin from people with active, inflamed morphea showed strong signals from the immune system, especially from T cells, a type of white blood cell. Many of these signals were driven by interferon, a chemical messenger of inflammation. A second immune pathway that is linked to scarring, called Th2, was switched on in both inflamed and hardened skin. Morphea skin also showed signs of senescent cells, even in hardened areas. Our findings suggest that adults with morphea could be treated according to how active their disease is. Medicines that block interferon signals, called JAK inhibitors, may help people with active disease. Medicines that may reduce the harm caused by senescent cells, such as GLP-1 medicines used for diabetes and weight loss, deserve testing in clinical trials.

## Introduction

Targeted therapies have transformed the treatment of psoriasis and atopic dermatitis thanks to a clear understanding of their dominant immune axes^1^. Morphea (localized scleroderma, LS) has not yet benefited from this shift, although it causes lasting functional and cosmetic morbidity^2,3^ and affects approximately 3 per 100,000 person-years^4^. In adults, treatment remains largely empirical rather than guided by disease biology^5^.

Most mechanistic data come from children: active pediatric lesions show interferon-γ and CXCL9/10/11 chemokine signatures, and single-cell studies identify inflammatory, interferon-responsive fibroblasts^2,6–8^. Data in adults are fewer and come from small studies. In a microarray study of 25 morphea lesions, the transcriptome was dominated by interferon-γ/Th1 signaling, with few fibrosis pathways^9^. Single-cell studies of 13 adults found keratinocyte– and macrophage-centered programs that appeared less inflammatory than those of children^10^, and cytotoxic tissue-resident memory T cells that injure the endothelium^11^. None of these studies examined clinical activity, histological stage and clinical subtype together, so it remains unclear whether adult LS is less inflammatory than pediatric disease or whether earlier findings reflect the activity and stage of the lesions sampled. Senescent cells accumulate in systemic sclerosis skin^12^, and senescent immune cells can amplify inflammation in autoimmunity^13^, but senescence has not been examined at both the transcript and protein level in adult LS. These gaps matter clinically: adult LS is still treated empirically, and the immune pathways that would justify targeted therapies, such as Janus kinase (JAK) inhibitors, have not been mapped to disease activity in adults.

We therefore profiled lesional skin from 60 adults with LS by bulk RNA sequencing (RNA-seq), cell-type deconvolution and p16^INK4a^ immunohistochemistry (**Figure 1**). We hypothesized that type-1/type-17 T-cell, interferon and senescence-associated programs would be most pronounced in clinically active, histologically inflammatory lesions and would point to actionable therapeutic targets. To our knowledge, this is the largest transcriptomic study of adult LS to date and the first to examine clinical activity, histological stage and clinical subtype together.

**Figure 1.**
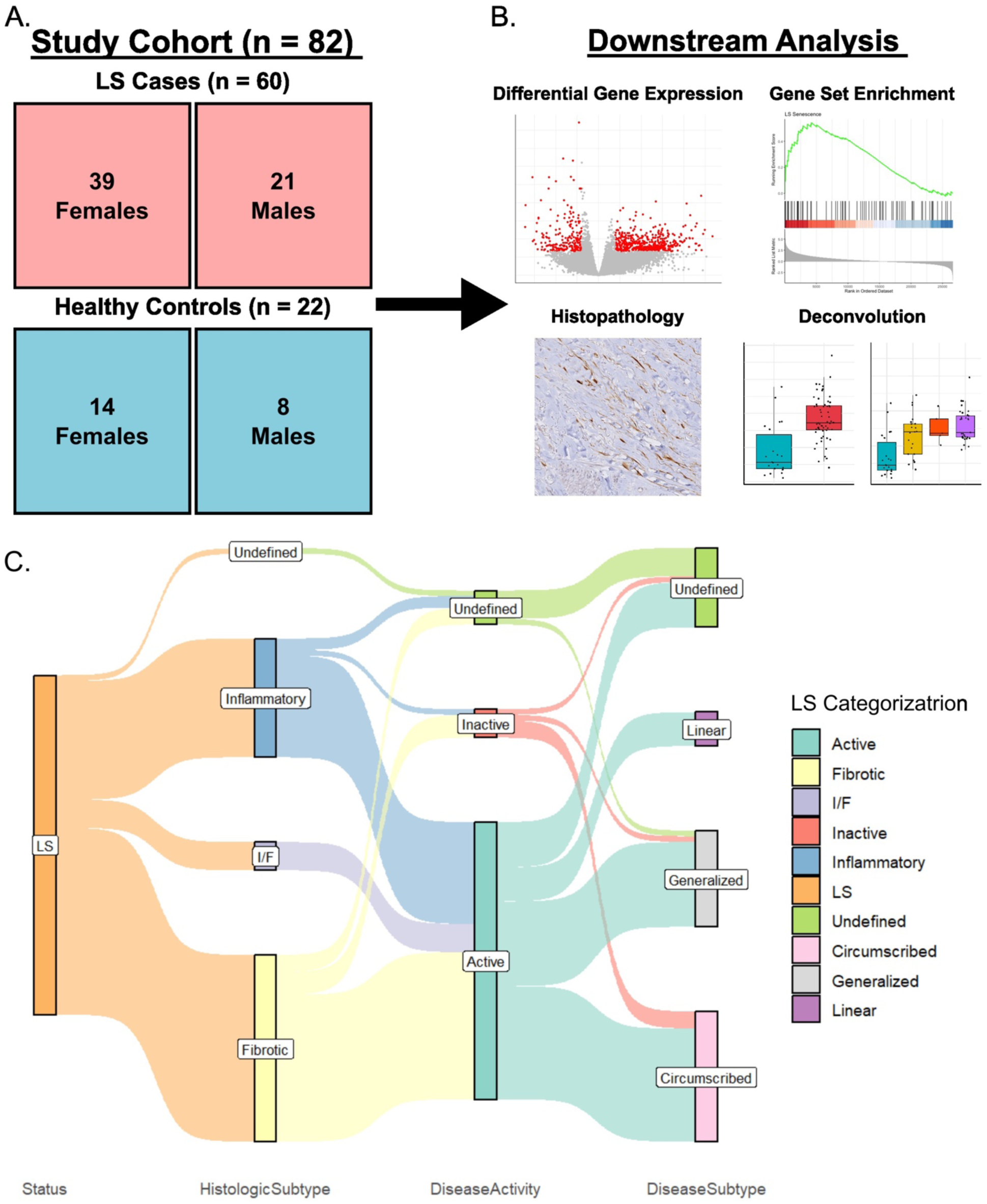
Study cohort, analysis workflow and overlap of clinical and histological classifications in LS. **Legend.** Panel A summarizes the study cohort of 60 adults with localized scleroderma and 22 healthy controls. Panel B is a flow diagram of the analyses: bulk RNA sequencing, differential expression and pathway analysis, cell-type deconvolution, immune-module scoring, senescence gene set analysis and p16^INK4a^ immunohistochemistry. Panel C is a Sankey diagram linking each lesion’s histological stage, clinical activity and clinical subtype.

## Results

### Cohort and sample characteristics

We analyzed lesional skin biopsies from 60 adults with LS and 22 healthy controls (HCs; **Tables 1 and 2**). The LS cohort had a mean age at biopsy of 52.5 ± 16.9 years and was predominantly female (39/60, 65%). The HC cohort had a mean age at biopsy of 58.9 ± 18.8 years and was also predominantly female (14/22, 63.6%). Clinical subtype, activity and histological stage are summarized in **Table 1** and **Figure 1c**. RNA yield averaged 28.8 ng/µl (range 0.4–110.0 ng/μl), RNA integrity numbers ranged from 1.5 to 3.1, as expected for archival formalin-fixed paraffin-embedded (FFPE) tissue, and sequencing yielded a mean of 41.4 million reads per sample (range 23–74 million).

**Table 1.** Demographic and Clinical Characteristics of LS Samples (n = 60) **Legend**. SD, standard deviation; n, number.

|  | **Mean ± SD** |  |
|---|---|---|
| **Age at the time of biopsy** | 52.5 ± 16.9 years |  |
|  | **Individuals (n)** | **Proportion (%)** |
| **Sex** |  |  |
| Female | 39 | 65 |
| Male | 21 | 35 |
| Disease Subtype |  |  |
| Linear | 6 | 10 |
| Circumscribed | 23 | 38 |
| Generalized | 17 | 28 |
| Undefined | 14 | 23 |
| Disease Activity |  |  |
| Active | 49 | 82 |
| Inactive | 5 | 8 |
| Undefined | 6 | 10 |
| Histologic Subtype |  |  |
| Inflammatory | 21 | 35 |
| Fibrotic | 33 | 55 |
| Mixed | 5 | 8 |
| Undefined | 1 | 2 |

**Table 2.** Demographic and Clinical Characteristics of HC Samples (n = 22)

|  | **Mean ± SD** |  |
|---|---|---|
| **Age at the time of biopsy** | 58.9 ± 18.8 years |  |
|  | **Individuals (n)** | **Proportion (%)** |
| **Sex** |  |  |
| Female | 14 | 64 |
| Male | 8 | 36 |

### Global transcriptional alterations in LS versus HC

Principal-component analysis (PCA) of log–counts-per-million (logCPM) values separated LS from HC along the first two components (**Figure 2a**). Differential expression identified 704 differentially expressed genes (DEGs) (488 upregulated, 216 downregulated) at |log₂ fold change (log₂FC)| ≥1 and false discovery rate (FDR) <0.05 (**Supplementary Tables S1–S3**). Upregulated genes included chemoattractants (*CCR5*, *CXCL11*, *CCL3L1*), interferon-signaling and antigen-presentation genes (*IFNG*, *TAP1*, *OAS3*, *HLA-DOB*, *HLA-DRB1*), T-cell activation markers (*STAT1*, *CD80*, *CD27*), type-2 mediators (*IL4I1*, *HAVCR1*), DNA-instability/oxidative-response genes (*PARP12*, *CYBA*, *SIRT4*), and senescence– and exhaustion-associated genes (*CDKN2A*, *PDCD1*, *LAG3*) (**Supplementary Table S2**). Downregulated genes were involved in oxidative stress (*MT1G*, *AHSP*), programmed cell death (*GREM2*, *FOSB*), negative regulation of transcription (*DUSP1*, *EGR1*), and extracellular matrix/adhesion (*SERPINA5*, *BMP3*, *PCOLCE2*) (**Supplementary Table S3**). Kyoto Encyclopedia of Genes and Genomes (KEGG) enrichment was led by cell-adhesion molecules and rheumatoid arthritis, with T-helper (Th)1/Th2 differentiation also enriched; Gene Ontology (GO) terms centered on lymphocyte and leukocyte activation and cell–cell adhesion (**Figure 2b, 2c**). Unsupervised clustering of the top 50 DEGs separated LS from HC into two distinct populations (**Figure 2d**).

**Figure 2.**
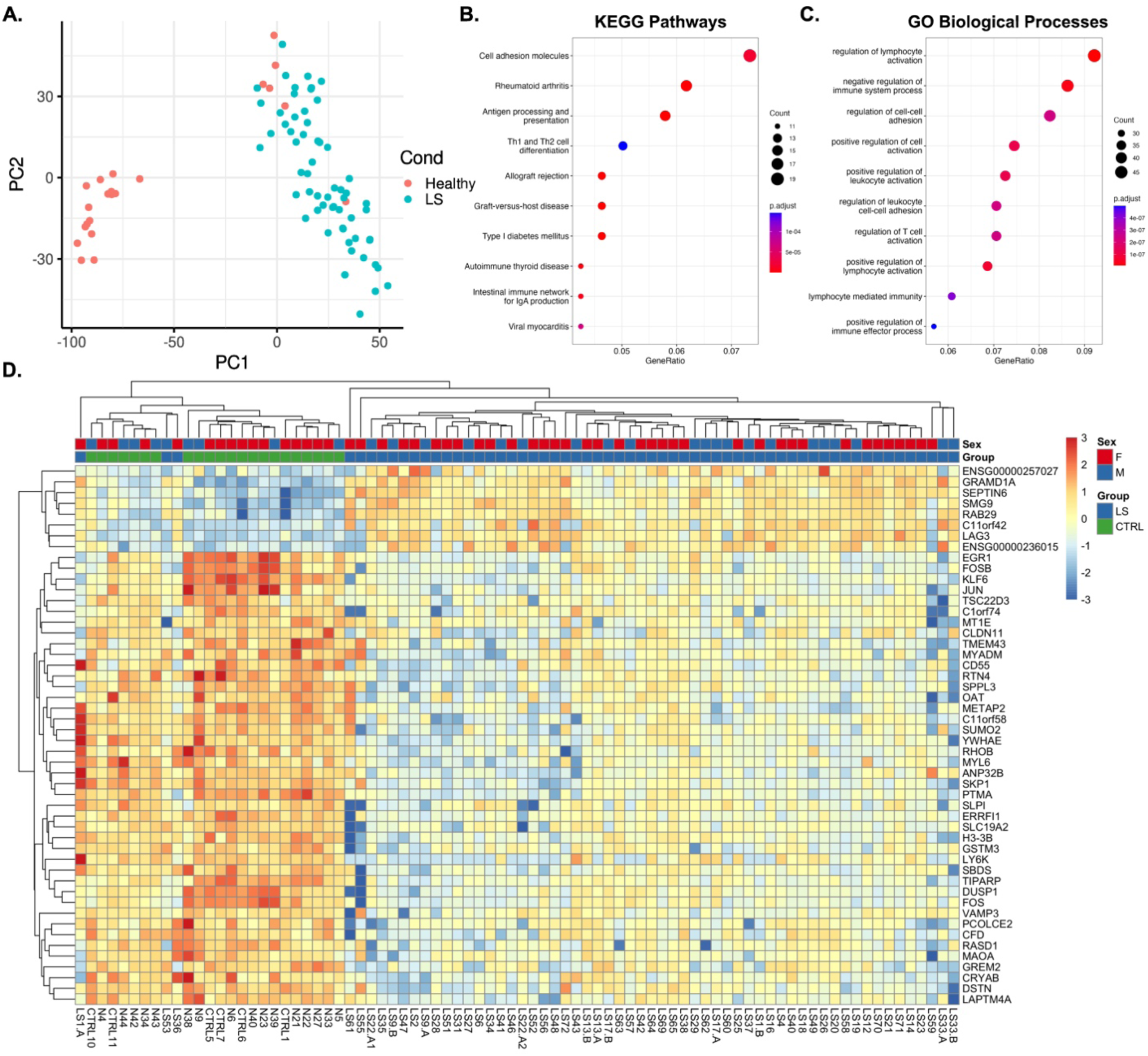
Dysregulation of transcription and pathway activity in LS *vs*. HC. **Legend.** Panel A is a principal-component plot in which localized scleroderma samples separate from healthy control samples. Panels B and C are dot plots of enriched KEGG and Gene Ontology pathways, led by immune activation, cell adhesion and T-helper differentiation terms. Panel D is a heatmap of the top differentially expressed genes, in which 58 of 60 localized scleroderma samples cluster apart from all healthy controls.

**Figure 3.**
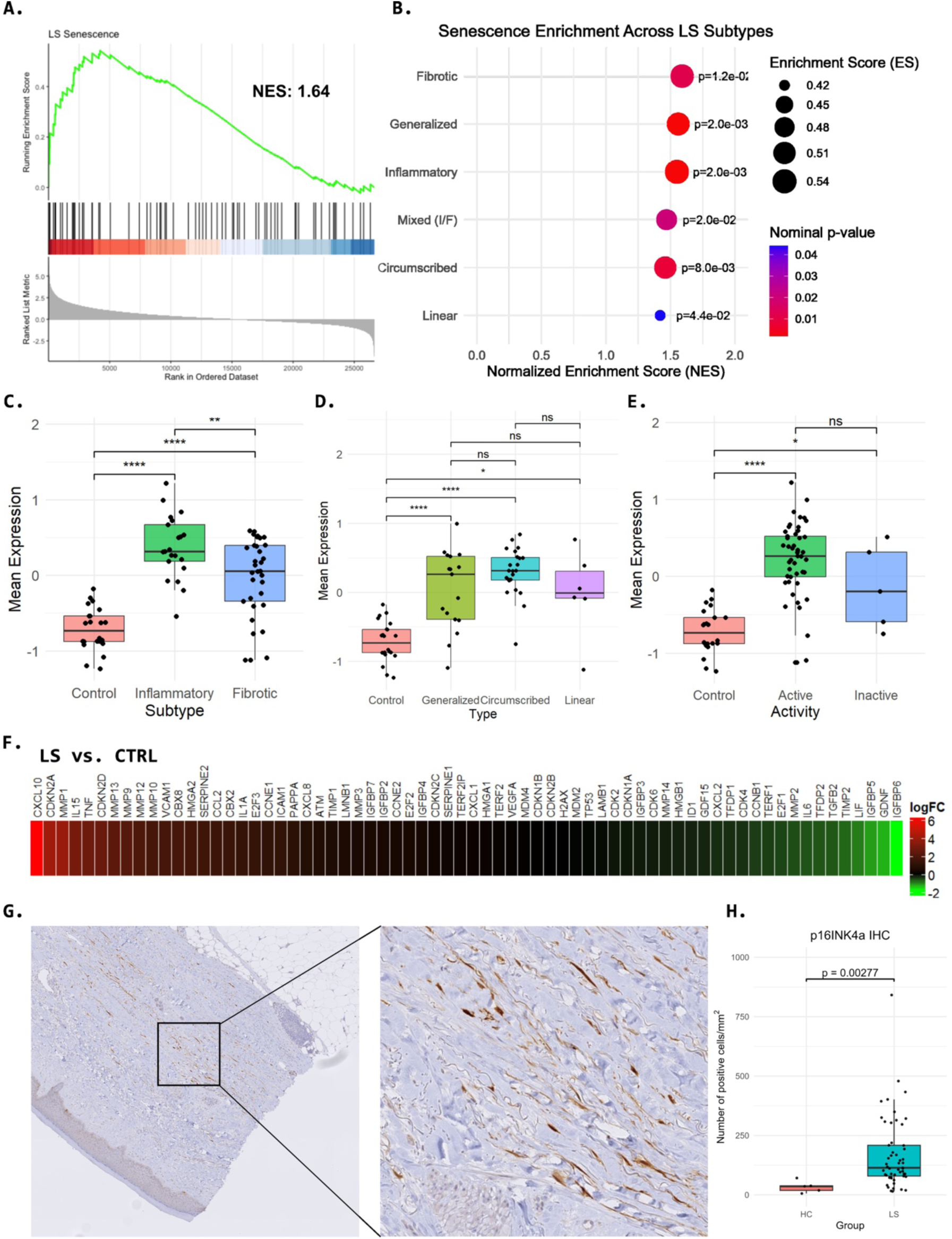
Senescence-associated gene expression and p16^INK4a^ immunohistochemistry in LS versus HC. **Legend.** Panels A and B are enrichment plots showing enrichment of a senescence-associated gene set in localized scleroderma overall and in each subgroup. Panels C to E are box plots showing higher mean expression of leading-edge senescence genes in disease subgroups than in healthy controls. Panel F is a heatmap of fold changes for senescence-associated genes. Panel G is a photomicrograph of p16INK4a-positive nuclei in lesional dermis, and panel H is a box plot showing higher p16INK4a-positive cell density in localized scleroderma than in healthy controls.

### Transcriptional changes vary with disease activity and histologic stage

Clustering of active LS (n = 49) versus HC produced clean separation, whereas inactive samples dispersed among active and HC clusters rather than forming a distinct group, possibly reflecting a transitional state (n = 5) (**Supplementary Figures S1, S2**). Active LS showed 607 upregulated and 247 downregulated genes versus HC (**Supplementary Tables S1, S4–S5**); 92% of the top 50 upregulated genes overlapped with the LS-versus-HC set, with *CCR5, MUC17, TMPRSS3 and XIRP1* ranking among the top 50 only in active disease (**Supplementary Table S4**), and KEGG enrichment of Th1/Th2/Th17 cell differentiation as well as antigen processing and presentation (**Supplementary Figure S1b**). GO process enrichment revealed enrichment for negative regulation of immune system process, leukocyte activation, and cell-cell adhesion (**Supplementary Figure S3**).

By histological stage, inflammatory LS separated completely from HC (**Supplementary Figure S4**), with 755 upregulated and 255 downregulated genes and newly implicated interferon-pathway genes (*STAT2*, *STAT4*, *IL12B*, *IRF9*), together with enrichments for Th1/Th2 cell differentiation, rheumatoid arthritis and lymphocyte activation (**Supplementary Tables S1, S13– S14; Supplementary Figure S5**). In contrast, fibrotic LS showed fewer transcriptional changes: two fibrotic samples clustered with HC and the remaining fibrotic samples formed a separate cluster (**Supplementary Figure S6**), with 103 upregulated and 82 downregulated DEGs (Supplementary Tables S1, S6–S7), isolated antigen-presentation (*TAP1*, *TAPBPL*) and interferon (*OAS3*) genes but no *HLA* induction, and absence of the type-1/type-2 signature seen in active and inflammatory disease (**Supplementary Tables S6, S7)**. Direct comparison of fibrotic and inflammatory lesions yielded 4 upregulated and 21 downregulated genes, with inflammatory lesions showing higher T-cell-receptor (*TRAV13-2*, *TRAV26-2*) and stress-response (*SELENOH*, *DNAJB12*) expression; only JAK–STAT signaling (*q* = 8.15×10^−3^) and transcriptional misregulation in cancer (*q* = 8.15×10^−3^) were enriched.

### Clinical subtype contrasts

Among clinical subtypes, circumscribed LS (n = 23) showed the most extensive changes (919 upregulated, 326 downregulated genes), generalized LS (n = 17) showed 52 upregulated (49 of 52 overlapping circumscribed) and 44 downregulated genes, and linear LS (n = 6) showed a single DEG (*IER5L*; *q* = 0.038, log₂FC 2.64) (**Supplementary Tables S8–S12)**. Circumscribed lesions uniquely upregulated cell-adhesion, T-helper, and antigen-presentation genes.

### Cell-type composition shifts in LS skin

Cell type deconvolution of bulk profiles against a single-cell skin reference inferred compositional shifts across disease states (**Figure 4, Supplementary Table S20)**. Keratinocytes, approximately 51.0% of cells in HC, were depleted in inflammatory (19.5%; *q* = 4.50×10^−5^) and fibrotic (24.5%; *q* = 1.20×10^−5^) LS. Fibroblasts rose from 17.7% in HC to 32.6% (inflammatory) and 39.5% (fibrotic; *q* = 1.60×10^−5^). Macrophages increased from 2.9% to 8.6% (inflammatory) and 5.9% (fibrotic), and dendritic cells from 0.43% to 1.74% and 1.29%, respectively. Regulatory T cells expanded markedly (1.2% in HC to 9.5% inflammatory, 6.0% fibrotic; *q* = 2.30×10^−11^), as did cytotoxic T cells (0.3% to 4.5% and 2.8%) and helper T cells (2.6% to 4.3% in inflammatory; not significant in fibrotic). Parallel shifts were seen in active and pooled LS (**Figure 4**).

**Figure 4.**
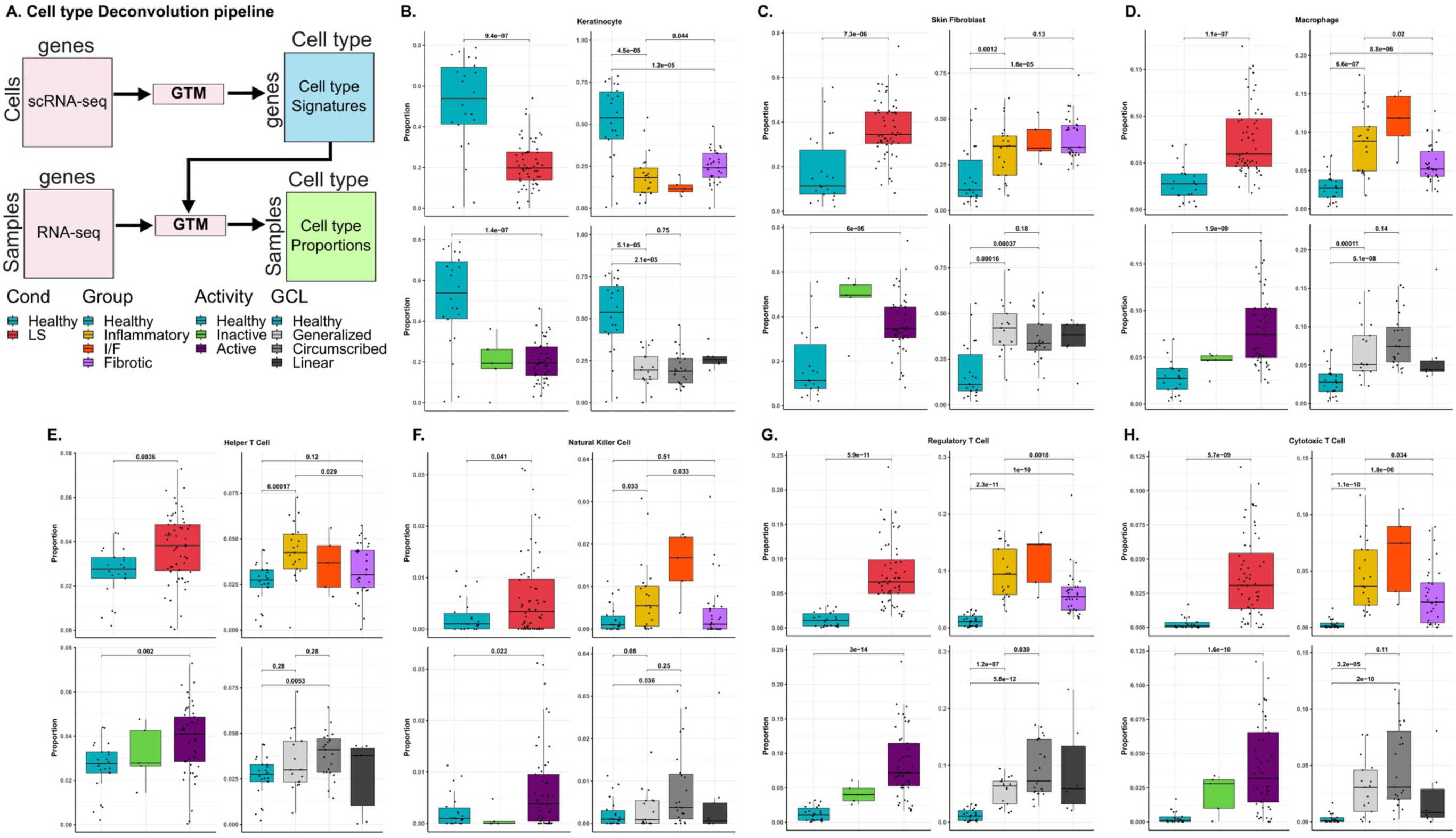
Deconvolution Pipeline and Differential Cell Type Abundance in LS *versus* HC. **Legend.** Panel A is a schematic of the deconvolution workflow combining a single-cell skin reference with bulk RNA sequencing data. Panels B to H are box plots of estimated cell-type proportions in healthy controls and localized scleroderma subgroups, showing fewer keratinocytes and more fibroblasts, macrophages and regulatory, cytotoxic and helper T cells in disease.

### Cell-type–specific transcriptional programs

Deconvolution-derived, cell-type–specific differential expression (**Figure 5; Supplementary Table S21)** revealed compartment-resolved programs. Keratinocytes downregulated stress-response mediators (*DUSP1*, log₂FC −2.55; *FOS*, −2.69) while upregulating interferon-stimulated genes (*GBP1*, *OASL*, *PLAU*). Fibroblasts showed a mixed fibrotic/immune/stress program (*THY1*, *PLCG2*, *SFMBT2*, *GBP1*, *PLAU*, *SEPTIN6* up; *KLF6*, *TSC22D3* down). In macrophages, the largest inferred fold change was for *CCL3L1* (log₂FC 10.61); *IL4I1* and *TNFRSF9* were also upregulated, consistent with a proinflammatory state. Regulatory T cells co-expressed suppressive/checkpoint markers (*TIGIT*, *TNFRSF9*) alongside interferon-stimulated transcripts, suggesting functional plasticity under inflammatory pressure. Cytotoxic T cells expressed a cytolytic-effector and exhaustion phenotype (*IFNG*, *GZMK*, *LAG3*, *SCML4*). Helper T cells upregulated inflammatory mediators including *TNF* and *TRIM22*. Concordance analysis against an independent LS single-cell dataset^7^ recovered overlapping DEGs in endothelial cells (*TNFRSF4*, *NLRC5*), macrophages (*TFEC*), pericytes (*FHL1*, *NR4A1*), and fibroblasts (*TSC22D3*, *PDGFRL*, *PTGIR*). Inferred cell-type proportions correlated with expression of canonical marker genes for most cell types. For helper T cells, this correlation was not significant (r = 0.154, *P* = 0.16); we therefore validated T-cell estimates by CD4 and CD8 immunohistochemistry (IHC), which showed higher densities of both cell types in LS than in HC (**Supplementary Figures S7–S8**).

**Figure 5.**
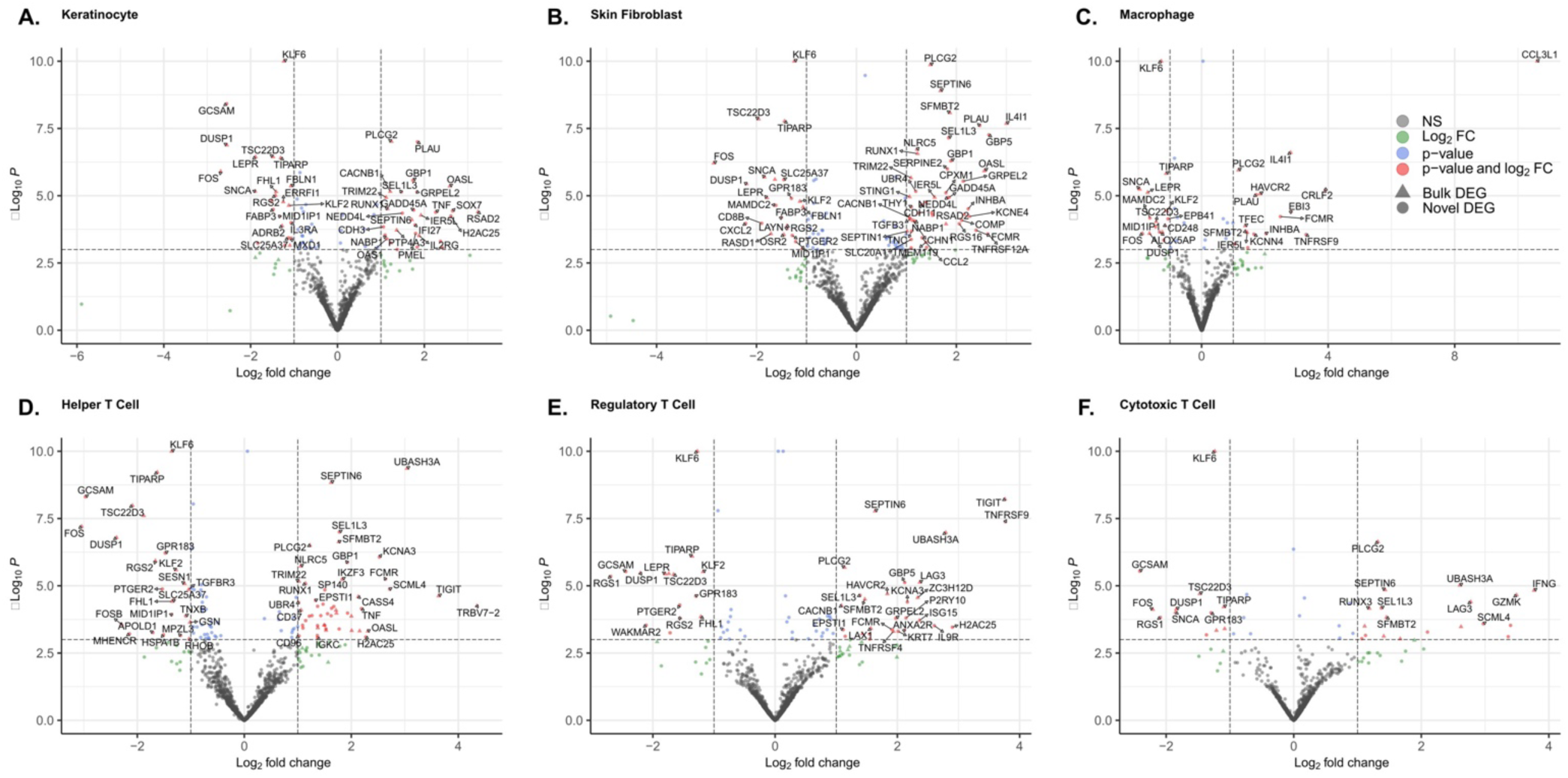
Cell type-specific differential gene expression in LS *versus* HC. **Legend.** Six volcano plots, one per cell type, plotting log2 fold change against significance for genes differentially expressed in localized scleroderma versus healthy controls. Triangles mark genes also found in the bulk analysis and circles mark genes found only in the cell-type-specific analysis.

### Immune-module activity

Immune-module scoring showed higher Th1 activity in LS overall than in HC (mean 0.0020; q = 8.10×10−5) and in inflammatory (0.0023), fibrotic (0.0016), active (0.0021) and circumscribed (0.0024) disease (**Figure 6**). Th2 and Th17 activity were higher in LS overall than in HC; differences in smaller subgroups were modest or not significant (e.g., generalized LS, Th2 q = 0.095 and Th17 q = 0.12). Th1 and Th17 activity were higher in inflammatory than fibrotic lesions, whereas Th2 activity was comparable between them (**Figure 6**).

**Figure 6.**
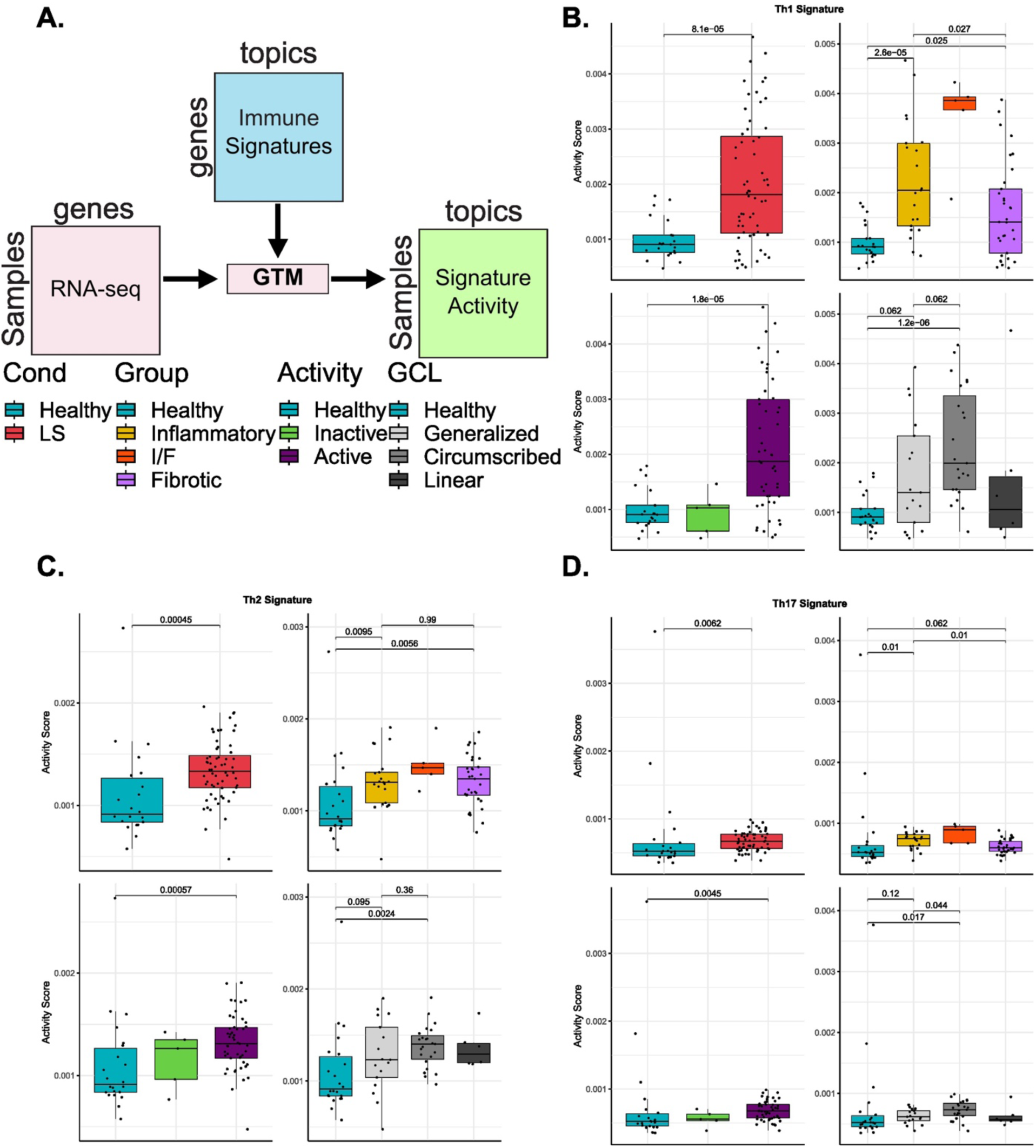
Th1/Th2/Th17 Immune Module Activity Across Clinical and Histological Subtypes of LS *versus* Healthy Skin. **Legend**. Panel A is a diagram of how immune-module activity was estimated. Panels B to D are box plots of Th1, Th2 and Th17 activity scores in healthy controls and in localized scleroderma grouped by histological stage, clinical activity and clinical subtype. Th1 and Th17 scores are highest in inflammatory lesions, whereas Th2 scores are similar in inflammatory and fibrotic lesions.

### Senescence-associated signature and p16^INK4a^ immunohistochemistry

LS lesions were enriched for a curated 68-gene senescence-associated gene set (Supplementary Table S19) relative to HC (NES = 1.64; *q* = 1.22×10^−3^), with leading-edge genes *CXCL10*, *CDKN2A*, *MMP1*, *IL15*, and *TNF* (**Figure 3a**). Nominally significant enrichment (P <0.05, unadjusted across subgroups) was observed in linear (NES 1.42), generalized (1.56), circumscribed (1.46), mixed (1.47), inflammatory (1.55), and fibrotic (1.59) LS (**Figure 3b**), the latter indicating that the signature is not confined to inflammatory disease; leading-edge genes also included *MMP13* and *CDKN2D*. Mean expression of leading-edge genes was higher in all LS subtypes than HC and higher in inflammatory than fibrotic LS (**Figure 3c–e**), and log₂-fold-change patterns are summarized by unsupervised clustering (**Figure 3f; Supplementary Figure S9**).

At the protein level, p16^INK4a^ immunohistochemistry restricted to dermal and hypodermal compartments (LS n = 52; HC n = 5) showed higher nuclear positivity in LS, with staining in fibroblasts, adipocytes, and inflammatory cells, most apparent in inflammatory and remodeling zones and largely absent in fully fibrotic tissue (**Figure 3g**). Quantitative image analysis gave a median density of 113.9 (IQR 77.9–213.8) p16^INK4a^-positive cells/mm² in LS versus 36.0 (IQR 12.3–54.4) in HC (P = 0.0028) (**Figure 3h**).

### Cross-disease positioning

Comparing bulk log-fold changes with published signatures from other inflammatory dermatoses^1,14^, LS senescence changes correlated most closely with cutaneous lupus erythematosus and lichen planus (Spearman ρ = 0.8 for both) and were opposite to Wells syndrome (ρ = −0.55); inflammatory-signal changes were positively correlated only with cutaneous lupus (ρ = 0.57), lichen planus (ρ = 0.56), and chronic spontaneous urticaria (CSU; ρ = 0.52) (**Supplementary Figure S10**).

## Discussion

This study provides a cell-resolved transcriptomic and histologic map of adult LS. Its principal finding is that adult LS lesional skin shows type-1/type-17 T-cell and interferon-associated programs that are most pronounced in clinically active and histologically inflammatory lesions and attenuated in fibrotic lesions, whereas type-2 (Th2) activity persisted across histological stages, together with a senescence-associated signature supported by increased dermal p16INK4a-positive cells. Rather than two discrete endotypes, the data are consistent with a continuum related to disease activity, consistent with the active-versus-inactive dichotomy repeatedly observed in pediatric LS^2,6^ and with the interferon-γ/Th1-dominant, fibrosis-sparse molecular identity recently described for morphea^9^.

Several features extend prior work. First, the cohort is the largest adult LS transcriptomic series to date and directly addresses the adult knowledge gap left by predominantly pediatric studies; the macrophage expansion we inferred is consistent with the macrophage-centered adult program identified by single-cell analysis^10^; however, the strong interferon and T-cell signal in clinically active lesions indicates that adult LS is not uniformly less inflammatory than pediatric disease. Our data also extend our previous findings of combined type-2 inflammation and immunosenescence^8^. Second, deconvolution localizes the inflammatory signal to expanded and activated T-cell, macrophage, and fibroblast compartments and identifies co-expression of effector and checkpoint/exhaustion markers (*LAG3* and *TIGIT*) in inferred cytotoxic and regulatory T-cell profiles, together with bulk upregulation of *PDCD1*, suggesting chronic antigenic stimulation with partial exhaustion.

Senescence emerged as a feature of both inflammatory and fibrotic disease. A senescence-associated signature was enriched across all subgroups, including fibrotic lesions, and p16^INK4a^-positive cells were increased, most prominently in inflammatory and remodeling zones. Because leading-edge genes include senescence-associated secretory phenotype (SASP) factors (*CXCL10*, *IL15*, *TNF*, *MMP1*, *MMP13*), this signature most likely captures a feed-forward circuit between senescence and inflammation rather than senescence alone. Resident cells appear to be active participants: keratinocyte and fibroblast profiles expressed interferon-stimulated genes, and p16^INK4a^ localized to fibroblasts and adipocytes as well as to immune cells. Expanded regulatory T cells co-expressed checkpoint and interferon-stimulated transcripts, consistent with an inflammation-adapted, less suppressive state; SASP-rich milieus impair regulatory T-cell function in autoimmunity^13^, which may explain why their expansion fails to resolve disease. Together with self-DNA sensing and cGAS–STING activation described in morphea^15^ and the induction of DNA damage-response and oxidative-stress genes in our cohort (*PARP12*, *CYBA*, *SIRT4*), these observations support a testable model in which tissue injury triggers interferon-driven T-cell activation, exhaustion and senescence, while persistent type-2 and SASP programs sustain fibrosis. This parallels the accumulation of senescent cells in systemic sclerosis skin^12^ and cytotoxic tissue-resident memory T-cell injury in established morphea^11^; whether senescence is a cause or a consequence of inflammation cannot be resolved cross-sectionally. Finally, the proximity of LS to cutaneous lupus and lichen planus, and its divergence from Wells syndrome, supports an interface-dermatitis-like, interferon-driven rather than eosinophil-driven biology.

These observations have direct translational implications. Interferon and JAK–STAT signaling were prominent in clinically active and histologically inflammatory lesions, and JAK– STAT was one of only two pathways enriched in the direct comparison of inflammatory with fibrotic lesions. This provides a mechanistic rationale for JAK inhibition, which has produced encouraging responses in refractory LS, with improvement reported in 15 of 17 published cases^16^, and which is reinforced by shared JAK–STAT biology between morphea and eosinophilic fasciitis^15^. Our data suggest that patients with clinically active, histologically inflammatory disease, in whom the interferon/Th1 program is most pronounced, are the most likely to benefit, and support prospective trials of JAK inhibitors in this population. By contrast, Th2 activity was present across LS and did not differ between inflammatory and fibrotic lesions, consistent with a profibrotic role of IL-4/IL-13 beyond the inflammatory phase. Type-2 blockade alone may nonetheless be insufficient: in a phase IIa trial of dupilumab in plaque-type and generalized morphea (EudraCT 2019-002036-90), the week-24 response rate did not differ significantly from placebo (57% vs 40%; P = 0.29). Because type-1 and type-2 programs coexist in active lesions, broader inhibition, such as with JAK inhibitors, may be preferable. The senescence-associated program provides a rationale for senescence-modulating therapies, by analogy to first-in-human senolytic studies in fibrotic disease^17^. Glucagon-like peptide-1 receptor agonists, which have anti-inflammatory and senescence-modulating effects in certain diseases^18^, are a pragmatic candidate given their wide availability; improvement of linear scleroderma with semaglutide has been reported^19^, and semaglutide is also being evaluated in systemic sclerosis^20^. Finally, the T-cell checkpoint/exhaustion signature raises the possibility of immune-modulatory strategies.

### Limitations

This study has limitations. First, it is retrospective, cross-sectional and single-centre, which precludes causal and temporal inference and limits generalizability to other populations and to pediatric disease. Second, HC transcriptomes were derived from separately collected and sequenced specimens, previously used as controls in our CSU study^14^, and were integrated after ComBat batch correction. Because HC status and sequencing batch largely coincide, batch correction cannot fully separate technical from biological variation, and differences in tissue procurement (surgical skin redundancy versus diagnostic biopsy), anatomical site and biopsy depth may contribute to some LS–HC differences, including immediate-early and stress-response genes (*FOS*, *JUN*, *EGR1*, *DUSP1*, *KLF6*) and inferred keratinocyte proportions. Comparisons between LS subgroups, such as inflammatory versus fibrotic lesions, are less affected by this limitation. Likewise, because CSU was compared with the same HC, its correlation with LS may be partly inflated by the shared controls. Third, RNA was derived from archival FFPE tissue with low RNA integrity numbers (1.5–3.1); although the percentage of RNA fragments >200 nucleotides (DV200)-based quality control was applied, degraded input can bias detection. Fourth, the senescence gene set overlaps with inflammatory and cell-cycle genes, p16^INK4a^ can be induced by non-senescent stress, and the IHC comparison included only five HC specimens; the senescence findings should therefore be regarded as senescence-associated. Fifth, cell-type proportions and cell-type–specific expression were inferred computationally using a reference of healthy, atopic dermatitis and psoriasis skin that did not include LS tissue, which may incompletely capture disease-specific states. Finally, subgroup analyses were constrained by uneven sample sizes (notably the linear, inactive and mixed groups), clinical activity and histological stage overlapped, missing clinical metadata created undefined categories, and treatment exposure at biopsy was not captured. Future studies with prospectively collected, site-matched controls processed in parallel, together with spatial transcriptomics, multiplexed imaging and functional assays, are needed to confirm these findings.

### Conclusion

Adult localized scleroderma lesional skin shows type-1/type-17 T-cell activation and interferon signaling that are most pronounced in clinically active and histologically inflammatory disease, together with a senescence-associated signature and increased dermal p16INK4a-positive cells. Type-2 and senescence-associated programs persist in fibrotic lesions. These findings support JAK inhibition in active disease and clinical evaluation of senescence-modulating strategies, including glucagon-like peptide-1 receptor agonists, across disease stages.

## Materials and Methods

### Study cohort and samples

We retrospectively analyzed archived FFPE lesional skin biopsies from 60 adults (≥18 years) with clinically and histologically confirmed LS (McGill University Health Centre LS registry and rare inflammatory skin diseases biobank) and 22 HC of similar age and sex distribution. Reporting follows the STROBE guideline for observational studies. LS was classified as circumscribed, linear, generalized or mixed according to the Padua classification^21^, and graded histologically as inflammatory, fibrotic, or mixed by two blinded dermatopathologists using hematoxylin-and-eosin slides, with discrepancies resolved by consensus^8^. Active disease was defined as Localized Scleroderma Skin Activity Index and/or Physician Global Assessment of Activity >0^22^. HC RNA-seq data were generated in our laboratories from histologically normal adult skin from sun-protected sites, comprising unpublished skin-redundancy tissue and previously published datasets; the same HC were used as the comparator group in our CSU study^14^. HC specimens underwent the same RNA extraction, library preparation and sequencing protocols as LS specimens. The study was approved by the McGill University Health Centre Research Ethics Board (REB 2021-7412), with a waiver of informed consent for anonymized archived tissue.

### RNA extraction and sequencing

Four 10-µm FFPE sections were processed with the RNeasy FFPE Kit (Cat. No. 73504, Qiagen, Hilden, Germany); yield and quality were assessed by NanoDrop and Agilent Bioanalyzer, and only samples with DV200 ≥36.5% were sequenced. Libraries were prepared using TruSeq RNA Access Library Prep Kit and sequenced on the Illumina NovaSeq 6000 platform (paired-end, 2 × 150 bp), targeting ∼37.5 million reads per sample.

### Bioinformatic processing and bulk differential gene expression

FASTQ files were processed with the GenPipes RNA-seq pipeline v4.4.5; reads were trimmed with Trimmomatic v0.36, aligned to GRCh38 with STAR v2.7.8a and quantified with HTSeq v2.0.4 using GENCODE v38 annotations^23^. Differential gene expression was performed using limma^24^ with batch as a covariate, and DEGs were defined by |log₂FC| ≥1 and Benjamini– Hochberg FDR <0.05. Contrasts compared LS, each clinical activity group, histological stage and clinical subtype with HC, and inflammatory with fibrotic lesions directly; results for mixed-stage and inactive lesions are provided in **Supplementary Tables S15–S18**. Pathway enrichment analysis was conducted using clusterProfiler (Gene Ontology, KEGG)^25^; dimensionality reduction used prcomp function in R version 4.2.2 on logCPM-normalized expression values; top-50-DEG clustering used pheatmap package^26^.

### Gene-set enrichment

Senescence-associated enrichment was assessed by preranked gene set enrichment analysis (fgsea R package; genes ranked by log2 fold change) using a curated set of 68 senescence-associated genes derived from Shi et al. (**Supplementary Table S19)**^12,27^, with NES quantifying activity (NES > 1 indicating positive enrichment relative to background expression). Each LS subgroup was compared with HC separately, and nominal P values are reported for subgroup analyses. A per-sample score was calculated as the mean expression of leading-edge genes.

### Cross-disease transcriptomic comparison

LS-versus-HC log2 fold changes were compared with published differential expression data from other inflammatory dermatoses, including lichen planus, atopic dermatitis, psoriasis, neutrophilic dermatoses, Wells syndrome and cutaneous lupus erythematosus^1^, and with CSU profiled by our group using the same pipeline and HC^14^. Because the published datasets were generated with a targeted NanoString panel, comparisons used log2 fold changes of genes measured in both datasets. Pairwise Spearman correlations and PCA were computed on disease-level log2 fold-change vectors for the inflammatory signature defined by Seremet et al. and the senescence-associated gene set (**Supplementary Table S19).**

### Cell-type and immune-module deconvolution

Cell composition and cell-type–specific expression were inferred with an Rcpp implementation of the guided topic model (GTM)-based method GTM-decon (SpaTM-G)^28,29^, trained on a single-cell skin reference (∼195,000 cells)^30^. Cell-type–specific DESeq2 was restricted to cell types with ≥1% median proportion, with batch and cell-type proportion as covariates and log-fold-change shrinkage^31^. Th1/Th2/Th17 modules were defined from curated markers^1^. An additional background module composed of non-DEGs (|log2FC| < 1 & p > 0.05) was included to regularize the inferred activity scores. Group comparisons used Mann–Whitney U tests with Benjamini–Hochberg correction; groups with fewer than 10 samples are shown descriptively. Deconvolution quality was assessed by Spearman correlation between inferred cell-type proportions and normalized expression of canonical cell-type markers and, for T cells, by CD4 and CD8 immunohistochemistry.

### Immunohistochemistry and image analysis

FFPE sections (4 µm) were stained for p16^INK4a^ using the CINtec p16 Histology Kit (Roche Diagnostics, Mannheim, Germany). CD4 (CD8) immunohistochemistry was performed on 53 (55) LS and 5 (5) HC sections. Slides were analyzed in QuPath v0.6.0^32^. Dermal and hypodermal regions were annotated (epidermis excluded); positive nuclei were detected with the positive cell detection algorithm with the same settings as in our CSU study^14^ and expressed as positive cells/mm² of annotated tissue. CD4+ and CD8+ cells were quantified in the same way. The five HC sections were the same specimens used in that study.

### Statistics

Analyses used R v4.2.2. Continuous variables were compared with Mann–Whitney U or Kruskal–Wallis tests; P values were adjusted using the Benjamini–Hochberg FDR method within each family of comparisons, and two-sided q <0.05 was considered significant. Analyses were exploratory, and no formal sample size calculation was performed. Data visualization was performed with ggplot2^33^.

## Author contributions (CRediT)

**LK:** Investigation, formal analysis, methodology, writing – original draft, writing – review & editing; **AO:** Formal analysis, methodology, software, visualization, writing – original draft, writing – review & editing; **EM-J:** formal analysis, visualization, writing – original draft; **DV:** formal analysis, visualization, writing – review & editing; **AR:** Investigation, Validation; **AA:** Investigation, Validation; **MC:** Investigation, Validation; **MO:** Conceptualization, Methodology; **PL:** Methodology, Resources; **LX:** Resources; **YL:** supervision, resources, writing – review & editing; **EN:** Funding acquisition, conceptualization, supervision, resources, formal analysis, writing – original draft, writing – review & editing.

AR, AA, and MC reviewed pathology slides and assisted with sample acquisition. MO contributed to planning the senescence analyses. PL contributed to experimental planning and provided control samples. LX provided control samples.

## Use of artificial intelligence

During manuscript preparation, the authors used Claude (Anthropic; model claude-opus-5-5; accessed 29 October and 20 November 2026) to review the manuscript before submission and to suggest edits to wording, structure and the description of Methods, which the authors reviewed, verified and accepted or rejected. No artificial intelligence tool was used for data generation, analysis, interpretation, or to create or modify figures. The authors take full responsibility for the content of the manuscript.

## Acknowledgements

We acknowledge the McGill Genome Centre and the Canadian Centre for Computational Genomics (C3G), a Genomics Technology Platform supported by the Canadian government through Genome Canada. **Funding.** The C-MORE registry and biobank creation used in this study were supported by the Canadian Drug Agency and SkiN Canada. E.N. holds a Junior 1 Clinician-Scientist award from the Fonds de recherche du Québec – Santé (FRQS). A.O. is supported by a training scholarship from the Canadian Institutes of Health Research. D.V. is supported by a training scholarship from FRQS.

## Data availability

The data supporting this study’s findings will be deposited in NCBI GEO upon publication, subject to approval from the McGill University Health Centre Research Ethics Board, and are available from the corresponding author on reasonable request. Code is available at github.com/aosakwe/LS_GTM.

## Ethics statement

All specimens were obtained under institutional research ethics board approval (REB 2021-7412) with a waiver of informed consent for the use of anonymized archived tissue.

## What is already known about this topic?

- Transcriptomic studies of localized scleroderma (morphea), conducted largely in children, link active disease to interferon-γ/Th1 chemokine signatures.
- Cellular senescence has been implicated in systemic sclerosis skin, but adult localized scleroderma remains less well characterized at the tissue level.

## What does this study add?

- In 60 adults, lesional transcriptional changes were extensive in clinically active and histologically inflammatory disease and limited in fibrotic lesions, with interferon-associated T-cell activation and Th1/Th17 module activity, and Th2 activity across histological stages.
- A senescence-associated gene signature and increased dermal p16^INK4a^-positive cells were observed, including in fibrotic lesions.

## What is the translational message?

- Clinically active, histologically inflammatory adult morphea shows interferon/JAK–STAT-associated T-cell activation, providing a mechanistic rationale for JAK inhibitors in this subgroup.
- A senescence-associated program, present even in fibrotic lesions, identifies senescence-modulating agents, such as glucagon-like peptide-1 receptor agonists, as candidates for clinical trials.

## Abbreviations

CSU: chronic spontaneous urticaria
DEG: differentially expressed gene
DV200: percentage of RNA fragments >200 nucleotides
FDR: false discovery rate
FFPE: formalin-fixed paraffin-embedded
GO: Gene Ontology
GSEA: gene set enrichment analysis
GTM: guided topic model
HC: healthy control
IHC: immunohistochemistry
JAK–STAT: Janus kinase–signal transducer and activator of transcription
KEGG: Kyoto Encyclopedia of Genes and Genomes
log₂FC: log₂ fold change
logCPM: log counts per million
LS: localized scleroderma
NES: normalized enrichment score
PCA: principal component analysis
RNA-seq: RNA sequencing
SASP: senescence-associated secretory phenotype
Th: T-helper

