## Supplemental Figures and Tables for "Interferon-driven T-cell activation and a senescence-associated signature characterize clinically active adult localized scleroderma"

**Supplementary Materials**

### Supplementary Figures

**Supplementary Figure 1.** Unsupervised hierarchical clustering of top 50 genes in LS subgroups *vs.* HC. The color key indicates gene expression as a z-score

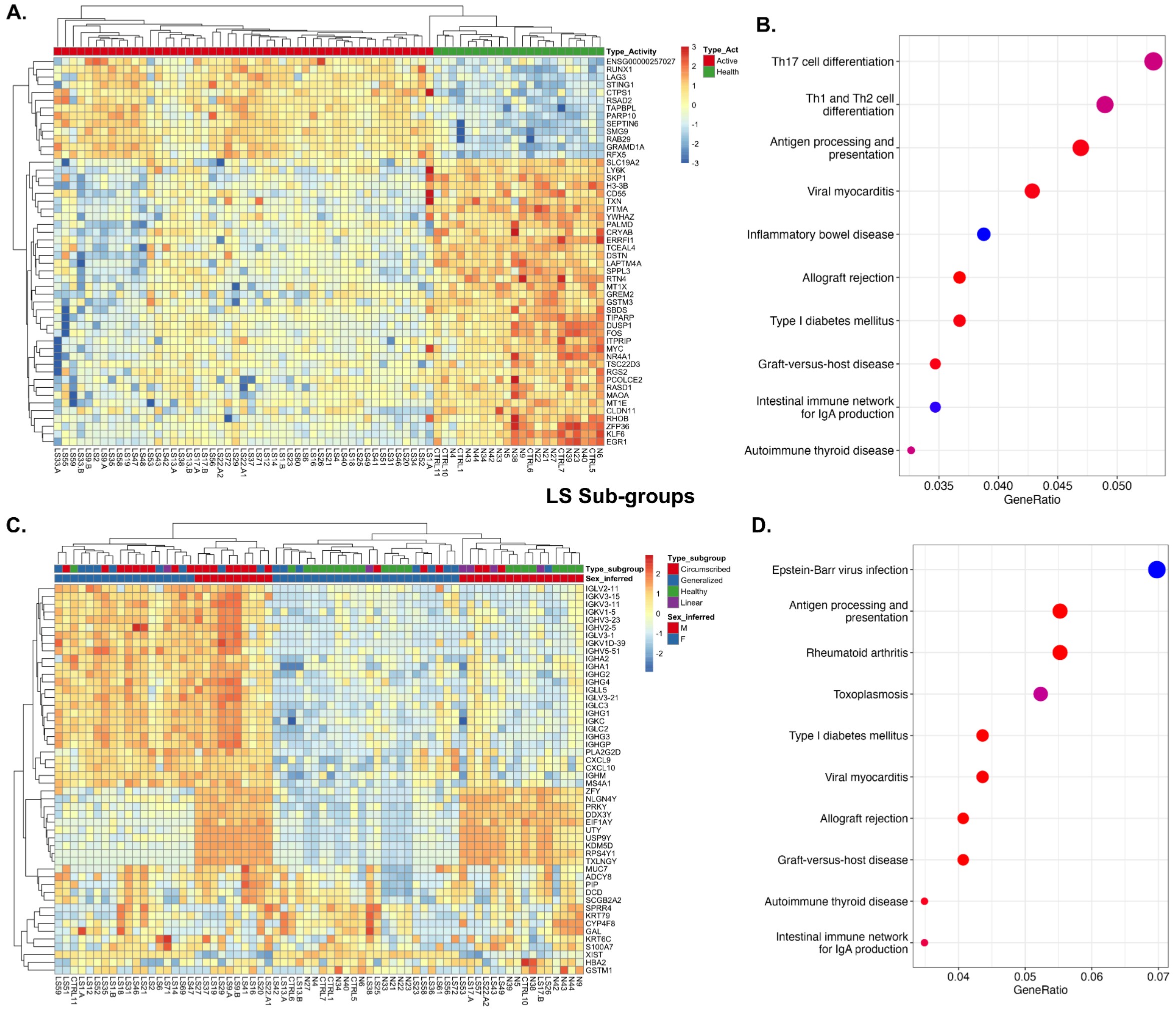


**Legend. (**A) Active LS samples (n = 49) are shown in red, HC (n = 22) in green. Two clusters were identified, the first containing all 49 active LS and the second containing all 22 HC samples. LS, localized scleroderma; HC, healthy controls. (B) KEGG Pathway enrichments for Active LS**.** (C) Circumscribed (n = 23) are shown in red, generalized in blue (n = 17), linear in purple (n = 6), and HC in green (n = 22). LS, localized scleroderma; HC, healthy controls. (D) KEGG pathway enrichments for Circumscribed LS.

**Supplementary Materials**

**Supplementary Figure 2.** Unsupervised hierarchical clustering of top 50 genes in active vs. inactive LS vs. HC. The color key indicates gene expression as a z-score.

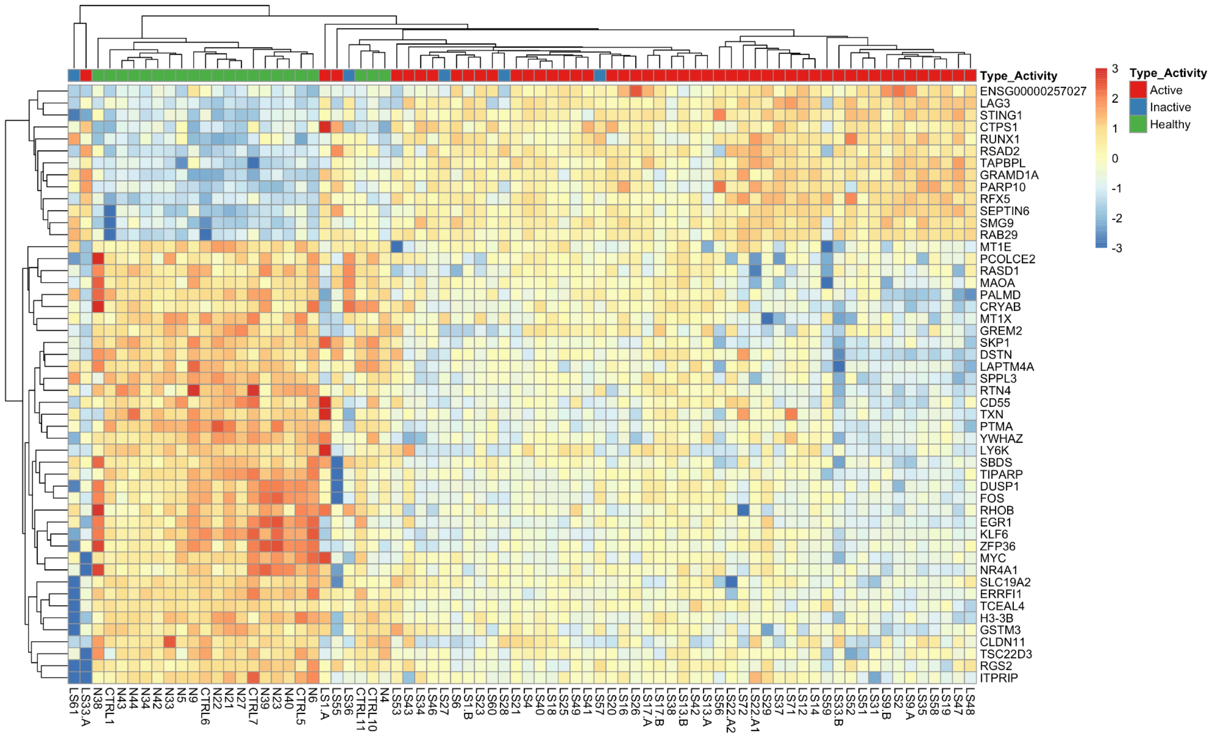


**Legend**. Active (n = 49), inactive LS samples (n = 5) are shown in red and blue respectively, HC (n = 22) in green. Three clusters were identified, the first containing one active and inactive sample, the second containing 19 HC samples, and the third containing 4 inactive LS, 3 HC, and 48 active LS samples. LS, localized scleroderma; HC, healthy controls.

**Supplementary Figure 3.** Upregulated GO Pathways in Active LS vs. HC

**Supplementary Materials**

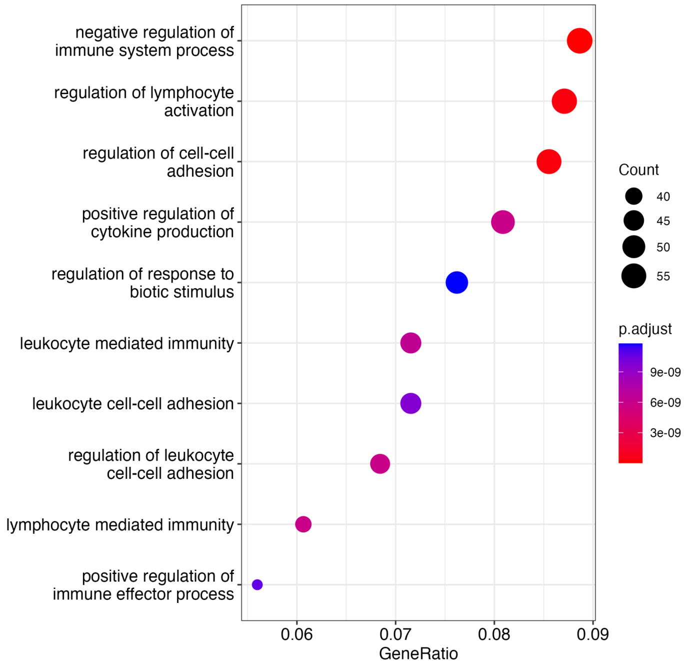


**Legend**. GO pathway enrichment analysis visualized in a dot plot. The GeneRatio indicates the proportion of genes in our dataset associated with each specific pathway, allowing us to identify pathways with a higher level of involvement from our dataset. The size of the dots represents the count of genes, while the color gradient reflects the Q-value.

#### Supplementary Materials

**Supplementary Figure 4.** Unsupervised hierarchical clustering of top 50 genes in Inflammatory LS vs. HC. The color key indicates gene expression as a z-score.

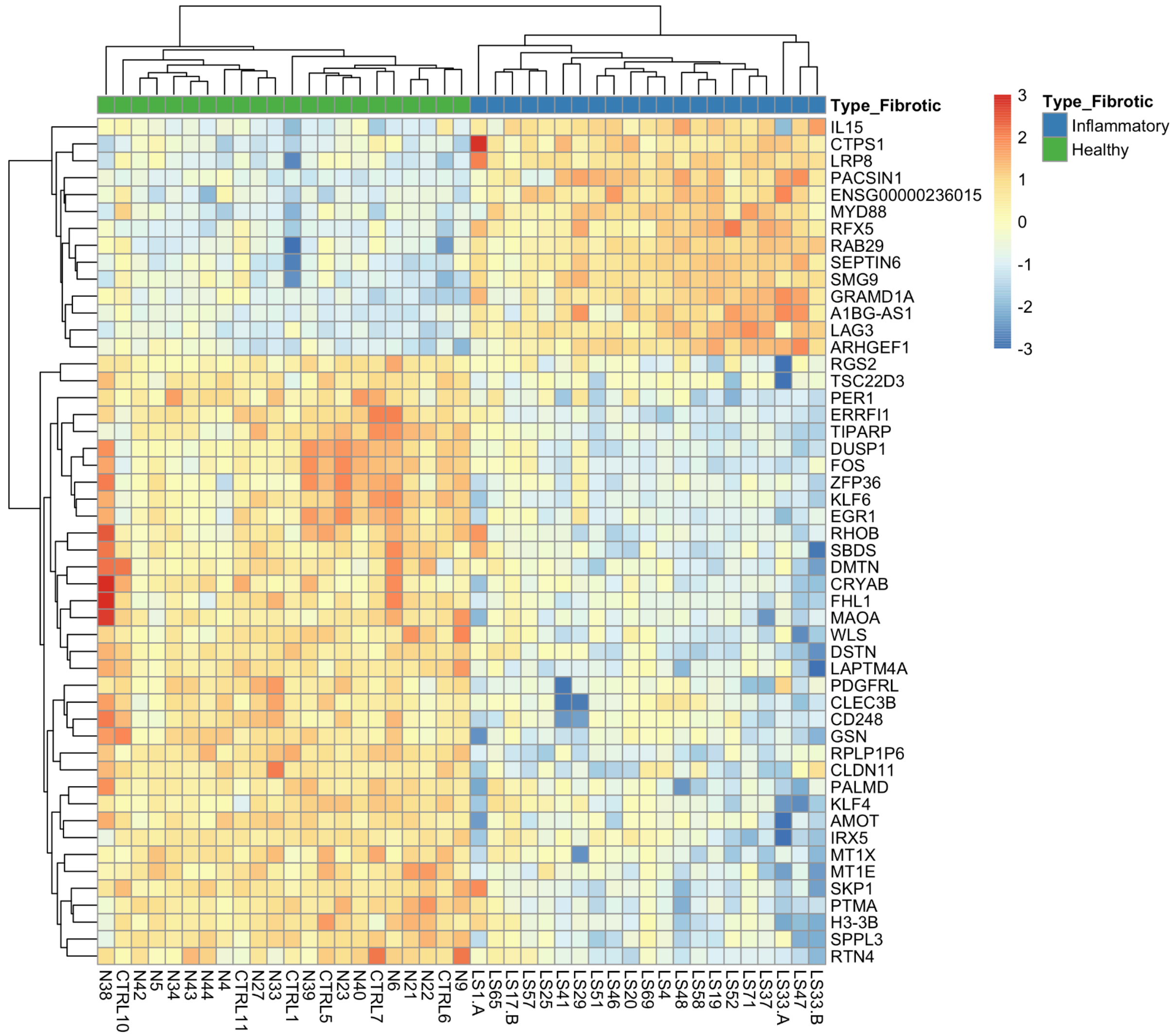


**Legend**. Inflammatory LS samples (n = 21) are shown in blue and HC (n = 22) in green. Two clusters were identified with all samples of each group clustering separately from each other. LS, localized scleroderma; HC, healthy controls.

**Supplementary Materials**

**Supplementary Figure 5.** The KEGG and GO upregulated pathways in Inflammatory *vs.* HC.

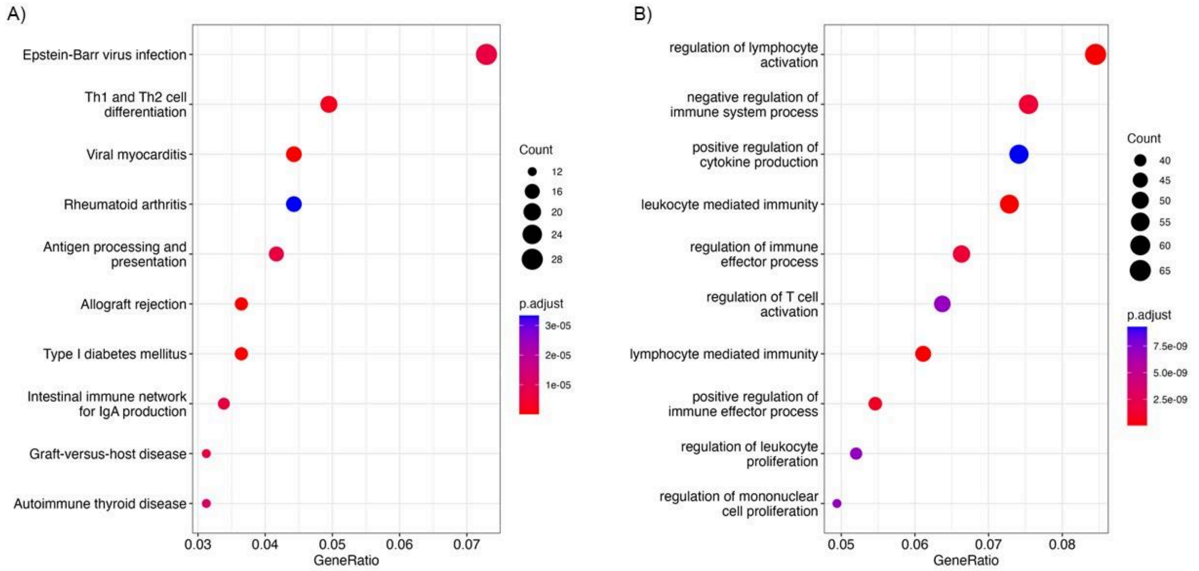


**Legend**. A) KEGG and B) GO pathway enrichment analysis visualized in dot plots. The GeneRatio indicates the proportion of genes in our dataset associated with each specific pathway, allowing us to identify pathways with a higher level of involvement from our dataset. The size of the dots represents the count of genes, while the color gradient reflects the Q-value.

**Supplementary Figure 6.** Unsupervised hierarchical clustering of top 50 genes in Fibrotic LS vs. HC. The color key indicates gene expression as a z-score.

#### Supplementary Materials

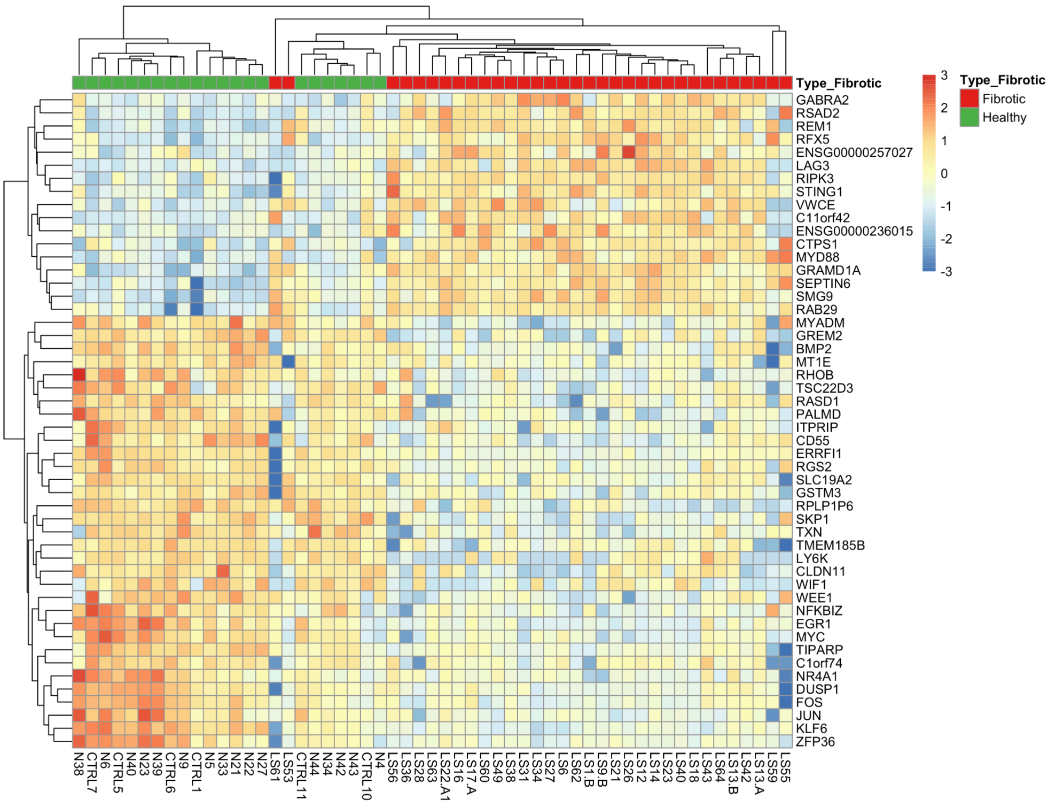


**Legend**. Fibrotic LS samples (n = 33) are shown in red and HC (n = 22) in green. Two clusters were identified. LS, localized scleroderma; HC, healthy controls.

**Supplementary Figure 7.** Expression-based validation of cell type deconvolution.

#### Supplementary Materials

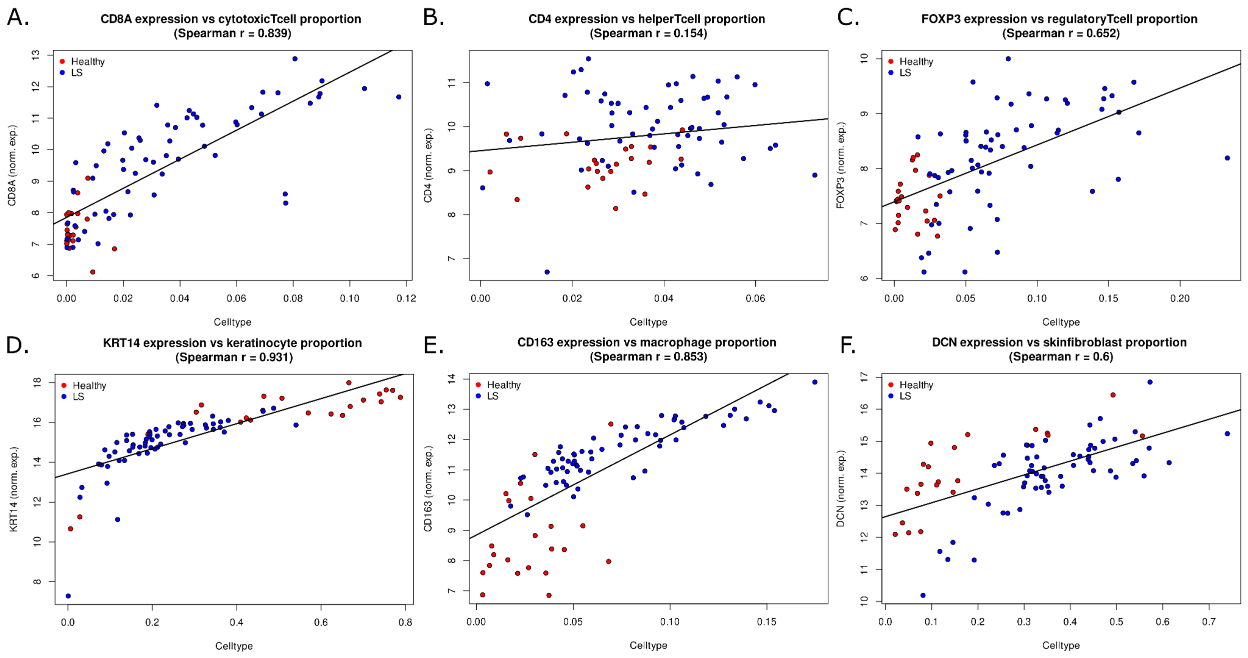


**Legend:** Spearman correlations between predicted cell type proportions and corresponding normalized marker gene expression in the LS and healthy control bulk samples.

**Supplementary Figure 8.** Immunohistochemistry validation of CD4+ and CD8+ T cell deconvolution.

**Supplementary Materials**

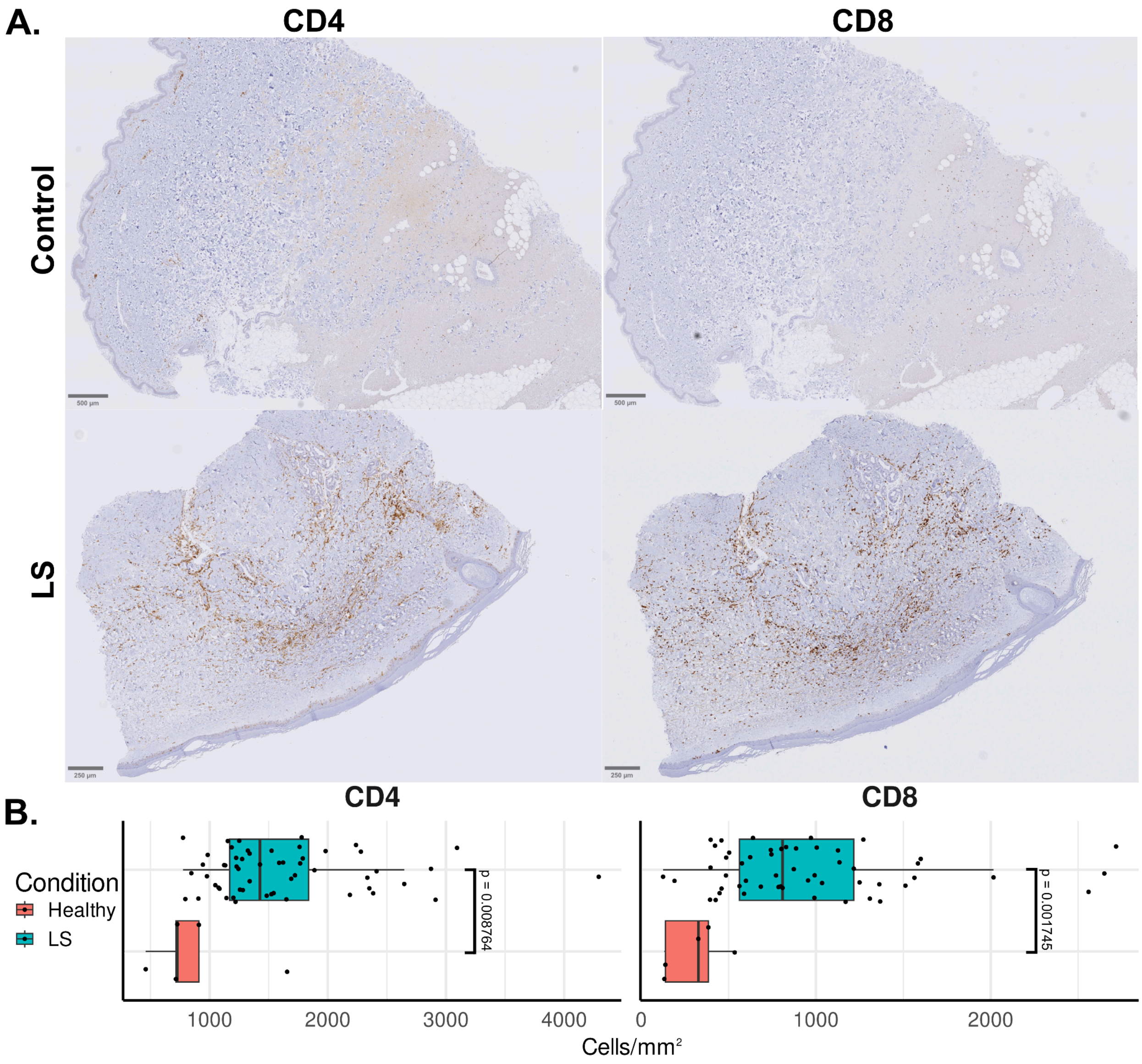


**Legend:** Immunohistochemistry analysis of helper and cytotoxic T cell abundance in LS and healthy control samples. (A) Representative IHC stains for CD4+ and CD8+ cells in LS and healthy controls. (B) QuPath-derived positive cell densities estimated for CD4+ and CD8+ cells for both conditions.

**Supplementary Materials**

**Supplementary Figure 9.** Unsupervised hierarchical clustering of top 50 senescent genes in LS vs. HC. The color key indicates gene expression as a z-score.

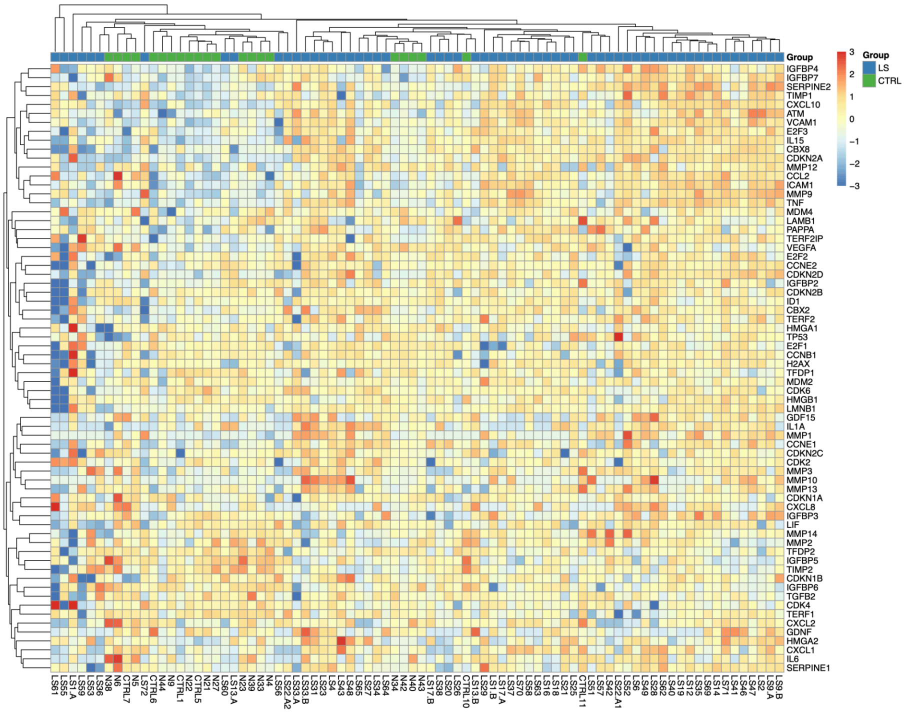


**Legend**. LS samples (n = 60) are shown in blue and HC (n = 22) in green. Two clusters were identified. The first containing 2 LS samples and the second with all 22 HC samples and 58 LS samples. LS, localized scleroderma; HC, healthy controls.

**Supplementary Figure 10.** Correlation analysis of senescence and inflammation signals in different inflammatory skin diseases.

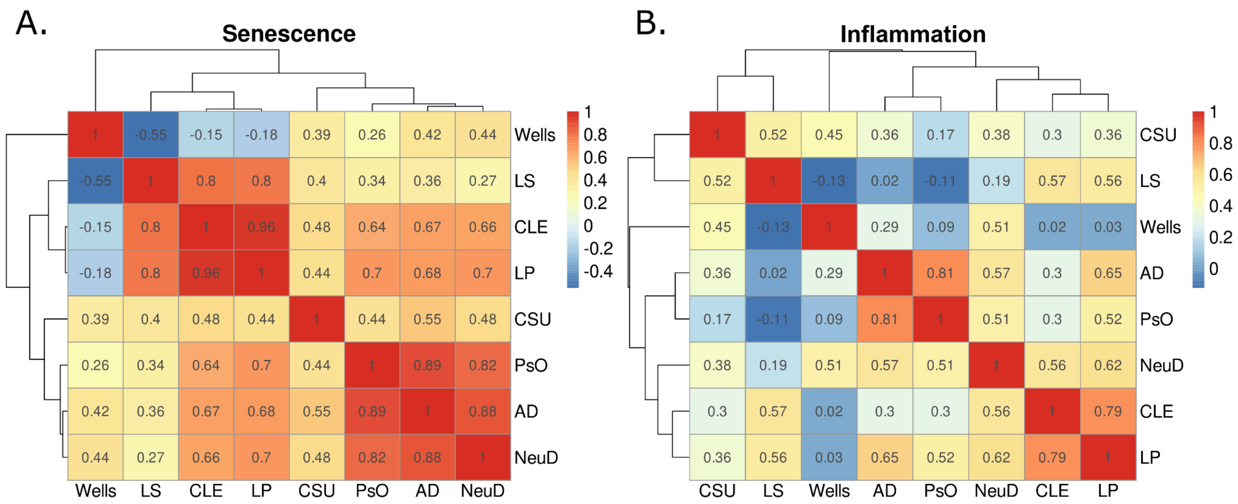


**Legend:** Correlation analysis of log-fold changes of senescence (A) and inflammation (B) gene sets with respect to healthy controls in multiple inflammatory skin diseases. Log-fold changes from CSU are as reported in Abboud et al. and all other diseases are as reported in Seremet et al.

### Supplementary Tables

**Supplementary Table 1.** Summary of Differentially Expressed Genes by Group.

| **Group *vs.* HC** | **Upregulated** | **Downregulated** | **Total** |
|---|---|---|---|
| **LS** | 488 | 216 | 704 |
| **Circumscribed LS** | 919 | 326 | 1245 |
| **Linear LS** | 1 | 0 | 1 |
| **Generalized LS** | 52 | 44 | 96 |
| **Active LS** | 607 | 247 | 854 |

#### Supplementary Materials

| | | | |
|---|---|---|---|
| **Inflammatory LS** | 755 | 255 | 1010 |
| **Fibrotic LS** | 103 | 82 | 185 |
| **Fibrotic *vs.* Inflammatory LS** | 4 | 21 | 25 |

**Supplemental Table 2.** Top 50 Upregulated Genes in LS *vs.* HC.

| Gene | Protein Encoded | logFC | Q-Value* | Functional Annotation** |
|---|---|---|---|---|
| *PLA2G2D* | Phospholipase A2 group IID | 6.688672643 | 0.007110983 | CD4-positive, CD25-positive, alpha-beta regulatory T cell differentiation, regulation of acute inflammatory response to antigenic stimulus, phospholipid metabolic process, inflammatory response, lipid catabolic process, negative regulation of T cell proliferation, phosphatidylethanolamine metabolic process, phosphatidylcholine metabolic process, phosphatidylglycerol metabolic process, and arachidonate secretion. |
| *CXCL11* | C-X-C motif chemokine ligand 11 | 6.245366036 | 0.003249031 | Chemotaxis, inflammatory response, signal transduction, adenylate cyclase-activating G protein-coupled receptor signaling pathway, cell-cell signaling, T cell chemotaxis, neutrophil chemotaxis, killing of cells of another organism, regulation of cell population proliferation, positive regulation of release of sequestered calcium ion into cytosol, antimicrobial humoral immune response mediated by antimicrobial peptide, chemokine-mediated signaling pathway, and cellular response to lipopolysaccharide. |
| *CCL3L1* | C-C motif chemokine ligand 3 like 3 | 6.142613845 | 0.005226239 | Inflammatory response, immune response, negative regulation of cell population proliferation, positive regulation of cell migration, eosinophil chemotaxis, cell chemotaxis, antimicrobial humoral immune response mediated by antimicrobial peptide, chemokine-mediated signaling pathway. |
| *ADAMDEC1* | ADAM like decysin 1 | 6.056222636 | 0.04070914 | Proteolysis, immune response, negative regulation of cell adhesion, and integrin-mediated signaling pathway. |
| *LAG3* | Lymphocyte activating 3 | 6.033139341 | $4.12 \times 10^{-5}$ | Adaptive immune response, plasmacytoid dendritic cell activation, cell surface receptor signaling pathway, negative regulation of interleukin-2 production, T cell activation, natural killer cell mediated cytotoxicity, innate immune response, negative regulation of regulatory T cell differentiation, positive regulation of natural killer cell mediated cytotoxicity, and regulation of immune response. |
| *FCRL3* | Fc receptor like 3 | 5.924085197 | 0.022750992 | Negative regulation of immunoglobulin production, immune response, cell surface receptor signaling pathway, single fertilization, positive regulation of B cell proliferation, regulation of toll-like receptor 9 signaling pathway, positive regulation of MAPK cascade, regulation of B cell differentiation, negative regulation of B cell receptor signaling pathway, regulation of B cell activation, and regulation of calcium ion import. |
| *GZMA* | Granzyme A | 5.871764965 | 0.023707833 | Apoptotic process, immune response, response to bacterium, killing of cells of another organism, negative regulation of endodeoxyribonuclease activity, positive regulation of apoptotic process, negative regulation of DNA binding, negative regulation of oxidoreductase activity, proteolysis involved in protein catabolic process, protein maturation, pyroptotic inflammatory response, granzyme-mediated programmed cell death signaling pathway, and cytotoxic T cell pyroptotic cell death. |
| *TTC24* | Tetratricopeptide repeat domain 24 | 5.755995659 | 0.001391623 | Unknown function. |
| *GZMK* | Granzyme K | 5.747502956 | 0.018256256 | Proteolysis, protein maturation, and granzyme-mediated programmed cell death signaling pathway. |
| *GZMB* | Granzyme B | 5.716554412 | 0.041048393 | Proteolysis, apoptotic process, negative regulation of translation, killing of cells of another organism, natural killer cell mediated cytotoxicity, proteolysis involved in protein catabolic process, protein maturation, pyroptotic inflammatory response, granzyme-mediated programmed cell death signaling pathway, and positive regulation of establishment of protein localization to mitochondrion. |
| *FCRL6* | Fc receptor like 6 | 5.694178167 | 0.002093133 | Immune response and cell surface receptor signaling pathway. |
| *IDO1* | Indoleamine 2,3-dioxygenase 1 | 5.683257857 | 0.018256256 | Immune system process, positive regulation of T cell tolerance induction, positive regulation of chronic inflammatory response, positive regulation of type 2 immune response, tryptophan catabolic process, inflammatory response, female pregnancy, tryptophan catabolic process to kynurenine, quinolinate biosynthetic process, response to |

### Supplementary Materials

| | | | | |
|---|---|---|---|---|
| | | | | lipopolysaccharide, negative regulation of interleukin-10 production, positive regulation of interleukin-12 production, multicellular organismal response to stress, kynurenic acid biosynthetic process, 'de novo' NAD biosynthetic process from tryptophan, swimming behavior, T cell proliferation, negative regulation of T cell proliferation, positive regulation of apoptotic process, negative regulation of activated T cell proliferation, negative regulation of T cell apoptotic process, and positive regulation of T cell apoptotic process. |
| *SLC12A3* | Solute carrier family 12 member 3 | 5.530573307 | 0.00958932 | Response to dietary excess, monoatomic ion transport, sodium ion transport, cell volume homeostasis, signal transduction, sodium ion transmembrane transport, chloride ion homeostasis, potassium ion homeostasis, sodium ion homeostasis, renal sodium ion absorption, chloride transmembrane transport, response to aldosterone, and potassium ion import across plasma membrane. |
| *TRAJ6* | T cell receptor alpha joining 6 | 5.463055029 | 0.003365506 | Unknown function. |
| *PACSIN1* | Protein kinase C and casein kinase substrate in neurons 1 | 5.372147688 | 0.000489485 | Cytoskeleton organization, actin filament organization, regulation of endocytosis, negative regulation of endocytosis, synaptic vesicle endocytosis, neuron projection morphogenesis, protein localization to membrane, protein localization to plasma membrane, plasma membrane tubulation, and positive regulation of dendrite development. |
| *CRTAM* | Cytotoxic and regulatory T cell molecule | 5.355148574 | 0.00936682 | Establishment of T cell polarity, positive regulation of cytokine production, adaptive immune response, detection of tumor cell, positive regulation of natural killer cell mediated cytotoxicity directed against tumor cell target, heterophilic cell-cell adhesion via plasma membrane cell adhesion molecules, cell recognition, positive regulation of type II interferon production, regulation of T cell differentiation, positive regulation of natural killer cell mediated cytotoxicity, negative regulation of activated T cell proliferation, regulation of T cell activation, detection of stimulus, lymphocyte migration into lymphoid organs, regulation of CD8-positive, and alpha-beta T cell activation. |
| *PLA1A* | Phospholipase A1 member A | 5.337813327 | 0.00890008 | Lipid metabolic process, phosphatidylserine metabolic process, lipid catabolic process, phosphatidylserine acyl-chain remodeling. |
| *UBASH3A* | Ubiquitin associated and SH3 domain containing A | 5.3009699 | 0.0272082 | Regulation of cytokine production, negative regulation of T cell receptor signaling pathway. |
| *C11orf42* | Chromosome 11 open reading frame 42 | 5.175823303 | 0.000108786 | Unknown function. |
| *TIGIT* | T cell immunoreceptor with Ig and ITIM domains | 5.145687245 | 0.009228263 | Negative regulation of interleukin-12 production, positive regulation of interleukin-10 production, negative regulation of natural killer cell mediated cytotoxicity, and negative regulation of T cell activation. |
| *TDO2* | Tryptophan 2,3-dioxygenase | 5.096672656 | 0.001839534 | Tryptophan metabolic process, tryptophan catabolic process to kynurenine, tryptophan catabolic process to acetyl-CoA, protein homotetramerization, and response to nitroglycerin. |
| *FHAD1* | Forkhead associated phosphopeptide binding domain 1 | 5.050017219 | 0.012345216 | Unknown function. |
| *IL4I1* | Interleukin 4 induced 1 | 5.033357726 | 0.013579386 | Adaptive immune response, regulation of adaptive immune response, negative regulation of T cell mediated immune response to tumor cell, L-phenylalanine catabolic process, tryptophan catabolic process, tyrosine catabolic process, amino acid catabolic process, tryptophan catabolic process to indole-3-acetate, negative regulation of T cell proliferation, regulation of B cell differentiation, positive regulation of regulatory T cell differentiation, positive regulation of transcription by RNA polymerase II, and negative regulation of T cell activation. |
| *CETP* | Cholesteryl ester transfer protein | 4.990702239 | 0.002109718 | Triglyceride metabolic process, lipid transport, cholesterol metabolic process, negative regulation of macrophage derived foam cell differentiation, regulation of cholesterol efflux, phospholipid transport, cholesterol transport, positive regulation of cholesterol transport, triglyceride transport, very-low-density lipoprotein particle remodeling, low-density lipoprotein particle remodeling, high-density lipoprotein particle remodeling, cholesterol homeostasis, reverse cholesterol transport, phosphatidylcholine metabolic process, lipid homeostasis, phospholipid homeostasis, triglyceride homeostasis, and positive regulation of phospholipid transport. |

#### Supplementary Materials

| | | | | |
|---|---|---|---|---|
| *NUGGC* | Nuclear GTPase, germinal center associated | 4.940816841 | 0.015344689 | Regulation of nuclear cell cycle DNA replication, negative regulation of apoptotic process, and cellular response to lipopolysaccharide. |
| *MYBPC2* | Myosin binding protein C2 | 4.859517253 | 0.006283998 | Cell adhesion and sarcomere organization. |
| *CNGB1* | Cyclic nucleotide gated channel subunit beta 1 | 4.858911412 | 0.014387163 | Retina homeostasis, monoatomic cation transport, potassium ion transport, sodium ion transport, calcium ion transport, visual perception, phototransduction, sensory perception of smell, positive regulation of gene expression, olfactory nerve maturation, protein localization to organelle, monoatomic ion transmembrane transport, sodium ion transmembrane transport, photoreceptor cell outer segment organization, photoreceptor cell maintenance, detection of light stimulus involved in visual perception, detection of chemical stimulus involved in sensory perception of smell, regulation of cytosolic calcium ion concentration, membrane depolarization, calcium ion transmembrane transport, monoatomic cation transmembrane transport, ion channel modulating, G protein-coupled receptor signaling pathway, and response to odorant. |
| *TRAJ27* | T cell receptor alpha joining 27 | 4.825429153 | 0.005434958 | Unknown function. |
| *IGHJ5* | Immunoglobulin heavy joining 5 | 4.78715156 | 0.013621039 | Unknown function. |
| *GZMH* | Granzyme H | 4.758083195 | 0.024103686 | Proteolysis, apoptotic process, killing of cells of another organism and protein maturation. |
| *SLCO5A1* | Solute carrier organic anion transporter family member 5A1 | 4.745371139 | 0.007940488 | Sodium-independent organic anion transport and transmembrane transport. |
| *CD27* | CD27 molecule | 4.647829102 | 0.043411078 | Chromatin remodeling, protein phosphorylation, intracellular zinc ion homeostasis, cell surface receptor protein tyrosine kinase signaling pathway, response to xenobiotic stimulus, peptidyl-tyrosine phosphorylation, hemopoiesis, platelet activation, T cell differentiation, T cell costimulation, positive regulation of heterotypic cell-cell adhesion, intracellular signal transduction, peptidyl-tyrosine autophosphorylation, Fc-gamma receptor signaling pathway, T cell activation, innate immune response, T cell receptor signaling pathway, B cell receptor signaling pathway, positive regulation of T cell receptor signaling pathway, positive regulation of T cell activation, leukocyte migration, release of sequestered calcium ion into cytosol, regulation of lymphocyte activation, CD27 signaling pathway, positive regulation of leukocyte cell-cell adhesion, positive regulation of cysteine-type endopeptidase activity, positive regulation of intrinsic apoptotic signaling pathway. |
| *MT-TF* | Mitochondrially encoded tRNA-Phe (UUU/C) | 4.643258439 | 0.01915914 | Unknown function. |
| *SI* | Sucrase-isomaltas e | 4.622431912 | 0.021193089 | Sucrose catabolic process, polysaccharide digestion. |
| *CD80* | CD80 molecule | 4.589830369 | 0.02333148 | Negative regulation of T cell mediated immunity, immune response, cell surface receptor signaling pathway, positive regulation of signal transduction, T cell costimulation, positive regulation of granulocyte macrophage colony-stimulating factor production, positive regulation of interleukin-2 production, intracellular signal transduction, positive regulation of T cell proliferation, T cell activation, negative regulation of T cell proliferation, positive regulation of T-helper 1 cell differentiation, positive regulation of DNA-templated transcription, symbiont entry into host cell, positive regulation of peptidyl-tyrosine phosphorylation, cellular response to lipopolysaccharide. |
| *LTB* | Lymphotoxin beta | 4.543480816 | 0.016756836 | Immune response, signal transduction, cell surface receptor signaling pathway, cell-cell signaling, gene expression, positive regulation of interleukin-12 production, positive regulation of canonical NF-kappaB signal transduction, skin development, lymph node development, and positive regulation of extrinsic apoptotic signaling pathway. |
| *FCGR1CP* | Fc gamma receptor Ic, pseudogene | 4.524153212 | 0.025053267 | Unknown function. |
| *CERKL* | Ceramide kinase like | 4.509781931 | 0.01318947 | Sphingolipid metabolic process, sphingolipid biosynthetic process, negative regulation of apoptotic process. |

#### Supplementary Materials

| | | | | |
|---|---|---|---|---|
| *SIRPG* | Signal regulatory protein gamma | 4.483421939 | 0.018283214 | Cell adhesion, cell-cell signaling, positive regulation of cell population proliferation, negative regulation of cell population proliferation, positive regulation of cell-cell adhesion, intracellular signal transduction, positive regulation of phagocytosis, and positive regulation of T cell activation. |
| *RSAD2* | Radical S-adenosyl methionine domain containing 2 | 4.446929375 | 0.006283998 | Response to virus, positive regulation of toll-like receptor 7 signaling pathway, positive regulation of toll-like receptor 9 signaling pathway, CD4-positive, alpha-beta T cell activation, CD4-positive, alpha-beta T cell differentiation, negative regulation of viral genome replication, innate immune response, negative regulation of protein secretion, positive regulation of immune response, defense response to virus, and positive regulation of T-helper 2 cell cytokine production. |
| *TRBV29-1* | T cell receptor beta variable 29-1 | 4.444754567 | 0.011852885 | Adaptive immune response, and cell surface receptor signaling pathway. |
| *CD38* | CD38 molecule | 4.439671087 | 0.021248028 | Response to hypoxia, signal transduction, positive regulation of cytosolic calcium ion concentration, female pregnancy, response to xenobiotic stimulus, negative regulation of neuron projection development, artery smooth muscle contraction, NAD metabolic process, positive regulation of cell growth, positive regulation of B cell proliferation, positive regulation of insulin secretion, response to estradiol, response to retinoic acid, response to progesterone, response to hydroperoxide, B cell proliferation, negative regulation of apoptotic process, negative regulation of bone resorption, negative regulation of DNA-templated transcription, positive regulation of DNA-templated transcription, positive regulation of vasoconstriction, B cell receptor signaling pathway, long-term synaptic depression, response to interleukin-1, and apoptotic signaling pathway. |
| *HSF5* | Heat shock transcription factor 5 | 4.377535487 | 0.014806683 | Regulation of transcription by RNA polymerase II, male meiotic nuclear division, spermatogenesis, cell differentiation, negative regulation of DNA-templated transcription, and positive regulation of DNA-templated transcription. |
| *TRAJ35* | T cell receptor alpha joining 35 | 4.364782256 | 0.022463611 | Unknown function. |
| *MZB1* | Marginal zone B and B1 cell specific protein | 4.359642883 | 0.027124816 | Positive regulation of immunoglobulin production, apoptotic process, positive regulation of cell population proliferation, regulation of B cell proliferation, integrin activation, regulation of cell population proliferation, and regulation of insulin receptor signaling pathway. |
| *GABRA2* | Gamma-aminobutyric acid type A receptor subunit alpha2 | 4.354552325 | 0.00529443 | Signal transduction, gamma-aminobutyric acid signaling pathway, nervous system process, synaptic transmission, GABAergic, regulation of postsynaptic membrane potential, regulation of presynaptic membrane potential, chloride transmembrane transport, and inhibitory synapse assembly. |
| *NKG7* | Natural killer cell granule protein 7 | 4.347761812 | 0.028245544 | Natural killer cell mediated cytotoxicity directed against tumor cell target, inflammatory response, killing of cells of another organism, CD4-positive, alpha-beta T cell activation, natural killer cell mediated cytotoxicity, defense response to protozoan, natural killer cell degranulation, positive regulation of inflammatory response, and granzyme-mediated programmed cell death signaling pathway. |
| *LCK* | LCK proto-oncogene, Src family tyrosine kinase | 4.345902917 | 0.046340803 | chromatin remodeling, protein phosphorylation, intracellular zinc ion homeostasis, cell surface receptor protein tyrosine kinase signaling pathway, response to xenobiotic stimulus, peptidyl-tyrosine phosphorylation, hemopoiesis, platelet activation, T cell differentiation, T cell costimulation, positive regulation of heterotypic cell-cell adhesion, intracellular signal transduction, peptidyl-tyrosine autophosphorylation, Fc-gamma receptor signaling pathway, T cell activation, innate immune response, T cell receptor signaling pathway, B cell receptor signaling pathway, positive regulation of T cell receptor signaling pathway, positive regulation of T cell activation, leukocyte migration, release of sequestered calcium ion into cytosol, regulation of lymphocyte activation, CD27 signaling pathway, positive regulation of leukocyte cell-cell adhesion, positive regulation of cysteine-type endopeptidase activity, and positive regulation of intrinsic apoptotic signaling pathway. |
| *TMPRSS15* | Transmembrane serine protease 15 | 4.325058959 | 0.027590791 | Proteolysis. |
| *ERN2* | Endoplasmic reticulum to nucleus signaling 2 | 4.272834206 | 0.02240727 | Chromatin remodeling, mRNA processing, protein phosphorylation, regulation of gene expression, rRNA catabolic process, apoptotic chromosome condensation, endoplasmic reticulum unfolded protein response, response to endoplasmic reticulum stress, IRE1-mediated unfolded protein response, negative regulation of DNA-templated transcription, intrinsic apoptotic signaling pathway in |

| | | | | |
|---|---|---|---|---|
| | | | | response to endoplasmic reticulum stress, and regulation of primary metabolic process. |

*P-values were corrected by multiple hypothesis testing to give the Q-value by the Benjamini-Hochberg method. **Functional Annotation is adapted from Database for Annotation, Visualization and Integrated Discovery (DAVID).

### Supplementary Materials

**Supplemental Table 3.** Top 50 Downregulated Genes in LS *vs.* HC.

| Gene | Protein Encoded | logFC | Q-Value* | Functional Annotation** |
|---|---|---|---|---|
| *WIF1* | WNT inhibitory factor 1] | -4.30775 | 0.002233 | Blood vessel development, signal transduction, determination of left/right symmetry, heart development, anterior/posterior pattern specification, Wnt signaling pathway, negative regulation of Wnt signaling pathway, nodal signaling pathway, and positive regulation of fat cell differentiation. |
| *FOSB* | FosB proto-oncogene, AP-1 transcription factor subunit | -4.27236 | 0.000139 | Negative regulation of transcription by RNA polymerase II, response to amphetamine, regulation of transcription by RNA polymerase II, transcription by RNA polymerase II, female pregnancy, response to xenobiotic stimulus, response to mechanical stimulus, response to progesterone, cellular response to hormone stimulus, response to morphine, response to ethanol, positive regulation of transcription by RNA polymerase II, behavioral response to cocaine, response to corticosterone, response to cAMP, and cellular response to calcium ion. |
| *FOS* | Fos proto-oncogene, AP-1 transcription factor subunit | -3.88019 | $3.97 \times 10^{-6}$ | Conditioned taste aversion, neural retina development, regulation of transcription by RNA polymerase II, transcription by RNA polymerase II, inflammatory response, transforming growth factor beta receptor signaling pathway, nervous system development, female pregnancy, response to xenobiotic stimulus, response to light stimulus, response to gravity, response to toxic substance, response to activity, skeletal muscle cell proliferation, cerebral cortex development, neuron differentiation, osteoclast differentiation, response to lipopolysaccharide, response to progesterone, response to insulin, cellular response to zinc ion starvation, cellular response to reactive oxygen species, response to immobilization stress, skeletal muscle cell differentiation, response to muscle stretch, response to ethanol, positive regulation of osteoclast differentiation, positive regulation of DNA-templated transcription, positive regulation of transcription by RNA polymerase II, response to corticosterone, myoblast proliferation, response to cAMP, SMAD protein signal transduction, cellular response to cadmium ion, cellular response to calcium ion, cellular response to tumor necrosis factor, cellular response to epidermal growth factor stimulus, cellular response to parathyroid hormone stimulus, cellular response to hypoxia, cellular response to transforming growth factor beta stimulus, medium-term memory, integrated stress response signaling, positive regulation of miRNA transcription, mononuclear cell differentiation, cellular response to phorbol 13-acetate 12-myristate, and cellular response to prolactin. |
| *SERTM1* | Serine rich and transmembrane domain containing 1 | -3.85252 | 0.013101 | Membrane and intracellular membrane-bounded organelle. |
| *CES1P1* | Carboxylesterase 1 pseudogene 1 | -3.779 | 0.011209 | Anatomical structure morphogenesis, and lipid catabolic process. |
| *SMIM24* | Small integral membrane protein 24 | -3.69327 | 0.018827 | Biological process. |
| *BMP3* | Bone morphogenetic protein 3 | -3.639 | 0.003995 | Skeletal system development, osteoblast differentiation, signal transduction, cell-cell signaling, negative regulation of ossification, and cartilage development. |
| *SERPINA5* | Serpin family A member 5 | -3.60132 | 0.001341 | Lipid transport, spermatogenesis, fusion of sperm to egg plasma membrane involved in single fertilization, negative regulation of hydrolase activity. |
| *AHSP* | Alpha hemoglobin stabilizing protein | -3.56512 | 0.037068 | Protein folding, hemoglobin metabolic process, hemopoiesis, erythrocyte differentiation, protein stabilization. |
| *CLDN11* | Claudin 11 | -3.52415 | 0.000694 | Cell adhesion, spermatogenesis, axon ensheathment, calcium-independent cell-cell adhesion via plasma membrane cell-adhesion molecules, bicellular tight junction assembly, and tight junction assembly, |
| *LY6K* | Lymphocyte antigen 6 family member K | -3.51171 | 0.00076 | Binding of sperm to zona pellucida. |
| *GREM2* | Gremlin 2, DAN family BMP antagonist | -3.38141 | $7.34 \times 10^{-5}$ | Animal organ morphogenesis, embryonic body morphogenesis, cytokine-mediated signaling pathway, sequestering of BMP from receptor via BMP binding, determination of dorsal identity, and regulation of cytokine activity. |

#### Supplementary Materials

| | | | | |
|---|---|---|---|---|
| *ENSG00000 167046* | Novel transcript, antisense to SLCO4A1 | -3.35299 | 0.01608 | *Pseudogene*. |
| *LCN2* | Lipocalin 2 | -3.31469 | 0.039033 | Apoptotic process, siderophore transport, defense response to bacterium, innate immune response, positive regulation of cold-induced thermogenesis. |
| *MATN4* | Matrilin 4 | -3.30803 | 0.027031 | Extracellular matrix organization. |
| *ENSG00000 262202* | Novel transcript | -3.29652 | 0.005071 | *Pseudogene*. |
| *HAS1* | Hyaluronan synthase 1 | -3.27285 | 0.007937 | Polysaccharide biosynthetic process, glycosaminoglycan biosynthetic process, cell adhesion, negative regulation of fibroblast migration, hyaluronan biosynthetic process, cellular response to platelet-derived growth factor stimulus, and extracellular matrix assembly. |
| *ENSG00000 286555* | Novel transcript, antisense to ARAP1 | -3.22237 | 0.008371 | *Pseudogene*. |
| *WBP11P3* | WBP11 pseudogene 3 | -3.21958 | 0.006321 | Unknown function. |
| *EDN3* | Endothelin 3 | -3.17405 | 0.042376 | Neural crest cell migration, positive regulation of leukocyte chemotaxis, regulation of systemic arterial blood pressure by endothelin, intracellular calcium ion homeostasis, signal transduction, cell surface receptor signaling pathway, cell-cell signaling, axon guidance, blood circulation, cell population proliferation, positive regulation of cell population proliferation, positive regulation of heart rate, regulation of gene expression, intracellular magnesium ion homeostasis, vein smooth muscle contraction, regulation of vasoconstriction, peptide hormone secretion, melanocyte differentiation, neutrophil chemotaxis, vasoconstriction, positive regulation of MAP kinase activity, positive regulation of cell differentiation, positive regulation of mitotic nuclear division, positive regulation of smooth muscle contraction, positive regulation of hormone secretion, regulation of developmental pigmentation, axon extension, establishment of localization in cell, potassium ion transmembrane transport, and positive regulation of potassium ion transmembrane transport. |
| *HBA2* | Hemoglobin subunit alpha 2 | -3.1303 | 0.03787 | Carbon dioxide transport, oxygen transport, nitric oxide transport, response to hydrogen peroxide, hydrogen peroxide catabolic process, and cellular oxidant detoxification. |
| *PBOV1* | Prostate and breast cancer overexpressed 1 | -3.10098 | 0.01608 | Nucleoplasm, cytosol, and centriolar satellite. |
| *EGILA* | EGFR interacting lncRNA | -3.05486 | 0.007663 | Unknown function. |
| *LYVE1* | Lymphatic vessel endothelial hyaluronan receptor 1 | -3.05249 | 0.004795 | Positive regulation of cellular extravasation, receptor-mediated endocytosis, cell adhesion, cell-matrix adhesion, response to wounding, anatomical structure morphogenesis, and hyaluronan catabolic process. |
| *TNMD* | Tenomodulin | -3.03149 | 0.025152 | Endothelial cell morphogenesis, negative regulation of endothelial cell proliferation, and negative regulation of angiogenesis. |
| *ENSG00000 212383* | | -3.01226 | 0.008371 | *Pseudogene*. |
| *ENSG00000 222007* | Novel transcript, antisense to AGAP1 | -2.99522 | 0.023409 | *Pseudogene*. |
| *VEGFD* | Vascular endothelial growth factor D | -2.96672 | 0.048087 | Response to hypoxia, positive regulation of protein phosphorylation, positive regulation of endothelial cell proliferation, sprouting angiogenesis, positive regulation of cell population proliferation, response to bacterium, vascular endothelial growth factor signaling pathway, positive regulation of angiogenesis, vascular endothelial growth factor receptor signaling pathway, fibroblast proliferation, positive chemotaxis, induction of positive chemotaxis, positive regulation of cell division, positive regulation of mast cell chemotaxis, dopaminergic neuron differentiation. |
| *EGR1* | Early growth response 1 | -2.92043 | $1.63 \times 10^{-5}$ | Negative regulation of transcription by RNA polymerase II, response to hypoxia, response to ischemia, regulation of transcription by RNA polymerase II, response to glucose, positive regulation of gene expression, T cell differentiation, BMP signaling pathway, positive regulation of chemokine production, positive regulation of interleukin-1 beta production, response to insulin, circadian regulation of gene |

| | | | | |
|---|---|---|---|---|
| | | | | expression, regulation of protein sumoylation, skeletal muscle cell differentiation, regulation of neuron apoptotic process, positive regulation of gene expression via chromosomal CpG island demethylation, estrous cycle, locomotor rhythm, positive regulation of DNA-templated transcription, positive regulation of transcription by RNA polymerase II, positive regulation of hormone biosynthetic process, rhythmic process, circadian temperature homeostasis, interleukin-1-mediated signaling pathway, cellular response to gamma radiation, cellular response to heparin, cellular response to mycophenolic acid, glomerular mesangial cell proliferation, positive regulation of glomerular metanephric mesangial cell proliferation, negative regulation of canonical Wnt signaling pathway, cellular response to interleukin-8, positive regulation of post-translational protein modification, positive regulation of miRNA transcription, and regulation of progesterone biosynthetic process. |
| *TCEAL7* | Transcription elongation factor A like 7 | -2.92014 | 0.021956 | Negative regulation of NF-kappaB transcription factor activity, and negative regulation of DNA-templated transcription, |
| *MT1G* | Metallothionein 1G | -2.90844 | 0.009113 | Intracellular zinc ion homeostasis, detoxification of copper ion, monocyte differentiation, cellular response to vascular endothelial growth factor stimulus, monocyte activation, negative regulation of growth, cellular response to cadmium ion, cellular response to copper ion, cellular response to zinc ion. |
| *ASS1P9* | Argininosuccinate synthetase 1 pseudogene 9 | -2.90158 | 0.009367 | Unknown function. |
| *DUSP1* | Dual specificity phosphatase 1 | -2.89487 | $3.97 \times 10^{-6}$ | Endoderm formation, chromatin remodeling, regulation of transcription by RNA polymerase II, negative regulation of cell adhesion, signal transduction, negative regulation of cell population proliferation, peptidyl-tyrosine dephosphorylation, peptidyl-threonine dephosphorylation, negative regulation of MAP kinase activity, negative regulation of MAPK cascade, negative regulation of meiotic cell cycle, peptidyl-serine dephosphorylation, negative regulation of monocyte chemotaxis, regulation of mitotic cell cycle spindle assembly checkpoint, negative regulation of p38MAPK cascade, and cellular response to chemokine. |
| *ATP5MC1P6* | ATP synthase membrane subunit c locus 1 pseudogene 6 | -2.85877 | 0.013579 | Unknown function. |
| *ENSG00000 250770* | Tetraspanin 11 (TSPAN11) pseudogene | -2.857 | 0.021193 | *Pseudogene*. |
| *RPLP1P6* | Ribosomal protein lateral stalk subunit P1 pseudogene 6 | -2.82165 | 0.001902 | Unknown function. |
| *OR51B6* | Olfactory receptor family 51 subfamily B member 6 | -2.81222 | 0.041359 | G protein-coupled receptor signaling pathway, sensory perception of smell, and detection of chemical stimulus involved in sensory perception of smell. |
| *PCOLCE2* | Procollagen C-endopeptidase enhancer 2 | -2.80883 | 0.000241 | Cellular response to leukemia inhibitory factor. |
| *BEX5* | Brain expressed X-linked 5 | -2.79737 | 0.024104 | Signal transduction. |
| *ENSG00000 249593* | Novel transcript | -2.71517 | 0.022554 | *Pseudogene*. |
| *MAB21L1* | mab-21 like 1 | -2.71066 | 0.041788 | Eye development, cell population proliferation, positive regulation of cell population proliferation, anatomical structure morphogenesis, and camera-type eye development. |
| *PCP4L1* | Purkinje cell protein 4 like 1 | -2.6771 | 0.043133 | Unknown function. |
| *OR5P1P* | Olfactory receptor family 5 subfamily P member 1 pseudogene | -2.67189 | 0.035258 | G protein-coupled receptor signaling pathway. |
| *ENSG00000 278001* | Family with sequence | -2.65331 | 0.027467 | *Pseudogene*. |

| | | | | |
|---|---|---|---|---|
| | similarity 27, member D1 (FAM27D1) pseudogene | | | |
| *BUB3P1* | BUB3 mitotic checkpoint protein pseudogene 1 | -2.64831 | 0.021243 | Unknown function. |
| *EAF1-AS1* | EAF1 antisense RNA 1 | -2.61058 | 0.041048 | Unknown function. |
| *CYP4Z2P* | Cytochrome P450 family 4 subfamily Z member 2, pseudogene | -2.54727 | 0.021709 | Membrane. |
| *ENSG00000 287458* | Novel transcript, antisense to FKBP5 | -2.53469 | 0.028476 | *Pseudogene*. |
| *ENSG00000 224172* | Myelin protein zero-like 1 (MPZL1) pseudogene | -2.51767 | 0.049966 | *Pseudogene*. |
| *NR4A1* | Nuclear receptor subfamily 4 group A member 1 | -2.50294 | 0.00311 | Positive regulation of endothelial cell proliferation, response to amphetamine, cell migration involved in sprouting angiogenesis, regulation of transcription by RNA polymerase II, transcription by RNA polymerase II, apoptotic process, inflammatory response, signal transduction, neurotransmitter secretion involved in regulation of skeletal muscle contraction, intracellular receptor signaling pathway, detection of lipopolysaccharide, endothelial cell chemotaxis, skeletal muscle cell differentiation, cellular response to vascular endothelial growth factor stimulus, positive regulation of apoptotic process, cellular response to fibroblast growth factor stimulus, fat cell differentiation, negative regulation of cell cycle, positive regulation of transcription by RNA polymerase II, response to electrical stimulus, regulation of type B pancreatic cell proliferation, cellular response to corticotropin-releasing hormone stimulus, and non-canonical inflammasome complex assembly. |

*P-values were corrected by multiple hypothesis testing to give the Q-value by the Benjamini-Hochberg method. **Functional Annotation** is adapted from Database for Annotation, Visualization and Integrated Discovery (DAVID).

**Supplemental Table 4.** Top 50 Upregulated Genes in Active LS *vs.* HC.

| Gen | Protein Encoded | logFC | Q-Value* | Functional Annotation** |
|---|---|---|---|---|
| *PLA2G2D* | Phospholipase A2 group IID | 7.145342 | 0.006775 | CD4-positive, CD25-positive, alpha-beta regulatory T cell differentiation, regulation of acute inflammatory response to antigenic stimulus, phospholipid metabolic process, inflammatory response, lipid catabolic process, negative regulation of T cell proliferation, phosphatidylethanolamine metabolic process, phosphatidylcholine metabolic process, phosphatidylglycerol metabolic process, and arachidonate secretion. |
| *CCL3L1* | C-C motif chemokine ligand 3 like 3 | 6.673807 | 0.003566 | Inflammatory response, immune response, negative regulation of cell population proliferation, positive regulation of cell migration, eosinophil chemotaxis, cell chemotaxis, antimicrobial humoral immune response mediated by antimicrobial peptide, and chemokine-mediated signaling pathway. |
| *CXCL11* | C-X-C motif chemokine ligand 11 | 6.444596 | 0.004036 | Chemotaxis, inflammatory response, signal transduction, adenylate cyclase-activating G protein-coupled receptor signaling pathway, cell-cell signaling, T cell chemotaxis, neutrophil chemotaxis, killing of cells of another organism, regulation of cell population proliferation, positive regulation of release of sequestered calcium ion into cytosol, antimicrobial humoral immune response mediated by antimicrobial peptide, chemokine-mediated signaling pathway, and cellular response to lipopolysaccharide. |
| *LAG3* | Lymphocyte activating 3 | 6.210131 | 0.000345 | Adaptive immune response, plasmacytoid dendritic cell activation, cell surface receptor signaling pathway, negative regulation of interleukin-2 production, T cell activation, natural killer cell mediated cytotoxicity, innate immune response, negative regulation of regulatory T cell |

#### Supplementary Materials

| | | | | |
|---|---|---|---|---|
| | | | | differentiation, positive regulation of natural killer cell mediated cytotoxicity, and regulation of immune response. |
| *FCRL3* | Fc receptor like 3 | 6.153138 | 0.030514 | Negative regulation of immunoglobulin production, immune response, cell surface receptor signaling pathway, single fertilization, positive regulation of B cell proliferation, regulation of toll-like receptor 9 signaling pathway, positive regulation of MAPK cascade, regulation of B cell differentiation, negative regulation of B cell receptor signaling pathway, regulation of B cell activation, and regulation of calcium ion import. |
| *GZMA* | Granzyme A | 6.138617 | 0.02761 | Apoptotic process, immune response, response to bacterium, killing of cells of another organism, negative regulation of endodeoxyribonuclease activity, positive regulation of apoptotic process, negative regulation of DNA binding, negative regulation of oxidoreductase activity, proteolysis involved in protein catabolic process, protein maturation, pyroptotic inflammatory response, granzyme-mediated programmed cell death signaling pathway, and cytotoxic T cell pyroptotic cell death. |
| *TTC24* | Tetratricopeptide repeat domain 24 | 6.096883 | 0.002165 | Unknown function. |
| *GZMB* | Granzyme B | 5.988126 | 0.013801 | Proteolysis, apoptotic process, negative regulation of translation, killing of cells of another organism, natural killer cell mediated cytotoxicity, proteolysis involved in protein catabolic process, protein maturation, pyroptotic inflammatory response, granzyme-mediated programmed cell death signaling pathway, and positive regulation of establishment of protein localization to mitochondrion. |
| *IDO1* | Indoleamine 2,3-dioxygenase 1 | 5.941746 | 0.009571 | Immune system process, positive regulation of T cell tolerance induction, positive regulation of chronic inflammatory response, positive regulation of type 2 immune response, tryptophan catabolic process, inflammatory response, female pregnancy, tryptophan catabolic process to kynurenine, quinolinate biosynthetic process, response to lipopolysaccharide, negative regulation of interleukin-10 production, positive regulation of interleukin-12 production, multicellular organismal response to stress, kynurenic acid biosynthetic process, 'de novo' NAD biosynthetic process from tryptophan, swimming behavior, T cell proliferation, negative regulation of T cell proliferation, positive regulation of apoptotic process, negative regulation of activated T cell proliferation, negative regulation of T cell apoptotic process, and positive regulation of T cell apoptotic process. |
| *GZMK* | Granzyme K | 5.937387 | 0.021246 | Proteolysis, protein maturation, granzyme-mediated programmed cell death signaling pathway. |
| *FCRL6* | Fc receptor like 6 | 5.888325 | 0.001391 | Immune response and cell surface receptor signaling pathway. |
| *CRTAM* | Cytotoxic and regulatory T cell molecule | 5.753232 | 0.003914 | Establishment of T cell polarity, positive regulation of cytokine production, adaptive immune response, detection of tumor cell, positive regulation of natural killer cell mediated cytotoxicity directed against tumor cell target, heterophilic cell-cell adhesion via plasma membrane cell adhesion molecules, cell recognition, positive regulation of type II interferon production, regulation of T cell differentiation, positive regulation of natural killer cell mediated cytotoxicity, negative regulation of activated T cell proliferation, regulation of T cell activation, detection of stimulus, lymphocyte migration into lymphoid organs, regulation of CD8-positive, alpha-beta T cell activation. |
| *TRAJ6* | T cell receptor alpha joining 6 | 5.720692 | 0.005306 | Unknown function. |
| *SLC12A3* | Solute carrier family 12 member 3 | 5.665861 | 0.006592 | Response to dietary excess, monoatomic ion transport, sodium ion transport, cell volume homeostasis, signal transduction, sodium ion transmembrane transport, chloride ion homeostasis, potassium ion homeostasis, sodium ion homeostasis, renal sodium ion absorption, chloride transmembrane transport, response to aldosterone, potassium ion import across plasma membrane. |
| *PACSIN1* | Protein kinase C and casein kinase substrate in neurons 1 | 5.54518 | 0.000723 | Cytoskeleton organization, actin filament organization, regulation of endocytosis, negative regulation of endocytosis, synaptic vesicle endocytosis, neuron projection morphogenesis, protein localization to membrane, protein localization to plasma membrane, plasma membrane tubulation, and positive regulation of dendrite development. |
| *CNGB1* | Cyclic nucleotide gated channel subunit beta 1 | 5.509974 | 0.003937 | Retina homeostasis, monoatomic cation transport, potassium ion transport, sodium ion transport, calcium ion transport, visual perception, phototransduction, sensory perception of smell, positive regulation of gene expression, olfactory nerve maturation, protein localization to organelle, monoatomic ion transmembrane transport, sodium |

| | | | | |
|---|---|---|---|---|
| | | | | ion transmembrane transport, photoreceptor cell outer segment organization, photoreceptor cell maintenance, detection of light stimulus involved in visual perception, detection of chemical stimulus involved in sensory perception of smell, regulation of cytosolic calcium ion concentration, membrane depolarization, calcium ion transmembrane transport, monoatomic cation transmembrane transport, ion channel modulating, G protein-coupled receptor signaling pathway, and response to odorant. |
| *TIGIT* | T cell immunoreceptor with Ig and ITIM domains | 5.475679 | 0.007181 | Negative regulation of interleukin-12 production, positive regulation of interleukin-10 production, negative regulation of natural killer cell mediated cytotoxicity, negative regulation of T cell activation. |
| *PLA1A* | Phospholipase A1 member A | 5.446666 | 0.014155 | Lipid metabolic process, phosphatidylserine metabolic process, lipid catabolic process, and phosphatidylserine acyl-chain remodeling. |
| *TDO2* | Tryptophan 2,3-dioxygenase | 5.442905 | 0.00123 | Tryptophan metabolic process, tryptophan catabolic process to kynurenine, tryptophan catabolic process to acetyl-CoA, protein homotetramerization, and response to nitroglycerin. |
| *UBASH3A* | Ubiquitin associated and SH3 domain containing A | 5.38737 | 0.038179 | Regulation of cytokine production, and negative regulation of T cell receptor signaling pathway. |
| *C11orf42* | Chromosome 11 open reading frame 42 | 5.339356 | 0.000768 | Unknown function. |
| *NUGGC* | Nuclear GTPase, germinal center associated | 5.244099 | 0.010634 | Regulation of nuclear cell cycle DNA replication, negative regulation of apoptotic process, and cellular response to lipopolysaccharide. |
| *IL4I1* | Interleukin 4 induced 1 | 5.228244 | 0.010467 | Adaptive immune response, regulation of adaptive immune response, negative regulation of T cell mediated immune response to tumor cell, L-phenylalanine catabolic process, tryptophan catabolic process, tyrosine catabolic process, amino acid catabolic process, tryptophan catabolic process to indole-3-acetate, negative regulation of T cell proliferation, regulation of B cell differentiation, positive regulation of regulatory T cell differentiation, positive regulation of transcription by RNA polymerase II, and negative regulation of T cell activation. |
| *FHAD1* | Forkhead associated phosphopeptide binding domain 1 | 5.217995 | 0.010634 | Unknown function. |
| *MYBPC2* | Myosin binding protein C2 | 5.163321 | 0.006322 | Cell adhesion and sarcomere organization. |
| *CETP* | Cholesteryl ester transfer protein | 5.109605 | 0.002301 | Triglyceride metabolic process, lipid transport, cholesterol metabolic process, negative regulation of macrophage derived foam cell differentiation, regulation of cholesterol efflux, phospholipid transport, cholesterol transport, positive regulation of cholesterol transport, triglyceride transport, very-low-density lipoprotein particle remodeling, low-density lipoprotein particle remodeling, high-density lipoprotein particle remodeling, cholesterol homeostasis, reverse cholesterol transport, phosphatidylcholine metabolic process, lipid homeostasis, phospholipid homeostasis, triglyceride homeostasis, positive regulation of phospholipid transport. |
| *SLCO5A1* | Solute carrier organic anion transporter family member 5A1 | 5.049784 | 0.004744 | Sodium-independent organic anion transport, and transmembrane transport. |
| *GZMH* | Granzyme H | 5.048751 | 0.027121 | Proteolysis, apoptotic process, killing of cells of another organism, and protein maturation. |
| *MT-TF* | Mitochondrially encoded tRNA-Phe (UUU/C) | 5.018391 | 0.02272 | Unknown function. |
| *SI* | Sucrase-isomaltase | 4.967545 | 0.016193 | Sucrose catabolic process and polysaccharide digestion. |
| *TRAJ27* | T cell receptor alpha joining 27 | 4.882745 | 0.009168 | Unknown function. |

#### Supplementary Materials

| | | | | |
|---|---|---|---|---|
| *FCGR1CP* | Fc gamma receptor Ic, pseudogene | 4.871066 | 0.016733 | Unknown function. |
| *IGHJ5* | Immunoglobulin heavy joining 5 | 4.846011 | 0.027666 | Unknown function. |
| *CD80* | CD80 molecule | 4.772745 | 0.011539 | Negative regulation of T cell mediated immunity, immune response, cell surface receptor signaling pathway, positive regulation of signal transduction, T cell costimulation, positive regulation of granulocyte macrophage colony-stimulating factor production, positive regulation of interleukin-2 production, intracellular signal transduction, positive regulation of T cell proliferation, T cell activation, negative regulation of T cell proliferation, positive regulation of T-helper 1 cell differentiation, positive regulation of DNA-templated transcription, symbiont entry into host cell, positive regulation of peptidyl-tyrosine phosphorylation, cellular response to lipopolysaccharide. |
| *CERKL* | Ceramide kinase like | 4.758256 | 0.018244 | Sphingolipid metabolic process, sphingolipid biosynthetic process, negative regulation of apoptotic process. |
| *TMPRSS15* | Transmembrane serine protease 15 | 4.697581 | 0.031376 | Proteolysis. |
| *XIRP1* | Xin actin binding repeat containing 1 | 4.641701 | 0.015676 | Actin filament organization, protein localization, myoblast differentiation involved in skeletal muscle regeneration, satellite cell activation involved in skeletal muscle regeneration, negative regulation of protein binding. |
| *CD38* | CD38 molecule | 4.623221 | 0.017265 | Response to hypoxia, signal transduction, positive regulation of cytosolic calcium ion concentration, female pregnancy, response to xenobiotic stimulus, negative regulation of neuron projection development, artery smooth muscle contraction, NAD metabolic process, positive regulation of cell growth, positive regulation of B cell proliferation, positive regulation of insulin secretion, response to estradiol, response to retinoic acid, response to progesterone, response to hydroperoxide, B cell proliferation, negative regulation of apoptotic process, negative regulation of bone resorption, negative regulation of DNA-templated transcription, positive regulation of DNA-templated transcription, positive regulation of vasoconstriction, B cell receptor signaling pathway, long-term synaptic depression, response to interleukin-1, and apoptotic signaling pathway. |
| *HSF5* | Heat shock transcription factor 5 | 4.603031 | 0.009236 | Regulation of transcription by RNA polymerase II, male meiotic nuclear division, spermatogenesis, cell differentiation, negative regulation of DNA-templated transcription, and positive regulation of DNA-templated transcription. |
| *SIRPG* | Signal regulatory protein gamma | 4.599195 | 0.016733 | Cell adhesion, cell-cell signaling, positive regulation of cell population proliferation, negative regulation of cell population proliferation, positive regulation of cell-cell adhesion, intracellular signal transduction, positive regulation of phagocytosis, and positive regulation of T cell activation. |
| *CCR5* | C-C motif chemokine receptor 5 | 4.581131 | 0.026418 | MAPK cascade, dendritic cell chemotaxis, calcium ion transport, apoptotic process, chemotaxis, inflammatory response, immune response, cellular defense response, cell surface receptor signaling pathway, G protein-coupled receptor signaling pathway, positive regulation of cytosolic calcium ion concentration, cell-cell signaling, release of sequestered calcium ion into cytosol by sarcoplasmic reticulum, calcium-mediated signaling, signaling, symbiont entry into host cell, cell chemotaxis, chemokine-mediated signaling pathway, response to cholesterol, cellular response to lipopolysaccharide, and negative regulation of macrophage apoptotic process. |
| *MUC17* | Mucin 17, cell surface associated | 4.580817 | 0.015661 | Cellular homeostasis. |
| *LTB* | Lymphotoxin beta | 4.570669 | 0.023857 | Immune response, signal transduction, cell surface receptor signaling pathway, cell-cell signaling, gene expression, positive regulation of interleukin-12 production, positive regulation of canonical NF-kappaB signal transduction, skin development, lymph node development, and positive regulation of extrinsic apoptotic signaling pathway. |
| *NKG7* | Natural killer cell granule protein 7 | 4.544554 | 0.02271 | Natural killer cell mediated cytotoxicity directed against tumor cell target, inflammatory response, killing of cells of another organism, CD4-positive, alpha-beta T cell activation, natural killer cell mediated cytotoxicity, defense response to protozoan, natural killer cell degranulation, positive regulation of inflammatory response, and granzyme-mediated programmed cell death signaling pathway. |
| *RSAD2* | Radical S-adenosyl | 4.535936 | 0.000402 | Response to virus, positive regulation of toll-like receptor 7 signaling pathway, positive regulation of toll-like receptor 9 signaling |

| | | | | |
|---|---|---|---|---|
| | methionine domain containing 2 | | | pathway, CD4-positive, alpha-beta T cell activation, CD4-positive, alpha-beta T cell differentiation, negative regulation of viral genome replication, innate immune response, negative regulation of protein secretion, positive regulation of immune response, defense response to virus, and positive regulation of T-helper 2 cell cytokine production. |
| *ERN2* | Endoplasmic reticulum to nucleus signaling 2 | 4.534816 | 0.017815 | Chromatin remodeling, mRNA processing, protein phosphorylation, regulation of gene expression, rRNA catabolic process, apoptotic chromosome condensation, endoplasmic reticulum unfolded protein response, response to endoplasmic reticulum stress, IRE1-mediated unfolded protein response, negative regulation of DNA-templated transcription, intrinsic apoptotic signaling pathway in response to endoplasmic reticulum stress, and regulation of primary metabolic process. |
| *TRAJ35* | T cell receptor alpha joining 35 (non-functional) | 4.521478 | 0.032457 | Unknown function. |
| *LCK* | LCK proto-oncogene, Src family tyrosine kinase | 4.512847 | 0.032199 | Chromatin remodeling, protein phosphorylation, intracellular zinc ion homeostasis, cell surface receptor protein tyrosine kinase signaling pathway, response to xenobiotic stimulus, peptidyl-tyrosine phosphorylation, hemopoiesis, platelet activation, T cell differentiation, T cell costimulation, positive regulation of heterotypic cell-cell adhesion, intracellular signal transduction, peptidyl-tyrosine autophosphorylation, Fc-gamma receptor signaling pathway, T cell activation, innate immune response, T cell receptor signaling pathway, B cell receptor signaling pathway, positive regulation of T cell receptor signaling pathway, positive regulation of T cell activation, leukocyte migration, release of sequestered calcium ion into cytosol, regulation of lymphocyte activation, CD27 signaling pathway, positive regulation of leukocyte cell-cell adhesion, positive regulation of cysteine-type endopeptidase activity, and positive regulation of intrinsic apoptotic signaling pathway. |
| *TMPRSS3* | Transmembrane serine protease 3 | 4.498638 | 0.018763 | Proteolysis, intracellular sodium ion homeostasis, sensory perception of sound. |
| *GABRA2* | Gamma-aminobutyric acid type A receptor subunit alpha2 | 4.492456 | 0.002318 | Signal transduction, gamma-aminobutyric acid signaling pathway, nervous system process, synaptic transmission, GABAergic, regulation of postsynaptic membrane potential, regulation of presynaptic membrane potential, chloride transmembrane transport, and inhibitory synapse assembly. |

*P-values were corrected by multiple hypothesis testing to give the Q-value by the Benjamini-Hochberg method. **Functional Annotation** is adapted from Database for Annotation, Visualization and Integrated Discovery (DAVID).

#### Supplementary Materials

**Supplemental Table 5.** Top 50 Downregulated Genes in Active LS *vs.* HC.

| Gen | Protein Encoded | logFC | Q-Value* | Functional Annotation** |
|---|---|---|---|---|
| *FOSB* | FosB proto-oncogene, AP-1 transcription factor subunit | -4.3914 | 0.002024 | Negative regulation of transcription by RNA polymerase II, response to amphetamine, regulation of transcription by RNA polymerase II, transcription by RNA polymerase II, female pregnancy, response to xenobiotic stimulus, response to mechanical stimulus, response to progesterone, cellular response to hormone stimulus, response to morphine, response to ethanol, positive regulation of transcription by RNA polymerase II, behavioral response to cocaine, response to corticosterone, response to cAMP, and cellular response to calcium ion. |
| *WIF1* | WNT inhibitory factor 1 | -4.22511 | 0.003037 | Blood vessel development, signal transduction, determination of left/right symmetry, heart development, anterior/posterior pattern specification, Wnt signaling pathway, negative regulation of Wnt signaling pathway, nodal signaling pathway, and positive regulation of fat cell differentiation. |
| *FOS* | Fos proto-oncogene, AP-1 transcription factor subunit | -3.9625 | 0.000138 | Conditioned taste aversion, neural retina development, regulation of transcription by RNA polymerase II, transcription by RNA polymerase II, inflammatory response, transforming growth factor beta receptor signaling pathway, nervous system development, female pregnancy, response to xenobiotic stimulus, response to light stimulus, response to gravity, response to toxic substance, response to activity, skeletal muscle cell proliferation, cerebral cortex development, neuron differentiation, osteoclast differentiation, response to lipopolysaccharide, response to progesterone, response to insulin, cellular response to zinc ion starvation, cellular response to reactive oxygen species, response to immobilization stress, skeletal muscle cell differentiation, response to muscle stretch, response to ethanol, positive regulation of osteoclast differentiation, positive regulation of DNA-templated transcription, positive regulation of transcription by RNA polymerase II, response to corticosterone, myoblast proliferation, response to cAMP, SMAD protein signal transduction, cellular response to cadmium ion, cellular response to calcium ion, cellular response to tumor necrosis factor, cellular response to epidermal growth factor stimulus, cellular response to parathyroid hormone stimulus, cellular response to hypoxia, cellular response to transforming growth factor beta stimulus, medium-term memory, integrated stress response signaling, positive regulation of miRNA transcription, mononuclear cell differentiation, cellular response to phorbol 13-acetate 12-myristate, and cellular response to prolactin. |
| *CES1P1* | Carboxylesterase 1 pseudogene 1 | -3.91195 | 0.009364 | Anatomical structure morphogenesis and lipid catabolic process. |
| *SERTM1* | Serine rich and transmembrane domain containing 1 | -3.85911 | 0.005306 | Membrane and intracellular membrane-bounded organelle. |
| *SMIM24* | Small integral membrane protein 24 | -3.74675 | 0.008594 | Biological process. |
| *CLDN11* | Claudin 11 | -3.71947 | 0.000402 | Cell adhesion, spermatogenesis, axon ensheathment, calcium-independent cell-cell adhesion via plasma membrane cell-adhesion molecules, bicellular tight junction assembly, and tight junction assembly. |
| *LCN2* | Lipocalin 2 | -3.60159 | 0.024515 | Apoptotic process, siderophore transport, defense response to bacterium, innate immune response, and positive regulation of cold-induced thermogenesis. |
| *AHSP* | Alpha hemoglobin stabilizing protein | -3.50024 | 0.03125 | Protein folding, hemoglobin metabolic process, hemopoiesis, erythrocyte differentiation, and protein stabilization. |
| *LY6K* | Lymphocyte antigen 6 family member K | -3.45932 | 0.000851 | Binding of sperm to zona pellucida. |
| *BMP3* | Bone morphogenetic protein 3 | -3.44182 | 0.006902 | Skeletal system development, osteoblast differentiation, signal transduction, cell-cell signaling, negative regulation of ossification, and cartilage development. |
| *GREM2* | Gremlin 2, DAN family BMP antagonist | -3.42979 | 0.000593 | Animal organ morphogenesis, embryonic body morphogenesis, cytokine-mediated signaling pathway, sequestering of BMP from receptor via BMP binding, determination of dorsal identity, regulation of cytokine activity, |

#### Supplementary Materials

| | | | | |
|---|---|---|---|---|
| *SERPINA5* | Serpin family A member 5 | -3.38738 | 0.005712 | Lipid transport, spermatogenesis, fusion of sperm to egg plasma membrane involved in single fertilization, and negative regulation of hydrolase activity. |
| *ENSG00000 167046* | Novel transcript, antisense to SLCO4A1 | -3.38305 | 0.009924 | *Pseudogene*. |
| *HBA2* | Hemoglobin subunit alpha 2 | -3.29759 | 0.045225 | Carbon dioxide transport, oxygen transport, nitric oxide transport, response to hydrogen peroxide, hydrogen peroxide catabolic process, and cellular oxidant detoxification. |
| *ENSG00000 286555* | Novel transcript, antisense to ARAP1 | -3.28985 | 0.003937 | *Pseudogene*. |
| *SNORD53* | Small nucleolar RNA, C/D box 53 | -3.27392 | 0.03499 | RNA processing. |
| *EDN3* | Endothelin 3 | -3.20579 | 0.049111 | Neural crest cell migration, positive regulation of leukocyte chemotaxis, regulation of systemic arterial blood pressure by endothelin, intracellular calcium ion homeostasis, signal transduction, cell surface receptor signaling pathway, cell-cell signaling, axon guidance, blood circulation, cell population proliferation, positive regulation of cell population proliferation, positive regulation of heart rate, regulation of gene expression, intracellular magnesium ion homeostasis, vein smooth muscle contraction, regulation of vasoconstriction, peptide hormone secretion, melanocyte differentiation, neutrophil chemotaxis, vasoconstriction, positive regulation of MAP kinase activity, positive regulation of cell differentiation, positive regulation of mitotic nuclear division, positive regulation of smooth muscle contraction, positive regulation of hormone secretion, regulation of developmental pigmentation, axon extension, establishment of localization in cell, potassium ion transmembrane transport, and positive regulation of potassium ion transmembrane transport. |
| *WBP11P3* | WBP11 pseudogene 3 | -3.12671 | 0.006902 | Unknown function. |
| *EGR1* | Early growth response 1 | -3.05651 | $1.93 \times 10^{-5}$ | Negative regulation of transcription by RNA polymerase II, response to hypoxia, response to ischemia, regulation of transcription by RNA polymerase II, response to glucose, positive regulation of gene expression, T cell differentiation, BMP signaling pathway, positive regulation of chemokine production, positive regulation of interleukin-1 beta production, response to insulin, circadian regulation of gene expression, regulation of protein sumoylation, skeletal muscle cell differentiation, regulation of neuron apoptotic process, positive regulation of gene expression via chromosomal CpG island demethylation, estrous cycle, locomotor rhythm, positive regulation of DNA-templated transcription, positive regulation of transcription by RNA polymerase II, positive regulation of hormone biosynthetic process, rhythmic process, circadian temperature homeostasis, interleukin-1-mediated signaling pathway, cellular response to gamma radiation, cellular response to heparin, cellular response to mycophenolic acid, glomerular mesangial cell proliferation, positive regulation of glomerular metanephric mesangial cell proliferation, negative regulation of canonical Wnt signaling pathway, cellular response to interleukin-8, positive regulation of post-translational protein modification, positive regulation of miRNA transcription, and regulation of progesterone biosynthetic process. |
| *MIR361* | MicroRNA 361 | -3.04203 | 0.036792 | Negative regulation of angiogenesis, negative regulation of transforming growth factor beta receptor signaling pathway, regulation of interleukin-1 beta production, regulation of interleukin-6 production, regulation of tumor necrosis factor production, miRNA-mediated post-transcriptional gene silencing, negative regulation of apoptotic process, regulation of inflammatory response, negative regulation of cell migration involved in sprouting angiogenesis, negative regulation of amyloid-beta formation, negative regulation of blood vessel endothelial cell proliferation involved in sprouting angiogenesis, and negative regulation of vascular endothelial growth factor production. |
| *HAS1* | Hyaluronan synthase 1 | -3.03759 | 0.01056 | Polysaccharide biosynthetic process, glycosaminoglycan biosynthetic process, cell adhesion, negative regulation of fibroblast migration, hyaluronan biosynthetic process, cellular response to platelet-derived growth factor stimulus, and extracellular matrix assembly. |
| *ENSG00000 222007* | Novel transcript, antisense to AGAP1 | -3.02051 | 0.019931 | *Pseudogene*. |

#### Supplementary Materials

| | | | | |
|---|---|---|---|---|
| *OR51B6* | Olfactory receptor family 51 subfamily B member 6 | -3.00885 | 0.017146 | G protein-coupled receptor signaling pathway, sensory perception of smell, and detection of chemical stimulus involved in sensory perception of smell. |
| *ENSG00000 262202* | Novel transcript | -3.00695 | 0.01056 | *Pseudogene*. |
| *MAB21L1* | mab-21 like 1 | -2.98474 | 0.032888 | Eye development, cell population proliferation, positive regulation of cell population proliferation, anatomical structure morphogenesis, and camera-type eye development. |
| *PBOV1* | Prostate and breast cancer overexpressed 1 | -2.96997 | 0.01914 | Nucleoplasm, cytosol, and centriolar satellite. |
| *DUSP1* | Dual specificity phosphatase 1 | -2.96878 | 0.000125 | Endoderm formation, chromatin remodeling, regulation of transcription by RNA polymerase II, negative regulation of cell adhesion, signal transduction, negative regulation of cell population proliferation, peptidyl-tyrosine dephosphorylation, peptidyl-threonine dephosphorylation, negative regulation of MAP kinase activity, negative regulation of MAPK cascade, negative regulation of meiotic cell cycle, peptidyl-serine dephosphorylation, negative regulation of monocyte chemotaxis, regulation of mitotic cell cycle spindle assembly checkpoint, negative regulation of p38MAPK cascade, and cellular response to chemokine. |
| *ASS1P9* | Argininosuccinate synthetase 1 pseudogene 9 | -2.95782 | 0.014701 | Unknown function. |
| *MT1G* | Metallothionein 1G | -2.95523 | 0.008875 | Intracellular zinc ion homeostasis, detoxification of copper ion, monocyte differentiation, cellular response to vascular endothelial growth factor stimulus, monocyte activation, negative regulation of growth, cellular response to cadmium ion, cellular response to copper ion, cellular response to zinc ion. |
| *TCEAL7* | Transcription elongation factor A like 7 | -2.94704 | 0.018689 | Negative regulation of NF-kappaB transcription factor activity, and negative regulation of DNA-templated transcription. |
| *LYVE1* | Lymphatic vessel endothelial hyaluronan receptor 1 | -2.94366 | 0.007827 | Positive regulation of cellular extravasation, receptor-mediated endocytosis, cell adhesion, cell-matrix adhesion, response to wounding, anatomical structure morphogenesis, and hyaluronan catabolic process. |
| *ATP5MC1P6* | ATP synthase membrane subunit c locus 1 pseudogene 6 | -2.90845 | 0.016334 | Unknown function. |
| *PCOLCE2* | Procollagen C-endopeptidase enhancer 2 | -2.87064 | 0.00123 | Cellular response to leukemia inhibitory factor. |
| *TNMD* | Tenomodulin | -2.85149 | 0.045738 | Endothelial cell morphogenesis, negative regulation of endothelial cell proliferation, negative regulation of angiogenesis. |
| *PCP4L1* | Purkinje cell protein 4 like 1 | -2.76761 | 0.040722 | Unknown function. |
| *OR5P1P* | Olfactory receptor family 5 subfamily P member 1 pseudogene | -2.76632 | 0.027143 | G protein-coupled receptor signaling pathway. |
| *BEX5* | Brain expressed X-linked 5 | -2.7446 | 0.027914 | Signal transduction. |
| *RPLP1P6* | Ribosomal protein lateral stalk subunit P1 pseudogene 6 | -2.71382 | 0.003332 | Skeletal system development, osteoblast differentiation, signal transduction, cell-cell signaling, negative regulation of ossification, cartilage development. |
| *CYP4Z2P* | Cytochrome P450 family 4 subfamily Z member 2, pseudogene | -2.67929 | 0.013056 | Membrane. |
| *ENSG00000 249593* | Novel transcript | -2.65124 | 0.038314 | *Pseudogene*. |
| *ENSG00000 257668* | Suppression of tumorigenicity | -2.63367 | 0.026429 | *Pseudogene*. |

| | | | | |
|---|---|---|---|---|
| | 13 (colon carcinoma) (Hsp70 interacting protein) (ST13) pseudogene | | | |
| *ENSG00000 278001* | Family with sequence similarity 27, member D1 (FAM27D1) pseudogene | -2.62499 | 0.029619 | *Pseudogene.* |
| *LINC00482* | Long intergenic non-protein coding RNA 482 | -2.62492 | 0.044789 | Unknown function. |
| *ENSG00000 250770* | Tetraspanin 11 (TSPAN11) pseudogene | -2.61514 | 0.037141 | *Pseudogene.* |
| *CES1* | Carboxylesterase 1 | -2.60977 | 0.001355 | Cholesterol biosynthetic process, cholesterol metabolic process, response to toxic substance, positive regulation of cholesterol efflux, negative regulation of cholesterol storage, lipid catabolic process, epithelial cell differentiation, cholesterol homeostasis, reverse cholesterol transport, medium-chain fatty acid metabolic process, regulation of bile acid biosynthetic process, cellular response to cholesterol, cellular response to low-density lipoprotein particle stimulus, cholesterol ester hydrolysis involved in cholesterol transport, positive regulation of cholesterol metabolic process, and regulation of bile acid secretion. |
| *NR4A1* | Nuclear receptor subfamily 4 group A member 1 | -2.5983 | 0.001008 | Positive regulation of endothelial cell proliferation, response to amphetamine, cell migration involved in sprouting angiogenesis, regulation of transcription by RNA polymerase II, transcription by RNA polymerase II, apoptotic process, inflammatory response, signal transduction, neurotransmitter secretion involved in regulation of skeletal muscle contraction, intracellular receptor signaling pathway, detection of lipopolysaccharide, endothelial cell chemotaxis, skeletal muscle cell differentiation, cellular response to vascular endothelial growth factor stimulus, positive regulation of apoptotic process, cellular response to fibroblast growth factor stimulus, fat cell differentiation, negative regulation of cell cycle, positive regulation of transcription by RNA polymerase II, response to electrical stimulus, regulation of type B pancreatic cell proliferation, cellular response to corticotropin-releasing hormone stimulus, and non-canonical inflammasome complex assembly. |
| *ENSG00000 287458* | Novel transcript, antisense to FKBP5 | -2.57631 | 0.044397 | *Pseudogene.* |
| *ENSG00000 275493* | Family with sequence similarity 27 member (FAM27D) pseudogene | -2.56022 | 0.026392 | *Pseudogene.* |
| *CYP4F26P* | Cytochrome P450 family 4 subfamily F member 26, pseudogene | -2.50984 | 0.027071 | Unknown function. |

*P-values were corrected by multiple hypothesis testing to give the Q-value by the Benjamini-Hochberg method. **Functional Annotation is adapted from Database for Annotation, Visualization and Integrated Discovery (DAVID).

**Supplemental Table 6.** Top 50 Upregulated Genes in Fibrotic LS *vs.* HC.

| Gen | Protein Encoded | logFC | Q-Value* | Functional Annotation** |
|---|---|---|---|---|
| *CXCL11* | C-X-C motif chemokine ligand 11 | 5.55661 | 0.04027 | Chemotaxis, inflammatory response, signal transduction, adenylate cyclase-activating G protein-coupled receptor signaling pathway, cell-cell signaling, T cell chemotaxis, neutrophil chemotaxis, killing of cells of another organism, regulation of cell population proliferation, positive regulation of release of sequestered calcium ion into cytosol, antimicrobial humoral immune response mediated by antimicrobial peptide, chemokine-mediated signaling pathway, cellular response to lipopolysaccharide. |

#### Supplementary Materials

| | | | | |
|---|---|---|---|---|
| *LAG3* | Lymphocyte activating 3 | 5.386998 | 0.00354 | Adaptive immune response, plasmacytoid dendritic cell activation, cell surface receptor signaling pathway, negative regulation of interleukin-2 production, T cell activation, natural killer cell mediated cytotoxicity, innate immune response, negative regulation of regulatory T cell differentiation, positive regulation of natural killer cell mediated cytotoxicity, regulation of immune response. |
| *FCRL6* | Fc receptor like 6 | 5.194828 | 0.012081 | Immune response, cell surface receptor signaling pathway. |
| *C11orf42* | Chromosome 11 open reading frame 42 | 5.102859 | 0.005442 | Unknown function. |
| *TTC24* | Tetratricopeptide repeat domain 24 | 4.991697 | 0.035392 | Unknown function. |
| *FHAD1* | Forkhead associated phosphopeptide binding domain 1 | 4.989522 | 0.04953 | Unknown function. |
| *MYBPC2* | Myosin binding protein C2 | 4.923845 | 0.033437 | Cell adhesion, sarcomere organization. |
| *PACSIN1* | Protein kinase C and casein kinase substrate in neurons 1 | 4.740086 | 0.013996 | Cytoskeleton organization, actin filament organization, regulation of endocytosis, negative regulation of endocytosis, synaptic vesicle endocytosis, neuron projection morphogenesis, protein localization to membrane, protein localization to plasma membrane, plasma membrane tubulation, and positive regulation of dendrite development. |
| *TDO2* | Tryptophan 2,3-dioxygenase | 4.717037 | 0.022475 | Tryptophan metabolic process, tryptophan catabolic process to kynurenine, tryptophan catabolic process to acetyl-CoA, protein homotetramerization, and response to nitroglycerin. |
| *TRAJ27* | T cell receptor alpha joining 27 | 4.571222 | 0.040689 | Unknown function. |
| *SLCO5A1* | Solute carrier organic anion transporter family member 5A1 | 4.553125 | 0.042818 | Sodium-independent organic anion transport, transmembrane transport. |
| *IL4I1* | Interleukin 4 induced 1 | 4.533697 | 0.039936 | Adaptive immune response, regulation of adaptive immune response, negative regulation of T cell mediated immune response to tumor cell, L-phenylalanine catabolic process, tryptophan catabolic process, tyrosine catabolic process, amino acid catabolic process, tryptophan catabolic process to indole-3-acetate, negative regulation of T cell proliferation, regulation of B cell differentiation, positive regulation of regulatory T cell differentiation, positive regulation of transcription by RNA polymerase II, negative regulation of T cell activation. |
| *GABRA2* | Gamma-aminobutyric acid type A receptor subunit alpha2 | 4.532455 | 0.006996 | Signal transduction, gamma-aminobutyric acid signaling pathway, nervous system process, synaptic transmission, GABAergic, regulation of postsynaptic membrane potential, regulation of presynaptic membrane potential, chloride transmembrane transport, inhibitory synapse assembly. |
| *CETP* | Cholesteryl ester transfer protein | 4.478423 | 0.017711 | Triglyceride metabolic process, lipid transport, cholesterol metabolic process, negative regulation of macrophage derived foam cell differentiation, regulation of cholesterol efflux, phospholipid transport, cholesterol transport, positive regulation of cholesterol transport, triglyceride transport, very-low-density lipoprotein particle remodeling, low-density lipoprotein particle remodeling, high-density lipoprotein particle remodeling, cholesterol homeostasis, reverse cholesterol transport, phosphatidylcholine metabolic process, lipid homeostasis, phospholipid homeostasis, triglyceride homeostasis, positive regulation of phospholipid transport. |
| *RSAD2* | Radical S-adenosyl methionine domain containing 2 | 4.275915 | 0.010203 | Response to virus, positive regulation of toll-like receptor 7 signaling pathway, positive regulation of toll-like receptor 9 signaling pathway, CD4-positive, alpha-beta T cell activation, CD4-positive, alpha-beta T cell differentiation, negative regulation of viral genome replication, innate immune response, negative regulation of protein secretion, positive regulation of immune response, defense response to virus, positive regulation of T-helper 2 cell cytokine production. |
| *FAM81A* | Family with sequence similarity 81 member A | 4.259548 | 0.016191 | Postsynaptic density, glutamatergic synapse, postsynaptic density, intracellular component. |
| *TLE6* | TLE family member 6, subcortical maternal complex member | 4.122797 | 0.014912 | Negative regulation of transcription by RNA polymerase II, regulation of DNA-templated transcription, actin filament organization, positive regulation of embryonic development, establishment of spindle localization, regulation of cell division, endoplasmic reticulum localization, mitochondrion localization, embryonic process involved in female pregnancy, negative regulation of canonical Wnt signaling pathway. |

#### Supplementary Materials

| | | | | |
|---|---|---|---|---|
| *MYCN* | MYCN proto-oncogene, bHLH transcription factor | 4.12176 | 0.036625 | Cartilage condensation, positive regulation of mesenchymal cell proliferation, regulation of transcription by RNA polymerase II, positive regulation of gene expression, negative regulation of gene expression, lung development, embryonic digit morphogenesis, positive regulation of programmed cell death, regulation of inner ear auditory receptor cell differentiation, positive regulation of DNA-templated transcription, positive regulation of transcription by RNA polymerase II, embryonic skeletal system morphogenesis, astrocyte differentiation, negative regulation of astrocyte differentiation, branching morphogenesis of an epithelial tube, epithelial cell proliferation, positive regulation of epithelial cell proliferation, autosome genomic imprinting, positive regulation of miRNA transcription, negative regulation of reactive oxygen species metabolic process. |
| *RIMS1* | Regulating synaptic membrane exocytosis 1 | 4.111956 | 0.042596 | Intracellular protein transport, exocytosis, visual perception, positive regulation of gene expression, synaptic vesicle exocytosis, synaptic vesicle docking, synaptic vesicle priming, calcium-ion regulated exocytosis, cell differentiation, regulation of membrane potential, regulated exocytosis, secretion, regulation of neurotransmitter secretion, regulation of synaptic plasticity, maintenance of presynaptic active zone structure, calcium ion-regulated exocytosis of neurotransmitter, positive regulation of synaptic transmission, acrosomal vesicle exocytosis, membrane fusion, protein-containing complex assembly, positive regulation of inhibitory postsynaptic potential, positive regulation of dendrite extension, regulation of synaptic vesicle exocytosis, and positive regulation of excitatory postsynaptic potential. |
| *REM1* | RRAD and GEM like GTPase 1 | 3.97167 | 0.005296 | Regulation of skeletal muscle contraction by calcium ion signaling and negative regulation of calcium ion transmembrane transport via high voltage-gated calcium channel. |
| *ENSG00000 174171* | Novel transcript, antisense to SPTBN5 | 3.911225 | 0.027722 | *Pseudogene*. |
| *DOCK8-AS1* | DOCK8 antisense RNA 1 | 3.850546 | 0.013941 | Unknown function. |
| *VWCE* | von Willebrand factor C and EGF domains | 3.817417 | 0.010203 | Cellular response to virus. |
| *CDKN2A* | Cyclin dependent kinase inhibitor 2A | 3.719878 | 0.033437 | Protein polyubiquitination, autophagy of mitochondrion, negative regulation of cell-matrix adhesion, rRNA processing, apoptotic process, Ras protein signal transduction, negative regulation of cell population proliferation, apoptotic mitochondrial changes, positive regulation of gene expression, protein sumoylation, negative regulation of cell growth, nuclear body organization, negative regulation of B cell proliferation, regulation of protein stability, protein destabilization, negative regulation of NF-kappaB transcription factor activity, negative regulation of immature T cell proliferation in thymus, positive regulation of protein sumoylation, positive regulation of smooth muscle cell apoptotic process, protein localization to nucleus, negative regulation of phosphorylation, positive regulation of apoptotic process, positive regulation of DNA damage response, signal transduction by p53 class mediator, negative regulation of cyclin-dependent protein serine/threonine kinase activity, negative regulation of DNA-templated transcription, positive regulation of DNA-templated transcription, positive regulation of transcription by RNA polymerase II, regulation of protein export from nucleus, somatic stem cell division, protein stabilization, negative regulation of ubiquitin-protein transferase activity, regulation of cell cycle, mitochondrial depolarization, protein K63-linked ubiquitination, cellular senescence, replicative senescence, apoptotic signaling pathway, positive regulation of protein localization to nucleus, positive regulation of signal transduction by p53 class mediator, regulation of apoptotic DNA fragmentation, protein localization to nucleolus, negative regulation of proteolysis involved in protein catabolic process, regulation of protein targeting to mitochondrion, negative regulation of ubiquitin protein ligase activity, amyloid fibril formation, regulation of G1/S transition of mitotic cell cycle, negative regulation of ubiquitin-dependent protein catabolic process, positive regulation of macrophage apoptotic process, negative regulation of protein neddylation. |
| *MYLK4* | Myosin light chain kinase family member 4 | 3.656177 | 0.036501 | Chromatin remodeling, signal transduction. |
| *RNF222* | Ring finger protein 222 | 3.602479 | 0.030393 | Membrane. |

#### Supplementary Materials

| | | | | |
|---|---|---|---|---|
| *SIGLEC15* | Sialic acid binding Ig like lectin 15 | 3.596505 | 0.023054 | Regulation of actin cytoskeleton organization, regulation of bone resorption, regulation of osteoclast development. |
| *ABHD17AP6* | ABHD17A pseudogene 6 | 3.560523 | 0.01218 | Unknown function. |
| *NTN3* | Netrin 3 | 3.491718 | 0.042596 | Regulation of transcription by RNA polymerase II, axon guidance. |
| *RPGRIP1* | RPGR interacting protein 1 | 3.474238 | 0.04637 | Visual perception, retinal rod cell development, neural precursor cell proliferation, non-motile cilium assembly. |
| *A1BG-AS1* | A1BG antisense RNA 1 | 3.468825 | 0.023054 | Unknown function. |
| *ENSG00000 235721* | Pseudogene similar to part of zinc finger protein 532 (ZNF532) | 3.449912 | 0.047215 | *Pseudogene.* |
| *STEAP1B* | STEAP family member 1B | 3.440973 | 0.047051 | Endosome, plasma membrane, membrane. |
| *GPR137C* | G protein-coupled receptor 137C | 3.347321 | 0.046808 | Positive regulation of TORC1 signaling. |
| *ENSG00000 224611* | TBPL1 pseudogene 1 | 3.334055 | 0.017792 | *Pseudogene.* |
| *SCO2* | Synthesis of cytochrome C oxidase 2 | 3.311892 | 0.040689 | Eye development, in utero embryonic development, muscle system process, intracellular copper ion homeostasis, response to activity, respiratory electron transport chain, mitochondrial cytochrome c oxidase assembly, |
| *RAB39A* | RAB39A, member RAS oncogene family | 3.259419 | 0.043949 | Autophagy, protein transport, vesicle-mediated transport, Rab protein signal transduction, phagosome acidification, phagosome-lysosome fusion. |
| *FAM78A* | Family with sequence similarity 78 member A | 3.228266 | 0.032464 | Unknown function. |
| *HCST* | Hematopoietic cell signal transducer | 3.12696 | 0.035269 | Protein phosphorylation, natural killer cell mediated cytotoxicity, regulation of immune response, positive regulation of phosphatidylinositol 3-kinase/protein kinase B signal transduction. |
| *GIPC3* | GIPC PDZ domain containing family member 3 | 3.089121 | 0.04234 | Unknown function. |
| *ENSG00000 236015* | Thioredoxin domain containing 1 (TXNDC1) pseudogene | 3.057558 | 0.006675 | *Pseudogene.* |
| *ASNS* | Asparagine synthetase (glutamine-hydrolyzing) | 3.039847 | 0.022342 | Asparagine biosynthetic process, glutamine metabolic process, cellular response to glucose starvation, negative regulation of apoptotic process, positive regulation of mitotic cell cycle, L-asparagine biosynthetic process. |
| *TOR4A* | Torsin family 4 member A [ | 3.034659 | 0.026879 | Extracellular region, nuclear envelope, endoplasmic reticulum lumen, membrane, platelet alpha granule lumen. |
| *PCDH17* | Protocadherin 17 | 2.909988 | 0.014367 | Cell adhesion, homophilic cell adhesion via plasma membrane adhesion molecules, adult behavior, negative regulation of synaptic transmission, synaptic membrane adhesion, presynaptic active zone assembly, regulation of synaptic vesicle clustering. |
| *SERF1B* | Small EDRK-rich factor 1B | 2.862248 | 0.049738 | Nervous system development, protein destabilization, amyloid fibril formation. |
| *IL15* | Interleukin 15 | 2.732843 | 0.01348 | Natural killer cell differentiation, natural killer cell proliferation, positive regulation of cytokine production, NK T cell proliferation, immune response, signal transduction, tyrosine phosphorylation of STAT protein, cell-cell signaling, positive regulation of cell population proliferation, macrophage differentiation, positive regulation of interleukin-17 production, positive regulation of natural killer cell proliferation, positive regulation of natural killer cell differentiation, positive regulation of tissue remodeling, interleukin-15-mediated signaling pathway, positive regulation of T cell proliferation, neutrophil activation, positive regulation of tyrosine phosphorylation of STAT protein, extrathymic T cell selection, regulation of T cell differentiation, cell maturation, lymph node development, regulation of defense response to virus by host, positive regulation of inflammatory response, positive regulation of peptidyl-tyrosine phosphorylation, positive regulation of phagocytosis, positive regulation of immune response, negative regulation of cold-induced thermogenesis, and positive regulation of protein O-linked glycosylation. |
| *HAVCR2* | Hepatitis A virus cellular receptor 2 | 2.678797 | 0.040689 | Adaptive immune response, macrophage activation involved in immune response, natural killer cell tolerance induction, regulation of tolerance |

| | | | | |
|---|---|---|---|---|
| | | | | induction dependent upon immune response, negative regulation of T-helper 1 type immune response, negative regulation of natural killer cell mediated cytotoxicity directed against tumor cell target, regulation of transcription by RNA polymerase II, inflammatory response, negative regulation of gene expression, negative regulation of myeloid dendritic cell activation, negative regulation of NF-kappaB transcription factor activity, negative regulation of interferon-alpha production, negative regulation of type II interferon production, negative regulation of interleukin-2 production, negative regulation of interleukin-3 production, negative regulation of interleukin-6 production, negative regulation of tumor necrosis factor production, positive regulation of chemokine production, positive regulation of type II interferon production, positive regulation of interleukin-1 production, positive regulation of interleukin-4 production, positive regulation of tumor necrosis factor production, negative regulation of natural killer cell activation, toll-like receptor 3 signaling pathway, toll-like receptor 7 signaling pathway, toll-like receptor 9 signaling pathway, positive regulation of T cell proliferation, negative regulation of T cell proliferation, positive regulation of macrophage activation, innate immune response, defense response to Gram-positive bacterium, maternal process involved in female pregnancy, positive regulation of ERK1 and ERK2 cascade, cellular response to lipopolysaccharide, negative regulation of granulocyte colony-stimulating factor production, negative regulation of defense response to bacterium, positive regulation of defense response to bacterium, positive regulation of non-canonical NF-kappaB signal transduction, negative regulation of immunological synapse formation, and negative regulation of T cell activation via T cell receptor contact with antigen bound to MHC molecule on antigen presenting cell. |
| *SCN1B* | Sodium voltage-gated channel beta subunit 1 | 2.668465 | 0.03107 | Sodium ion transport, cell adhesion, axon guidance, positive regulation of sodium ion transport, positive regulation of neuron projection development, neuronal action potential propagation, corticospinal neuron axon guidance, monoatomic ion transmembrane transport, sodium ion transmembrane transport, locomotion, membrane depolarization, cardiac muscle contraction, regulation of ventricular cardiac muscle cell membrane repolarization, regulation of atrial cardiac muscle cell membrane depolarization, cardiac conduction, cardiac muscle cell action potential involved in contraction, membrane depolarization during action potential, membrane depolarization during cardiac muscle cell action potential, membrane depolarization during Purkinje myocyte cell action potential, regulation of heart rate by cardiac conduction, regulation of sodium ion transmembrane transport, positive regulation of voltage-gated sodium channel activity. |
| *PEX12* | Peroxisomal biogenesis factor 12 | 2.644937 | 0.033291 | Protein polyubiquitination, protein monoubiquitination, protein quality control for misfolded or incompletely synthesized proteins, protein targeting to peroxisome, peroxisome organization, protein import into peroxisome matrix, protein import into peroxisome matrix, receptor recycling, cellular response to reactive oxygen species, proteasome-mediated ubiquitin-dependent protein catabolic process, protein import into peroxisome matrix, and substrate release. |
| *PBX4* | PBX homeobox 4 | 2.590101 | 0.030393 | Eye development, regulation of transcription by RNA polymerase II, brain development, animal organ morphogenesis, positive regulation of DNA-templated transcription, embryonic organ development, neuron development, |

#### Supplementary Materials

**Supplemental Table 7.** Top 50 Downregulated Genes in Fibrotic LS *vs.* HC.

| Gen | Protein Encoded | logFC | Q-Value* | Functional Annotation** |
|---|---|---|---|---|
| *WIF1* | WNT inhibitory factor 1 | -4.337 | 0.011922 | Blood vessel development, signal transduction, determination of left/right symmetry, heart development, anterior/posterior pattern specification, Wnt signaling pathway, negative regulation of Wnt signaling pathway, nodal signaling pathway, positive regulation of fat cell differentiation. |
| *FOSB* | FosB proto-oncogene, AP-1 transcription factor subunit | -3.90343 | 0.015168 | Negative regulation of transcription by RNA polymerase II, response to amphetamine, regulation of transcription by RNA polymerase II, transcription by RNA polymerase II, female pregnancy, response to xenobiotic stimulus, response to mechanical stimulus, response to progesterone, cellular response to hormone stimulus, response to morphine, response to ethanol, positive regulation of transcription by RNA polymerase II, behavioral response to cocaine, response to corticosterone, response to cAMP, cellular response to calcium ion. |
| *BMP3* | Bone morphogenetic protein 3 | -3.62818 | 0.014367 | Skeletal system development, osteoblast differentiation, signal transduction, cell-cell signaling, negative regulation of ossification, cartilage development. |
| *FOS* | Fos proto-oncogene, AP-1 transcription factor subunit | -3.61641 | 0.002633 | Oxidative Stress Induced Gene Expression Via Nrf2, B Cell Survival Pathway, BCR Signaling Pathway, Role of EGF Receptor Transactivation by GPCRs in Cardiac Hypertrophy, Pertussis toxin-insensitive CCR5 Signaling in Macrophage, Cadmium induces DNA synthesis and proliferation in macrophages, Repression of Pain Sensation by the Transcriptional Regulator DREAM, EGF Signaling Pathway, EPO Signaling Pathway, METS affect on Macrophage Differentiation, Fc Epsilon Receptor I Signaling in Mast Cells, Inhibition of Cellular Proliferation by Gleevec, Signaling Pathway from G-Protein Families, IGF-1 Signaling Pathway, IL 2 signaling pathway, IL-2 Receptor Beta Chain in T cell Activation, IL 3 signaling pathway, IL 6 signaling pathway, Insulin Signaling Pathway, Keratinocyte Differentiation, MAPKinase Signaling Pathway, Signaling of Hepatocyte Growth Factor Receptor, Nerve growth factor pathway (NGF), PDGF Signaling Pathway, Bone Remodelling, T Cell Receptor Signaling Pathway, Toll-Like Receptor Pathway, TPO Signaling Pathway, and TSP-1 Induced Apoptosis in Microvascular Endothelial Cell. |
| *PKMP2* | Pyruvate kinase M1/2 pseudogene 2 | -3.58904 | 0.033437 | Unknown function. |
| *ENSG00000 229569* | Novel transcript | -3.56887 | 0.032464 | *Pseudogene*. |
| *ENSG00000 262202* | Novel transcript | -3.53776 | 0.014367 | *Pseudogene*. |
| *ENSG00000 167046* | Novel transcript, antisense to SLCO4A1 | -3.4393 | 0.032464 | *Pseudogene*. |
| *SERTM1* | Serine rich and transmembrane domain containing 1 | -3.42522 | 0.037166 | Membrane and intracellular membrane-bounded organelle. |
| *SERPINA5* | Serpin family A member 5 | -3.36872 | 0.022342 | Lipid transport, spermatogenesis, fusion of sperm to egg plasma membrane involved in single fertilization, negative regulation of hydrolase activity. |
| *CLDN11* | Claudin 11 | -3.3459 | 0.007568 | Cell adhesion, spermatogenesis, axon ensheathment, calcium-independent cell-cell adhesion via plasma membrane cell-adhesion molecules, bicellular tight junction assembly, and tight junction assembly. |
| *GREM2* | Gremlin 2, DAN family BMP antagonist | -3.34392 | 0.004765 | Animal organ morphogenesis, embryonic body morphogenesis, cytokine-mediated signaling pathway, sequestering of BMP from receptor via BMP binding, determination of dorsal identity, and regulation of cytokine activity. |
| *HAS1* | Hyaluronan synthase 1 | -3.32818 | 0.019406 | Polysaccharide biosynthetic process, glycosaminoglycan biosynthetic process, cell adhesion, negative regulation of fibroblast migration, hyaluronan biosynthetic process, cellular response to platelet-derived growth factor stimulus, and extracellular matrix assembly. |
| *WBP11P3* | WBP11 pseudogene 3 | -3.31509 | 0.019406 | Unknown function. |
| *PBOV1* | Prostate and breast cancer overexpressed 1 | -3.2659 | 0.028085 | Nucleoplasm, cytosol, and centriolar satellite. |
| *LY6K* | Lymphocyte antigen 6 family member K | -3.20499 | 0.01171 | Binding of sperm to zona pellucida. |
| *ENSG00000 286555* | Novel transcript, antisense to ARAP1 | -3.15558 | 0.026416 | *Pseudogene*. |

#### Supplementary Materials

| | | | | |
|---|---|---|---|---|
| *ENSG00000 271452* | Novel transcript, sense intronic to MRPL19 | -3.05892 | 0.02688 | *Pseudogene*. |
| *ENSG00000 257668* | Suppression of tumorigenicity 13 (colon carcinoma) (Hsp70 interacting protein) (ST13) pseudogene | -3.03289 | 0.038701 | *Pseudogene*. |
| *ENSG00000 287458* | Novel transcript, antisense to FKBP5 | -2.99629 | 0.046808 | *Pseudogene*. |
| *ENSG00000 222007* | Novel transcript, antisense to AGAP1 | -2.98678 | 0.044792 | *Pseudogene*. |
| *EGR1* | Early growth response 1 | -2.74702 | 0.000423 | Negative regulation of transcription by RNA polymerase II, response to hypoxia, response to ischemia, regulation of transcription by RNA polymerase II, response to glucose, positive regulation of gene expression, T cell differentiation, BMP signaling pathway, positive regulation of chemokine production, positive regulation of interleukin-1 beta production, response to insulin, circadian regulation of gene expression, regulation of protein sumoylation, skeletal muscle cell differentiation, regulation of neuron apoptotic process, positive regulation of gene expression via chromosomal CpG island demethylation, estrous cycle, locomotor rhythm, positive regulation of DNA-templated transcription, positive regulation of transcription by RNA polymerase II, positive regulation of hormone biosynthetic process, rhythmic process, circadian temperature homeostasis, interleukin-1-mediated signaling pathway, cellular response to gamma radiation, cellular response to heparin, cellular response to mycophenolic acid, glomerular mesangial cell proliferation, positive regulation of glomerular metanephric mesangial cell proliferation, negative regulation of canonical Wnt signaling pathway, cellular response to interleukin-8, positive regulation of post-translational protein modification, positive regulation of miRNA transcription, and regulation of progesterone biosynthetic process. |
| *DUSP1* | Dual specificity phosphatase 1 | -2.67327 | 0.002633 | Endoderm formation, chromatin remodeling, regulation of transcription by RNA polymerase II, negative regulation of cell adhesion, signal transduction, negative regulation of cell population proliferation, peptidyl-tyrosine dephosphorylation, peptidyl-threonine dephosphorylation, negative regulation of MAP kinase activity, negative regulation of MAPK cascade, negative regulation of meiotic cell cycle, peptidyl-serine dephosphorylation, negative regulation of monocyte chemotaxis, regulation of mitotic cell cycle spindle assembly checkpoint, negative regulation of p38MAPK cascade, and cellular response to chemokine. |
| *RPLP1P6* | Ribosomal protein lateral stalk subunit P1 pseudogene 6 | -2.66456 | 0.005713 | Unknown function. |
| *ATF3* | Activating transcription factor 3 | -2.60739 | 0.027549 | Negative regulation of transcription by RNA polymerase II, gluconeogenesis, regulation of transcription by RNA polymerase II, positive regulation of cell population proliferation, positive regulation of gene expression, endoplasmic reticulum unfolded protein response, cellular response to amino acid starvation, response to endoplasmic reticulum stress, skeletal muscle cell differentiation, positive regulation of transcription by RNA polymerase II, negative regulation of ERK1 and ERK2 cascade, and positive regulation of TRAIL-activated apoptotic signaling pathway. |
| *AREG* | Amphiregulin | -2.56811 | 0.024517 | Epidermal growth factor receptor signaling pathway, G protein-coupled receptor signaling pathway, cell-cell signaling, positive regulation of cell population proliferation, positive regulation of keratinocyte proliferation, glial cell proliferation, neuron projection development, response to estradiol, ERBB2-EGFR signaling pathway, response to hydrogen peroxide, response to peptide hormone, negative regulation of osteoblast differentiation, positive regulation of peptidyl-tyrosine phosphorylation, response to glucocorticoid, response to cAMP, dichotomous subdivision of terminal units involved in mammary gland duct morphogenesis, mammary gland branching involved in thelarche, mammary gland alveolus development, and epithelial cell proliferation involved in mammary gland duct elongation. |
| *RASD1* | Ras related dexamethasone induced 1 | -2.54255 | 0.000423 | Signal transduction, G protein-coupled receptor signaling pathway, nitric oxide mediated signal transduction, and negative regulation of DNA-templated transcription. |

#### Supplementary Materials

| | | | | |
|---|---|---|---|---|
| *PCOLCE2* | Procollagen C-endopeptidase enhancer 2 | -2.40737 | 0.017711 | Cellular response to leukemia inhibitory factor. |
| *NR4A1* | Nuclear receptor subfamily 4 group A member 1 | -2.36886 | 0.010203 | Positive regulation of endothelial cell proliferation, response to amphetamine, cell migration involved in sprouting angiogenesis, regulation of transcription by RNA polymerase II, transcription by RNA polymerase II, apoptotic process, inflammatory response, signal transduction, neurotransmitter secretion involved in regulation of skeletal muscle contraction, intracellular receptor signaling pathway, detection of lipopolysaccharide, endothelial cell chemotaxis, skeletal muscle cell differentiation, cellular response to vascular endothelial growth factor stimulus, positive regulation of apoptotic process, cellular response to fibroblast growth factor stimulus, fat cell differentiation, negative regulation of cell cycle, positive regulation of transcription by RNA polymerase II, response to electrical stimulus, regulation of type B pancreatic cell proliferation, cellular response to corticotropin-releasing hormone stimulus, and non-canonical inflammasome complex assembly. |
| *ZDHHC11* | Zinc finger DHHC-type containing 11 | -2.24334 | 0.041459 | Positive regulation of defense response to virus by host, protein targeting to membrane, peptidyl-L-cysteine S-palmitoylation, antiviral innate immune response. |
| *LAMB2P1* | Laminin subunit beta 2 pseudogene 1 | -2.24321 | 0.010767 | Unknown function. |
| *FXYD1* | FXYD domain containing ion transport regulator 1 | -2.17712 | 0.037743 | Monoatomic ion transport, potassium ion transport, sodium ion transport, chloride transport, muscle contraction, regulation of heart contraction, negative regulation of protein glutathionylation, regulation of monoatomic ion transport, regulation of cardiac muscle cell membrane potential, chloride transmembrane transport, positive regulation of sodium ion export across plasma membrane, regulation of sodium ion transmembrane transporter activity. |
| *C1QTNF7* | C1q and TNF related 7 | -2.12199 | 0.037166 | Extracellular region and collagen trimer. |
| *CCDC126* | Coiled-coil domain containing 126 | -2.00246 | 0.043949 | Extracellular region and membrane. |
| *C1orf74* | Chromosome 1 open reading frame 74 | -2.00202 | 0.010203 | Unknown function. |
| *CES1* | carboxylesterase 1 | -1.98343 | 0.035852 | Cholesterol biosynthetic process, cholesterol metabolic process, response to toxic substance, positive regulation of cholesterol efflux, negative regulation of cholesterol storage, lipid catabolic process, epithelial cell differentiation, cholesterol homeostasis, reverse cholesterol transport, medium-chain fatty acid metabolic process, regulation of bile acid biosynthetic process, cellular response to cholesterol, cellular response to low-density lipoprotein particle stimulus, cholesterol ester hydrolysis involved in cholesterol transport, positive regulation of cholesterol metabolic process, and regulation of bile acid secretion, |
| *SLPI* | Secretory leukocyte peptidase inhibitor | -1.97865 | 0.033849 | Immune response, antibacterial humoral response, negative regulation of protein binding, response to lipopolysaccharide, modulation of process of another organism, negative regulation of viral genome replication, and innate immune response. |
| *JUN* | Jun proto-oncogene, AP-1 transcription factor subunit | -1.90884 | 0.009749 | Negative regulation of transcription by RNA polymerase II, regulation of transcription by RNA polymerase II, transforming growth factor beta receptor signaling pathway, release from viral latency, cellular response to reactive oxygen species, response to endoplasmic reticulum stress, regulation of cell population proliferation, positive regulation of apoptotic process, negative regulation of DNA binding, negative regulation by host of viral transcription, positive regulation by host of viral transcription, negative regulation of DNA-templated transcription, positive regulation of DNA-templated transcription, positive regulation of transcription by RNA polymerase II, response to steroid hormone, regulation of cell cycle, SMAD protein signal transduction, cellular response to cadmium ion, integrated stress response signaling, positive regulation of miRNA transcription, positive regulation of vascular associated smooth muscle cell proliferation, and positive regulation of DNA-templated transcription initiation. |
| *ERRFI1* | ERBB receptor feedback inhibitor 1 | -1.87947 | $1.49 \times 10^{-6}$ | Liver development, tissue homeostasis, glucose metabolic process, bile acid biosynthetic process, apoptotic process, epidermal growth factor receptor signaling pathway, negative regulation of epidermal growth factor-activated receptor activity, embryo implantation, cholesterol metabolic process, response to xenobiotic stimulus, gene expression, cell migration, negative regulation of protein autophosphorylation, response to estradiol, response to progesterone, response to insulin, uterine epithelium |

#### Supplementary Materials

| | | | | |
|---|---|---|---|---|
| | | | | development, chondrocyte proliferation, limb joint morphogenesis, negative regulation of epidermal growth factor receptor signaling pathway, cholesterol homeostasis, phosphatidylinositol 3-kinase/protein kinase B signal transduction, skin morphogenesis, regulation of keratinocyte differentiation, lung alveolus development, epithelial cell proliferation, progesterone receptor signaling pathway, cartilage development, lung vasculature development, lung epithelium development, fat pad development, protein localization to plasma membrane, response to 1-oleoyl-sn-glycerol 3-phosphate. |
| *NMNAT3* | Nicotinamide nucleotide adenylyltransferase 3 | -1.82257 | 0.040689 | tRNA processing, biosynthetic process, nucleotide biosynthetic process, NAD biosynthetic process, response to wounding, response to tumor necrosis factor. |
| *SNCA* | Synuclein alpha | -1.81389 | 0.044792 | Negative regulation of transcription by RNA polymerase II, microglial cell activation, positive regulation of receptor recycling, positive regulation of neurotransmitter secretion, negative regulation of protein kinase activity, fatty acid metabolic process, neutral lipid metabolic process, phospholipid metabolic process, mitochondrial membrane organization, chemical synaptic transmission, adult locomotory behavior, response to xenobiotic stimulus, response to iron(II) ion, regulation of phospholipase activity, negative regulation of platelet-derived growth factor receptor signaling pathway, regulation of glutamate secretion, regulation of dopamine secretion, synaptic vesicle exocytosis, synaptic vesicle priming, regulation of transmembrane transporter activity, negative regulation of microtubule polymerization, receptor internalization, protein destabilization, response to magnesium ion, negative regulation of transporter activity, response to lipopolysaccharide, negative regulation of monooxygenase activity, positive regulation of peptidyl-serine phosphorylation, response to type II interferon, cellular response to oxidative stress, SNARE complex assembly, positive regulation of SNARE complex assembly, regulation of locomotion, dopamine biosynthetic process, mitochondrial ATP synthesis coupled electron transport, regulation of macrophage activation, positive regulation of apoptotic process, negative regulation of apoptotic process, negative regulation of neuron apoptotic process, positive regulation of endocytosis, negative regulation of exocytosis, positive regulation of exocytosis, regulation of long-term neuronal synaptic plasticity, synaptic vesicle endocytosis, synaptic vesicle transport, positive regulation of inflammatory response, synapse organization, regulation of acyl-CoA biosynthetic process, protein tetramerization, positive regulation of release of sequestered calcium ion into cytosol, neuron apoptotic process, dopamine uptake involved in synaptic transmission, negative regulation of dopamine uptake involved in synaptic transmission, negative regulation of serotonin uptake, regulation of norepinephrine uptake, negative regulation of norepinephrine uptake, excitatory postsynaptic potential, long-term synaptic potentiation, positive regulation of inositol phosphate biosynthetic process, negative regulation of thrombin-activated receptor signaling pathway, response to interleukin-1, cellular response to copper ion, cellular response to epinephrine stimulus, positive regulation of protein serine/threonine kinase activity, supramolecular fiber organization, negative regulation of mitochondrial electron transport, NADH to ubiquinone, positive regulation of glutathione peroxidase activity, positive regulation of hydrogen peroxide catabolic process, regulation of synaptic vesicle recycling, regulation of reactive oxygen species biosynthetic process, positive regulation of protein localization to cell periphery, negative regulation of chaperone-mediated autophagy, regulation of presynapse assembly, and amyloid fibril formation. |
| *MYC* | MYC proto-oncogene, bHLH transcription factor | -1.80453 | 0.00384 | G1/S transition of mitotic cell cycle, negative regulation of transcription by RNA polymerase II, MAPK cascade, branching involved in ureteric bud morphogenesis, positive regulation of mesenchymal cell proliferation, chromatin remodeling, regulation of DNA-templated transcription, regulation of transcription by RNA polymerase II, intracellular iron ion homeostasis, DNA damage response, positive regulation of cell population proliferation, response to xenobiotic stimulus, response to gamma radiation, regulation of gene expression, regulation of cell cycle process, positive regulation of gene expression, rRNA metabolic process, regulation of telomere maintenance, positive regulation of telomere maintenance, negative regulation of stress-activated MAPK cascade, protein-DNA complex disassembly, cellular response to UV, negative regulation of apoptotic process, negative regulation of gene expression via chromosomal CpG island methylation, fibroblast apoptotic process, negative regulation of monocyte differentiation, positive regulation |

| | | | | |
|---|---|---|---|---|
| | | | | of translation, positive regulation of DNA-templated transcription, positive regulation of transcription by RNA polymerase II, positive regulation of fibroblast proliferation, negative regulation of fibroblast proliferation, positive regulation of epithelial cell proliferation, chromosome organization, negative regulation of cell division, negative regulation of transcription initiation by RNA polymerase II, ERK1 and ERK2 cascade, response to growth factor, cellular response to hypoxia, cellular response to xenobiotic stimulus, positive regulation of metanephric cap mesenchymal cell proliferation, positive regulation of intrinsic apoptotic signaling pathway by p53 class mediator, positive regulation of miRNA transcription, regulation of somatic stem cell population maintenance, and positive regulation of cysteine-type endopeptidase activity. |
| *ZFP36* | ZFP36 ring finger protein | -1.78862 | 0.002066 | Negative regulation of transcription by RNA polymerase II, MAPK cascade, nuclear-transcribed mRNA catabolic process, deadenylation-dependent decay, nuclear-transcribed mRNA poly(A) tail shortening, mRNA catabolic process, response to wounding, regulation of keratinocyte proliferation, myeloid cell differentiation, nuclear-transcribed mRNA catabolic process, deadenylation-independent decay, regulation of tumor necrosis factor production, negative regulation of interleukin-2 production, negative regulation of viral transcription, miRNA-mediated gene silencing by inhibition of translation, p38MAPK cascade, response to starvation, regulation of mRNA stability, cellular response to fibroblast growth factor stimulus, positive regulation of fat cell differentiation, regulation of keratinocyte differentiation, negative regulation of erythrocyte differentiation, negative regulation of inflammatory response, mRNA transport, positive regulation of nuclear-transcribed mRNA poly(A) tail shortening, hematopoietic stem cell differentiation, 3'-UTR-mediated mRNA destabilization, 3'-UTR-mediated mRNA stabilization, cellular response to lipopolysaccharide, cellular response to tumor necrosis factor, cellular response to epidermal growth factor stimulus, cellular response to glucocorticoid stimulus, cellular response to granulocyte macrophage colony-stimulating factor stimulus, positive regulation of nuclear-transcribed mRNA catabolic process, deadenylation-dependent decay, positive regulation of deadenylation-independent decapping of nuclear-transcribed mRNA, negative regulation of hematopoietic stem cell differentiation, regulation of keratinocyte apoptotic process, negative regulation of polynucleotide adenylyltransferase activity, positive regulation of intracellular mRNA localization, and positive regulation of miRNA-mediated gene silencing. |
| *BMP2* | Bone morphogenetic protein 2 | -1.78573 | 0.007568 | Negative regulation of transcription by RNA polymerase II, skeletal system development, ossification, osteoblast differentiation, branching involved in ureteric bud morphogenesis, response to hypoxia, in utero embryonic development, epithelial to mesenchymal transition, positive regulation of protein phosphorylation, chondrocyte differentiation, heart induction, endodermal-mesodermal cell signaling, aortic valve development, atrioventricular valve morphogenesis, endocardial cushion morphogenesis, cardiac atrium formation, endocardial cushion formation, positive regulation of extracellular matrix constituent secretion, proteoglycan metabolic process, regulation of DNA-templated transcription, transcription by RNA polymerase II, inflammatory response, Notch signaling pathway, cell-cell signaling, heart development, positive regulation of cell population proliferation, negative regulation of cell population proliferation, response to bacterium, animal organ morphogenesis, positive regulation of gene expression, negative regulation of gene expression, positive regulation of epithelial to mesenchymal transition, negative regulation of steroid biosynthetic process, positive regulation of phosphatase activity, telencephalon development, telencephalon regionalization, cell differentiation, positive regulation of Wnt signaling pathway, bone mineralization, osteoclast differentiation, positive regulation of cell migration, positive regulation of bone mineralization, BMP signaling pathway, negative regulation of transforming growth factor beta receptor signaling pathway, protein destabilization, positive regulation of protein binding, negative regulation of aldosterone biosynthetic process, positive regulation of osteoblast proliferation, cardiocyte differentiation, embryonic heart tube anterior/posterior pattern specification, positive regulation of peroxisome proliferator activated receptor signaling pathway, ameloblast differentiation, odontogenesis of dentin-containing tooth, positive regulation of odontogenesis, regulation of odontogenesis of dentin-containing tooth, positive regulation of apoptotic process, negative regulation of MAP |

| | | | | |
|---|---|---|---|---|
| | | | | kinase activity, positive regulation of MAPK cascade, negative regulation of insulin-like growth factor receptor signaling pathway, cell fate commitment, negative regulation of fat cell differentiation, positive regulation of fat cell differentiation, positive regulation of neuron differentiation, positive regulation of osteoblast differentiation, positive regulation of ossification, negative regulation of cell cycle, negative regulation of DNA-templated transcription, positive regulation of DNA-templated transcription, positive regulation of transcription by RNA polymerase II, negative regulation of smooth muscle cell proliferation, astrocyte differentiation, positive regulation of astrocyte differentiation, mesenchymal cell differentiation, inner ear development, negative regulation of calcium-independent cell-cell adhesion, positive regulation of DNA-binding transcription factor activity, negative regulation of muscle cell differentiation, cartilage development, cardiac muscle cell differentiation, cardiac muscle tissue morphogenesis, pericardium development, corticotropin hormone secreting cell differentiation, thyroid-stimulating hormone-secreting cell differentiation, cardiac epithelial to mesenchymal transition, bone development, positive regulation of SMAD protein signal transduction, lung vasculature development, mesenchyme development, positive regulation of cartilage development, positive regulation of ERK1 and ERK2 cascade, cellular response to organic cyclic compound, cellular response to BMP stimulus, mesenchymal cell proliferation involved in ureteric bud development, negative regulation of canonical Wnt signaling pathway, positive regulation of bone mineralization involved in bone maturation, positive regulation of p38MAPK cascade, positive regulation of odontoblast differentiation, positive regulation of miRNA transcription, cardiac jelly development, atrioventricular canal morphogenesis, negative regulation of cortisol biosynthetic process, and negative regulation of cardiac muscle cell differentiation. |
| *MT1E* | Metallothionein 1E | -1.70537 | 0.003381 | Intracellular zinc ion homeostasis, detoxification of copper ion, negative regulation of growth, cellular response to cadmium ion, cellular response to copper ion, and cellular response to zinc ion. |
| *MT2A* | Metallothionein 2A | -1.6971 | 0.021926 | Intracellular copper ion homeostasis, intracellular zinc ion homeostasis, detoxification of copper ion, cellular response to interleukin-3, cellular response to erythropoietin, negative regulation of growth, cellular response to cadmium ion, cellular response to copper ion, and cellular response to zinc ion. |
| *WEE1* | WEE1 G2 checkpoint kinase | -1.57374 | 0.01218 | G2/M transition of mitotic cell cycle, microtubule cytoskeleton organization, mitotic cell cycle, chromatin remodeling, negative regulation of G2/M transition of mitotic cell cycle, establishment of cell polarity, positive regulation of DNA replication, neuron projection morphogenesis, cell division, negative regulation of G2/MI transition of meiotic cell cycle, and negative regulation of G1/S transition of mitotic cell cycle. |
| *SLC19A2* | Solute carrier family 19 member 2 | -1.54853 | 0.004782 | Spermatogenesis, thiamine diphosphate biosynthetic process, folic acid transport, thiamine transport, pyridoxine transport, thiamine-containing compound metabolic process, transmembrane transport, and thiamine transmembrane transport. |
| *TIPARP* | TCDD inducible poly(ADP-ribose) polymerase | -1.54127 | 0.000165 | Vasculogenesis, kidney development, androgen metabolic process, estrogen metabolic process, female gonad development, post-embryonic development, negative regulation of gene expression, hemopoiesis, positive regulation of protein catabolic process, platelet-derived growth factor receptor signaling pathway, skeletal system morphogenesis, smooth muscle tissue development, roof of mouth development, face morphogenesis, cellular response to organic cyclic compound, and response to 2,3,7,8-tetrachlorodibenzodioxine. |
| *GSTM3* | Glutathione S-transferase mu 3 | -1.52241 | 0.01348 | Glutathione metabolic process, establishment of blood-nerve barrier, nitrobenzene metabolic process, xenobiotic catabolic process, response to estrogen, and cellular detoxification of nitrogen compound. |

*P-values were corrected by multiple hypothesis testing to give the Q-value by the Benjamini-Hochberg method. **Functional Annotation is adapted from Database for Annotation, Visualization and Integrated Discovery (DAVID).

#### Supplementary Materials

**Supplemental Table 8.** Top 50 Upregulated Genes in Circumscribed LS *versus* HC.

| Symbol | Protein Encoded | logFC | Q-Value* | Functional Annotation** |
|---|---|---|---|---|
| *PLA2G2D* | Phospholipase A2 group IID | 7.846367 | 0.006867 | CD4-positive, CD25-positive, alpha-beta regulatory T cell differentiation, regulation of acute inflammatory response to antigenic stimulus, phospholipid metabolic process, inflammatory response, lipid catabolic process, negative regulation of T cell proliferation, phosphatidylethanolamine metabolic process, phosphatidylcholine metabolic process, phosphatidylglycerol metabolic process, and arachidonate secretion. |
| *CXCL10* | C-X-C motif chemokine ligand 10 | 7.450173 | 0.014074 | Chemotaxis, inflammatory response, signal transduction, cell surface receptor signaling pathway, G protein-coupled receptor signaling pathway, adenylate cyclase-activating G protein-coupled receptor signaling pathway, cell-cell signaling, muscle organ development, blood circulation, positive regulation of cell population proliferation, response to gamma radiation, T cell chemotaxis, regulation of T cell chemotaxis, response to auditory stimulus, negative regulation of angiogenesis, neutrophil chemotaxis, killing of cells of another organism, response to vitamin D, cellular response to heat, endothelial cell activation, regulation of cell population proliferation, regulation of apoptotic process, negative regulation of myoblast differentiation, positive regulation of transcription by RNA polymerase II, positive chemotaxis, positive regulation of release of sequestered calcium ion into cytosol, antimicrobial humoral immune response mediated by antimicrobial peptide, chemokine-mediated signaling pathway, cellular response to lipopolysaccharide, positive regulation of monocyte chemotaxis, cellular response to interleukin-17, cellular response to virus, antiviral innate immune response, regulation of endothelial tube morphogenesis, negative regulation of myoblast fusion, and positive regulation of T cell migration. |
| *CCL3L1* | C-C motif chemokine ligand 3 like 3 | 7.059067 | 0.004747 | Inflammatory response, immune response, negative regulation of cell population proliferation, positive regulation of cell migration, eosinophil chemotaxis, cell chemotaxis, antimicrobial humoral immune response mediated by antimicrobial peptide, and chemokine-mediated signaling pathway. |
| *TTC24* | Tetratricopeptide repeat domain 24 | 6.973114 | 0.000736 | Unknown function. |
| *CXCL11* | C-X-C motif chemokine ligand 11 | 6.970333 | 0.001958 | Chemotaxis, inflammatory response, signal transduction, adenylate cyclase-activating G protein-coupled receptor signaling pathway, cell-cell signaling, T cell chemotaxis, neutrophil chemotaxis, killing of cells of another organism, regulation of cell population proliferation, positive regulation of release of sequestered calcium ion into cytosol, antimicrobial humoral immune response mediated by antimicrobial peptide, chemokine-mediated signaling pathway, and cellular response to lipopolysaccharide. |
| *ADAMDEC1* | ADAM like decysin 1 | 6.954252 | 0.027805 | Proteolysis, immune response, negative regulation of cell adhesion, integrin-mediated signaling pathway. |
| *GZMK* | Granzyme K | 6.817184 | 0.009219 | Proteolysis, protein maturation, granzyme-mediated programmed cell death signaling pathway. |
| *FCRL3* | Fc receptor like 3 | 6.790222 | 0.026317 | Negative regulation of immunoglobulin production, immune response, cell surface receptor signaling pathway, single fertilization, positive regulation of B cell proliferation, regulation of toll-like receptor 9 signaling pathway, positive regulation of MAPK cascade, regulation of B cell differentiation, negative regulation of B cell receptor signaling pathway, regulation of B cell activation and regulation of calcium ion import. |
| *IGHGP* | Immunoglobulin heavy constant gamma P (non-functional) | 6.74447 | 0.03464 | Unknown function. |
| *LAG3* | lymphocyte activating 3 | 6.692189 | $3.38 \times 10^{-6}$ | Adaptive immune response, plasmacytoid dendritic cell activation, cell surface receptor signaling pathway, negative regulation of interleukin-2 production, T cell activation, natural killer cell mediated cytotoxicity, innate immune response, negative regulation of regulatory T cell differentiation, positive regulation of natural killer cell mediated cytotoxicity and regulation of immune response. |
| *IGHV3-30* | Immunoglobulin heavy variable 3-30 | 6.625022 | 0.026235 | Immune response and immunoglobulin mediated immune response. |

#### Supplementary Materials

| | | | | |
|---|---|---|---|---|
| *TRAJ6* | T cell receptor alpha joining 6 | 6.617791 | 0.002873 | Unknown function. |
| *MS4A1* | Membrane spanning 4-domains A1 | 6.557053 | 0.040451 | Store-operated calcium entry, humoral immune response, cell surface receptor signaling pathway, response to bacterium, B cell differentiation, B cell proliferation, B cell activation, B cell receptor signaling pathway, protein tetramerization, calcium ion import into cytosol, and positive regulation of calcium ion import across plasma membrane. |
| *GZMA* | Granzyme A | 6.553916 | 0.020462 | Apoptotic process, immune response, response to bacterium, killing of cells of another organism, negative regulation of endodeoxyribonuclease activity, positive regulation of apoptotic process, negative regulation of DNA binding, negative regulation of oxidoreductase activity, proteolysis involved in protein catabolic process, protein maturation, pyroptotic inflammatory response, granzyme-mediated programmed cell death signaling pathway, and cytotoxic T cell pyroptotic cell death. |
| *FCRL6* | Fc receptor like 6 | 6.507585 | 0.000427 | Immune response and cell surface receptor signaling pathway. |
| *IDO1* | Indoleamine 2,3-dioxygenase 1 | 6.414389 | 0.009143 | Immune system process, positive regulation of T cell tolerance induction, positive regulation of chronic inflammatory response, positive regulation of type 2 immune response, tryptophan catabolic process, inflammatory response, female pregnancy, tryptophan catabolic process to kynurenine, quinolinate biosynthetic process, response to lipopolysaccharide, negative regulation of interleukin-10 production, positive regulation of interleukin-12 production, multicellular organismal response to stress, kynurenic acid biosynthetic process, 'de novo' NAD biosynthetic process from tryptophan, swimming behavior, T cell proliferation, negative regulation of T cell proliferation, positive regulation of apoptotic process, negative regulation of activated T cell proliferation, negative regulation of T cell apoptotic process, and positive regulation of T cell apoptotic process. |
| *IGHV3-23* | Immunoglobulin heavy variable 3-23 | 6.412313 | 0.039195 | Immune response and immunoglobulin mediated immune response. |
| *FCGR1A* | Fc gamma receptor Ia | 6.38978 | 0.032364 | Antibody-dependent cellular cytotoxicity, positive regulation of type IIa hypersensitivity, positive regulation of type III hypersensitivity, receptor-mediated endocytosis, phagocytosis, recognition, phagocytosis, engulfment, immune response, signal transduction, cell surface receptor signaling pathway, positive regulation of tumor necrosis factor production, Fc-gamma receptor signaling pathway, antigen processing and presentation of exogenous peptide antigen via MHC class I, defense response to bacterium, innate immune response, and positive regulation of phagocytosis. |
| *SLC12A3* | Solute carrier family 12 member 3 | 6.263333 | 0.00166 | Response to dietary excess, monoatomic ion transport, sodium ion transport, cell volume homeostasis, signal transduction, sodium ion transmembrane transport, chloride ion homeostasis, potassium ion homeostasis, sodium ion homeostasis, renal sodium ion absorption, chloride transmembrane transport, response to aldosterone, potassium ion import across plasma membrane. |
| *GZMB* | Granzyme B | 6.251824 | 0.01091 | Proteolysis, apoptotic process, negative regulation of translation, killing of cells of another organism, natural killer cell mediated cytotoxicity, proteolysis involved in protein catabolic process, protein maturation, pyroptotic inflammatory response, granzyme-mediated programmed cell death signaling pathway, positive regulation of establishment of protein localization to mitochondrion. |
| *IGHJ5* | Immunoglobulin heavy joining 5 | 6.21613 | 0.007065 | Unknown function. |
| *IGKV1-6* | Immunoglobulin kappa variable 1-6 | 6.113421 | 0.017917 | Adaptive immune response and immune response. |
| *PACSIN1* | Protein kinase C and casein kinase substrate in neurons 1 | 6.091967 | 0.000617 | Cytoskeleton organization, actin filament organization, regulation of endocytosis, negative regulation of endocytosis, synaptic vesicle endocytosis, neuron projection morphogenesis, protein localization to membrane, protein localization to plasma membrane, plasma membrane tubulation, positive regulation of dendrite development, |
| *UBASH3A* | Ubiquitin associated and SH3 domain containing A | 6.044887 | 0.005433 | Regulation of cytokine production, negative regulation of T cell receptor signaling pathway. |
| *CRTAM* | Cytotoxic and regulatory T cell molecule | 6.026627 | 0.005677 | Establishment of T cell polarity, positive regulation of cytokine production, adaptive immune response, detection of tumor cell, positive regulation of natural killer cell mediated cytotoxicity directed against tumor cell target, heterophilic cell-cell adhesion via plasma membrane cell adhesion molecules, cell recognition, positive regulation of type II interferon |

| | | | | |
|---|---|---|---|---|
| | | | | production, regulation of T cell differentiation, positive regulation of natural killer cell mediated cytotoxicity, negative regulation of activated T cell proliferation, regulation of T cell activation, detection of stimulus, lymphocyte migration into lymphoid organs, regulation of CD8-positive, and alpha-beta T cell activation. |
| *PLA1A* | Phospholipase A1 member A | 5.971587 | 0.008348 | Lipid metabolic process, phosphatidylserine metabolic process, lipid catabolic process, phosphatidylserine acyl-chain remodeling, |
| *TIGIT* | T cell immunoreceptor with Ig and ITIM domains | 5.942895 | 0.004866 | Negative regulation of interleukin-12 production, positive regulation of interleukin-10 production, negative regulation of natural killer cell mediated cytotoxicity, negative regulation of T cell activation, |
| *NUGGC* | Nuclear GTPase, germinal center associated | 5.885355 | 0.005882 | Regulation of nuclear cell cycle DNA replication, negative regulation of apoptotic process, cellular response to lipopolysaccharide, |
| *EOMES* | Eomesodermin | 5.87264 | 0.020924 | Negative regulation of transcription by RNA polymerase II, endoderm formation, mesoderm formation, endodermal cell fate specification, neuron migration, trophectodermal cell differentiation, adaptive immune response, CD8-positive, alpha-beta T cell differentiation involved in immune response, chromatin remodeling, regulation of transcription by RNA polymerase II, brain development, cardioblast differentiation, gene expression, stem cell population maintenance, olfactory bulb development, cerebral cortex regionalization, cerebral cortex neuron differentiation, skeletal muscle cell differentiation, negative regulation of DNA-binding transcription factor activity, positive regulation of DNA-templated transcription, positive regulation of transcription by RNA polymerase II, mesendoderm development, astrocyte differentiation, cell differentiation involved in embryonic placenta development, mesodermal to mesenchymal transition involved in gastrulation, |
| *TRAJ47* | T cell receptor alpha joining 47 | 5.78917 | 0.000517 | Unknown function. |
| *SLCO5A1* | Solute carrier organic anion transporter family member 5A1 | 5.774601 | 0.001016 | Sodium-independent organic anion transport, transmembrane transport. |
| *GZMH* | Granzyme H | 5.638661 | 0.015552 | Proteolysis, apoptotic process, killing of cells of another organism, protein maturation. |
| *IL4I1* | Interleukin 4 induced 1 | 5.613917 | 0.000991 | Adaptive immune response, regulation of adaptive immune response, negative regulation of T cell mediated immune response to tumor cell, L-phenylalanine catabolic process, tryptophan catabolic process, tyrosine catabolic process, amino acid catabolic process, tryptophan catabolic process to indole-3-acetate, negative regulation of T cell proliferation, regulation of B cell differentiation, positive regulation of regulatory T cell differentiation, positive regulation of transcription by RNA polymerase II, negative regulation of T cell activation. |
| *GBP5* | Guanylate binding protein 5 | 5.56275 | 0.040073 | Activation of innate immune response, protein targeting, inflammatory response, positive regulation of interleukin-1 beta production, positive regulation of interleukin-18 production, protein localization to Golgi apparatus, defense response to bacterium, innate immune response, positive regulation of innate immune response, protein homotetramerization, defense response to virus, cytolysis in another organism, cellular response to lipopolysaccharide, cellular response to type II interferon, positive regulation of pyroptotic inflammatory response, positive regulation of AIM2 inflammasome complex assembly, positive regulation of cytokine production involved in inflammatory response, positive regulation of NLRP3 inflammasome complex assembly. |
| *TRAJ27* | T cell receptor alpha joining 27 | 5.556808 | 0.006995 | Unknown function. |
| *CETP* | Cholesteryl ester transfer protein | 5.556032 | 0.000989 | Triglyceride metabolic process, lipid transport, cholesterol metabolic process, negative regulation of macrophage derived foam cell differentiation, regulation of cholesterol efflux, phospholipid transport, cholesterol transport, positive regulation of cholesterol transport, triglyceride transport, very-low-density lipoprotein particle remodeling, low-density lipoprotein particle remodeling, high-density lipoprotein particle remodeling, cholesterol homeostasis, reverse cholesterol transport, phosphatidylcholine metabolic process, lipid homeostasis, phospholipid homeostasis, triglyceride homeostasis, and positive regulation of phospholipid transport. |
| *CERKL* | Ceramide kinase like | 5.5407 | 0.004781 | Sphingolipid metabolic process, sphingolipid biosynthetic process, and negative regulation of apoptotic process. |

### Supplementary Materials

| | | | | |
|---|---|---|---|---|
| *FHAD1* | Forkhead associated phosphopeptide binding domain 1 | 5.514903 | 0.003452 | Unknown function. |
| *IFNG* | Interferon gamma | 5.502609 | 0.004222 | Negative regulation of transcription by RNA polymerase II, microglial cell activation, positive regulation of cytokine production, adaptive immune response, macrophage activation involved in immune response, apoptotic process, humoral immune response, cell surface receptor signaling pathway, cell surface receptor signaling pathway via JAK-STAT, positive regulation of cell population proliferation, response to virus, positive regulation of autophagy, positive regulation of gene expression, negative regulation of gene expression, positive regulation of epithelial cell migration, regulation of metabolic process, macrophage differentiation, negative regulation of epithelial cell differentiation, positive regulation of protein-containing complex assembly, negative regulation of interleukin-17 production, positive regulation of chemokine production, positive regulation of interleukin-1 beta production, positive regulation of interleukin-12 production, positive regulation of interleukin-23 production, positive regulation of interleukin-6 production, positive regulation of tumor necrosis factor production, positive regulation of CD4-positive, CD25-positive, alpha-beta regulatory T cell differentiation involved in immune response, positive regulation of peptidyl-serine phosphorylation of STAT protein, positive regulation of smooth muscle cell apoptotic process, Fc-gamma receptor signaling pathway involved in phagocytosis, type III interferon-mediated signaling pathway, positive regulation of protein import into nucleus, positive regulation of tyrosine phosphorylation of STAT protein, positive regulation of MHC class II biosynthetic process, positive regulation of nitric oxide biosynthetic process, positive regulation of osteoclast differentiation, positive regulation of glycolytic process, negative regulation of DNA-templated transcription, positive regulation of transcription by RNA polymerase II, astrocyte activation, negative regulation of smooth muscle cell proliferation, positive regulation of inflammatory response, positive regulation of phagocytosis, positive regulation of neurogenesis, regulation of insulin secretion, positive regulation of membrane protein ectodomain proteolysis, defense response to virus, positive regulation of killing of cells of another organism, positive regulation of nitric-oxide synthase biosynthetic process, type II interferon-mediated signaling pathway, positive regulation of fructose 1,6-bisphosphate 1-phosphatase activity, positive regulation of fructose 1,6-bisphosphate metabolic process, positive regulation of vitamin D biosynthetic process, positive regulation of calcidiol 1-monooxygenase activity, positive regulation of protein serine/threonine kinase activity, positive regulation of protein deacetylation, extrinsic apoptotic signaling pathway, cell surface receptor signaling pathway via STAT, cellular response to virus, neuroinflammatory response, negative regulation of amyloid-beta clearance, positive regulation of cellular respiration, positive regulation of amyloid-beta formation, negative regulation of tau-protein kinase activity, positive regulation of protein localization to plasma membrane, positive regulation of exosomal secretion, positive regulation of iron ion import across plasma membrane, positive regulation of NMDA glutamate receptor activity, positive regulation of core promoter binding, positive regulation of signaling receptor activity, and positive regulation of tumor necrosis factor (ligand) superfamily member 11 production. |
| *TRAJ9* | T cell receptor alpha joining 9 | 5.400578 | 0.001628 | Unknown function. |
| *IGHV3-20* | Immunoglobulin heavy variable 3-20 | 5.391024 | 0.02324 | Adaptive immune response and immunoglobulin mediated immune response. |
| *IGKV3D-15* | Immunoglobulin kappa variable 3D-15 | 5.365235 | 0.031434 | Adaptive immune response and immune response. |
| *IGHV3-33* | Immunoglobulin heavy variable 3-33 | 5.323658 | 0.048232 | Adaptive immune response, immune response, and immunoglobulin mediated immune response. |
| *TRAJ7* | T cell receptor alpha joining 7 | 5.305635 | 0.004158 | Unknown function. |
| *SI* | Sucrase-isomaltase | 5.290422 | 0.014201 | Sucrose catabolic process, polysaccharide digestion. |
| *FCRL5* | Fc receptor like 5 | 5.278893 | 0.039791 | Immune response, cell surface receptor signaling pathway, single fertilization, B cell activation. |
| *C11orf42* | Chromosome 11 open reading frame 42 | 5.272745 | 0.001937 | Unknown function. |

| | | | | |
|---|---|---|---|---|
| *SIRPG* | Signal regulatory protein gamma | 5.22519 | 0.006911 | Cell adhesion, cell-cell signaling, positive regulation of cell population proliferation, negative regulation of cell population proliferation, positive regulation of cell-cell adhesion, intracellular signal transduction, positive regulation of phagocytosis, positive regulation of T cell activation, |
| *MZB1* | Marginal zone B and B1 cell specific protein | 5.224271 | 0.01048 | Positive regulation of immunoglobulin production, apoptotic process, positive regulation of cell population proliferation, regulation of B cell proliferation, integrin activation, regulation of cell population proliferation, and regulation of insulin receptor signaling pathway. |
| *IGHJ6* | Immunoglobulin heavy joining 6 | 5.221338 | 0.006237 | Unknown function. |

*P-values were corrected by multiple hypothesis testing to give the Q-value by the Benjamini-Hochberg method. **Functional Annotation is adapted from Database for Annotation, Visualization and Integrated Discovery (DAVID).

**Supplemental Table 9.** Top 50 Downregulated Genes in Circumscribed LS *vs.* HC.

| **Gene** | **Protein Encoded** | **logFC** | **Q-Value*** | **Functional Annotation**** |
|---|---|---|---|---|
| *FOSB* | FosB proto-oncogene, AP-1 transcription factor subunit | -4.68826 | 0.010539 | Negative regulation of transcription by RNA polymerase II, response to amphetamine, regulation of transcription by RNA polymerase II, transcription by RNA polymerase II, female pregnancy, response to xenobiotic stimulus, response to mechanical stimulus, response to progesterone, cellular response to hormone stimulus, response to morphine, response to ethanol, positive regulation of transcription by RNA polymerase II, behavioral response to cocaine, response to corticosterone, response to cAMP, and cellular response to calcium ion. |
| *WIF1* | WNT inhibitory factor 1 | -4.39104 | 0.004523 | Blood vessel development, signal transduction, determination of left/right symmetry, heart development, anterior/posterior pattern specification, Wnt signaling pathway, negative regulation of Wnt signaling pathway, nodal signaling pathway, and positive regulation of fat cell differentiation. |
| *TNMD* | Tenomodulin | -4.27343 | 0.005185 | Endothelial cell morphogenesis, negative regulation of endothelial cell proliferation, and negative regulation of angiogenesis. |
| *FOS* | Fos proto-oncogene, AP-1 transcription factor subunit | -4.2277 | 0.000813 | Conditioned taste aversion, neural retina development, regulation of transcription by RNA polymerase II, transcription by RNA polymerase II, inflammatory response, transforming growth factor beta receptor signaling pathway, nervous system development, female pregnancy, response to xenobiotic stimulus, response to light stimulus, response to gravity, response to toxic substance, response to activity, skeletal muscle cell proliferation, cerebral cortex development, neuron differentiation, osteoclast differentiation, response to lipopolysaccharide, response to progesterone, response to insulin, cellular response to zinc ion starvation, cellular response to reactive oxygen species, response to immobilization stress, skeletal muscle cell differentiation, response to muscle stretch, response to ethanol, positive regulation of osteoclast differentiation, positive regulation of DNA-templated transcription, positive regulation of transcription by RNA polymerase II, response to corticosterone, myoblast proliferation, response to cAMP, SMAD protein signal transduction, cellular response to cadmium ion, cellular response to calcium ion, cellular response to tumor necrosis factor, cellular response to epidermal growth factor stimulus, cellular response to parathyroid hormone stimulus, cellular response to hypoxia, cellular response to transforming growth factor beta stimulus, medium-term memory, integrated stress response signaling, positive regulation of miRNA transcription, mononuclear cell differentiation, cellular response to phorbol 13-acetate 12-myristate, and cellular response to prolactin. |
| *LY6K* | Lymphocyte antigen 6 family member K | -4.14496 | 0.000163 | Binding of sperm to zona pellucida. |
| *MATN4* | Matrilin 4 | -4.01375 | 0.008985 | Extracellular matrix organization. |
| *VEGFD* | Vascular endothelial growth factor D | -3.95141 | 0.013835 | Response to hypoxia, positive regulation of protein phosphorylation, positive regulation of endothelial cell proliferation, sprouting angiogenesis, positive regulation of cell population proliferation, response to bacterium, vascular endothelial growth factor signaling pathway, positive regulation of angiogenesis, vascular endothelial growth factor receptor signaling pathway, fibroblast proliferation, positive chemotaxis, induction of positive chemotaxis, positive regulation of cell division, positive regulation of mast cell chemotaxis, and dopaminergic neuron differentiation. |

#### Supplementary Materials

| | | | | |
|---|---|---|---|---|
| *LYVE1* | Lymphatic vessel endothelial hyaluronan receptor 1 | -3.8022 | 0.00075 | Positive regulation of cellular extravasation, receptor-mediated endocytosis, cell adhesion, cell-matrix adhesion, response to wounding, anatomical structure morphogenesis, and hyaluronan catabolic process. |
| *AHSP* | Alpha hemoglobin stabilizing protein | -3.76507 | 0.021604 | Protein folding, hemoglobin metabolic process, hemopoiesis, erythrocyte differentiation, and protein stabilization. |
| *CES1P1* | Carboxylesterase 1 pseudogene 1 | -3.69957 | 0.022235 | Anatomical structure morphogenesis, and lipid catabolic process. |
| *BMP3* | Bone morphogenetic protein 3 | -3.66231 | 0.004156 | Skeletal system development, osteoblast differentiation, signal transduction, cell-cell signaling, negative regulation of ossification, and cartilage development. |
| *PCOLCE2* | Procollagen C-endopeptidase enhancer 2 | -3.59924 | 0.000349 | Cellular response to leukemia inhibitory factor. |
| *CLDN11* | Claudin 11 | -3.56394 | 0.001874 | Cell adhesion, spermatogenesis, axon ensheathment, calcium-independent cell-cell adhesion via plasma membrane cell-adhesion molecules, bicellular tight junction assembly, and tight junction assembly. |
| *ENSG00000 167046* | Novel transcript, antisense to SLCO4A1 | -3.51529 | 0.009446 | *Pseudogene*. |
| *MAB21L1* | mab-21 like 1 | -3.50427 | 0.017489 | Eye development, cell population proliferation, positive regulation of cell population proliferation, anatomical structure morphogenesis, and camera-type eye development. |
| *EDN3* | Endothelin 3 | -3.46589 | 0.032021 | Neural crest cell migration, positive regulation of leukocyte chemotaxis, regulation of systemic arterial blood pressure by endothelin, intracellular calcium ion homeostasis, signal transduction, cell surface receptor signaling pathway, cell-cell signaling, axon guidance, blood circulation, cell population proliferation, positive regulation of cell population proliferation, positive regulation of heart rate, regulation of gene expression, intracellular magnesium ion homeostasis, vein smooth muscle contraction, regulation of vasoconstriction, peptide hormone secretion, melanocyte differentiation, neutrophil chemotaxis, vasoconstriction, positive regulation of MAP kinase activity, positive regulation of cell differentiation, positive regulation of mitotic nuclear division, positive regulation of smooth muscle contraction, positive regulation of hormone secretion, regulation of developmental pigmentation, axon extension, establishment of localization in cell, potassium ion transmembrane transport, and positive regulation of potassium ion transmembrane transport. |
| *ASS1P9* | Argininosuccinate synthetase 1 pseudogene 9 | -3.41593 | 0.0076 | Unknown function. |
| *EGR1* | Early growth response 1 | -3.341 | 0.00011 | Negative regulation of transcription by RNA polymerase II, response to hypoxia, response to ischemia, regulation of transcription by RNA polymerase II, response to glucose, positive regulation of gene expression, T cell differentiation, BMP signaling pathway, positive regulation of chemokine production, positive regulation of interleukin-1 beta production, response to insulin, circadian regulation of gene expression, regulation of protein sumoylation, skeletal muscle cell differentiation, regulation of neuron apoptotic process, positive regulation of gene expression via chromosomal CpG island demethylation, estrous cycle, locomotor rhythm, positive regulation of DNA-templated transcription, positive regulation of transcription by RNA polymerase II, positive regulation of hormone biosynthetic process, rhythmic process, circadian temperature homeostasis, interleukin-1-mediated signaling pathway, cellular response to gamma radiation, cellular response to heparin, cellular response to mycophenolic acid, glomerular mesangial cell proliferation, positive regulation of glomerular metanephric mesangial cell proliferation, negative regulation of canonical Wnt signaling pathway, cellular response to interleukin-8, positive regulation of post-translational protein modification, positive regulation of miRNA transcription, and regulation of progesterone biosynthetic process. |
| *ENSG00000 286555* | Novel transcript, antisense to ARAP1 | -3.28986 | 0.005846 | *Pseudogene*. |
| *CES1* | Carboxylesterase 1 | -3.28469 | 0.000101 | Anatomical structure morphogenesis and lipid catabolic process. |
| *PRSS42P* | Serine protease 42, pseudogene | -3.26384 | 0.042038 | Proteolysis, spermatogenesis, and cell differentiation. |

#### Supplementary Materials

| | | | | |
|---|---|---|---|---|
| *ENSG00000 262202* | Novel transcript | -3.26158 | 0.0141 | *Pseudogene*. |
| *MT1G* | Metallothionein 1G | -3.23642 | 0.008785 | Intracellular zinc ion homeostasis, detoxification of copper ion, monocyte differentiation, cellular response to vascular endothelial growth factor stimulus, monocyte activation, negative regulation of growth, cellular response to cadmium ion, cellular response to copper ion, and cellular response to zinc ion. |
| *DUSP1* | Dual specificity phosphatase 1 | -3.19481 | 0.000543 | Endoderm formation, chromatin remodeling, regulation of transcription by RNA polymerase II, negative regulation of cell adhesion, signal transduction, negative regulation of cell population proliferation, peptidyl-tyrosine dephosphorylation, peptidyl-threonine dephosphorylation, negative regulation of MAP kinase activity, negative regulation of MAPK cascade, negative regulation of meiotic cell cycle, peptidyl-serine dephosphorylation, negative regulation of monocyte chemotaxis, regulation of mitotic cell cycle spindle assembly checkpoint, negative regulation of p38MAPK cascade, and cellular response to chemokine. |
| *CLEC3B* | C-type lectin domain family 3 member B | -3.17462 | 0.000437 | Ossification, positive regulation of plasminogen activation, and bone mineralization. |
| *TNXA* | Tenascin XA (pseudogene) | -3.14042 | 0.04919 | Extracellular exosome. |
| *GREM2* | Gremlin 2, DAN family BMP antagonist | -3.12661 | 0.002165 | Animal organ morphogenesis, embryonic body morphogenesis, cytokine-mediated signaling pathway, sequestering of BMP from receptor via BMP binding, determination of dorsal identity, and regulation of cytokine activity. |
| *SERPINA5* | Serpin family A member 5 | -3.11843 | 0.017729 | Lipid transport, spermatogenesis, fusion of sperm to egg plasma membrane involved in single fertilization, and negative regulation of hydrolase activity. |
| *LINC00482* | Long intergenic non-protein coding RNA 482 | -3.10413 | 0.014201 | Unknown function. |
| *WBP11P3* | WBP11 pseudogene 3 | -3.07025 | 0.015707 | Unknown function. |
| *CYP4Z2P* | Cytochrome P450 family 4 subfamily Z member 2, pseudogene | -3.06174 | 0.005984 | Membrane |
| *ENSG00000 250770* | Tetraspanin 11 (TSPAN11) pseudogene | -3.06099 | 0.014357 | *Pseudogene*. |
| *RPLP1P6* | Ribosomal protein lateral stalk subunit P1 pseudogene 6 | -3.03741 | 0.001744 | Unknown function. |
| *CYP4Z1* | Cytochrome P450 family 4 subfamily Z member 1 | -3.01757 | 0.037985 | Arachidonate metabolic process and lauric acid metabolic process. |
| *ENSG00000 288700* | Novel transcrpt | -2.95153 | 0.047857 | *Pseudogene*. |
| *ENSG00000 222007* | Novel transcript, antisense to AGAP1 | -2.92334 | 0.037671 | *Pseudogene*. |
| *NR4A1* | Nuclear receptor subfamily 4 group A member 1 | -2.89431 | 0.001937 | Positive regulation of endothelial cell proliferation, response to amphetamine, cell migration involved in sprouting angiogenesis, regulation of transcription by RNA polymerase II, transcription by RNA polymerase II, apoptotic process, inflammatory response, signal transduction, neurotransmitter secretion involved in regulation of skeletal muscle contraction, intracellular receptor signaling pathway, detection of lipopolysaccharide, endothelial cell chemotaxis, skeletal muscle cell differentiation, cellular response to vascular endothelial growth factor stimulus, positive regulation of apoptotic process, cellular response to fibroblast growth factor stimulus, fat cell differentiation, negative regulation of cell cycle, positive regulation of transcription by RNA polymerase II, response to electrical stimulus, regulation of type B pancreatic cell proliferation, cellular response to corticotropin-releasing hormone stimulus, and non-canonical inflammasome complex assembly. |
| *PBOV1* | Prostate and breast cancer overexpressed 1 | -2.87205 | 0.024764 | Nucleoplasm, cytosol, and centriolar satellite. |

#### Supplementary Materials

| | | | | |
|---|---|---|---|---|
| *TCEAL7* | Transcription elongation factor A like 7 | -2.86371 | 0.032377 | Negative regulation of NF-kappaB transcription factor activity, and negative regulation of DNA-templated transcription. |
| *ATP5MC1P6* | ATP synthase membrane subunit c locus 1 pseudogene 6 | -2.82571 | 0.030682 | Unknown function. |
| *KLF14* | KLF transcription factor 14 | -2.82067 | 0.049802 | Regulation of transcription by RNA polymerase II, positive regulation of transcription by RNA polymerase II, and positive regulation of sphingolipid mediated signaling pathway. |
| *OR51B6* | Olfactory receptor family 51 subfamily B member 6 | -2.80666 | 0.035002 | G protein-coupled receptor signaling pathway, sensory perception of smell, detection of chemical stimulus involved in sensory perception of smell. |
| *TMEM100* | Transmembrane protein 100 | -2.80072 | 0.044552 | Angiogenesis, vasculogenesis, in utero embryonic development, epithelial to mesenchymal transition involved in endocardial cushion formation, Notch signaling pathway, BMP signaling pathway, positive regulation of endothelial cell differentiation, regulation of calcium-mediated signaling, positive regulation of phosphatidylinositol 3-kinase/protein kinase B signal transduction, regulation of sensory perception of pain, arterial endothelial cell differentiation, cellular response to BMP stimulus, and positive regulation of vasculogenesis. |
| *RPL7P60* | Ribosomal protein L7 pseudogene 60 | -2.79521 | 0.035914 | Unknown function. |
| *ENSG00000 213068* | Ribosomal protein S17 (RPS17) pseudogene | -2.79199 | 0.021528 | *Pseudogene*. |
| *ENSG00000 249593* | Novel transcript | -2.7751 | 0.036552 | *Pseudogene*. |
| *RBM24* | RNA binding motif protein 24 | -2.76537 | 0.012653 | Regulation of alternative mRNA splicing, via spliceosome, endocardial cushion development, mRNA processing, DNA damage response, RNA splicing, regulation of myotube differentiation, positive regulation of myotube differentiation, cell differentiation, regulation of mRNA stability, positive regulation of myoblast differentiation, mRNA stabilization, mRNA destabilization, 3'-UTR-mediated mRNA destabilization, positive regulation of skeletal muscle fiber differentiation, positive regulation of 3'-UTR-mediated mRNA stabilization, positive regulation of stem cell differentiation, negative regulation of cytoplasmic translation. |
| *PDGFRL* | Platelet derived growth factor receptor like | -2.75117 | 0.00106 | G protein-coupled receptor signaling pathway, platelet-derived growth factor receptor-beta signaling pathway. |
| *CYP4F26P* | Cytochrome P450 family 4 subfamily F member 26, pseudogene | -2.74443 | 0.018666 | Unknown function. |
| *SLPI* | Secretory leukocyte peptidase inhibitor | -2.72685 | 0.002145 | Immune response, antibacterial humoral response, negative regulation of protein binding, response to lipopolysaccharide, modulation of process of another organism, negative regulation of viral genome replication, innate immune response. |

*P-values were corrected by multiple hypothesis testing to give the Q-value by the Benjamini-Hochberg method. **Functional Annotation is adapted from Database for Annotation, Visualization and Integrated Discovery (DAVID).

#### Supplementary Materials

**Supplemental Table 5.** Top 50 Upregulated Genes in Generalized LS *vs.* HC.

| Gene | Protein Encoded | logFC | Q-Value* | Functional Annotation** |
|---|---|---|---|---|
| *LAG3* | Lymphocyte activating 3 | 5.942803 | 0.000414 | Adaptive immune response, plasmacytoid dendritic cell activation, cell surface receptor signaling pathway, negative regulation of interleukin-2 production, T cell activation, natural killer cell mediated cytotoxicity, innate immune response, negative regulation of regulatory T cell differentiation, positive regulation of natural killer cell mediated cytotoxicity, and regulation of immune response. |
| *TDO2* | Tryptophan 2,3-dioxygenase | 5.670039 | 0.017576 | Tryptophan metabolic process, tryptophan catabolic process to kynurenine, tryptophan catabolic process to acetyl-CoA, protein homotetramerization, and response to nitroglycerin. |
| *SLC12A3* | Solute carrier family 12 member 3 | 5.355284 | 0.047669 | Response to dietary excess, monoatomic ion transport, sodium ion transport, cell volume homeostasis, signal transduction, sodium ion transmembrane transport, chloride ion homeostasis, potassium ion homeostasis, sodium ion homeostasis, renal sodium ion absorption, chloride transmembrane transport, response to aldosterone, and potassium ion import across plasma membrane. |
| *FHAD1* | Forkhead associated phosphopeptide binding domain 1 | 5.27887 | 0.035055 | Unknown function. |
| *FCRL6* | Fc receptor like 6 | 5.180693 | 0.034519 | Immune response and cell surface receptor signaling pathway. |
| *C11orf42* | Chromosome 11 open reading frame 42 | 5.147841 | 0.022466 | Unknown function. |
| *PACSIN1* | Protein kinase C and casein kinase substrate in neurons 1 | 5.109105 | 0.034519 | Cytoskeleton organization, actin filament organization, regulation of endocytosis, negative regulation of endocytosis, synaptic vesicle endocytosis, neuron projection morphogenesis, protein localization to membrane, protein localization to plasma membrane, plasma membrane tubulation, and positive regulation of dendrite development. |
| *OR1J1* | Olfactory receptor family 1 subfamily J member 1 | 5.104323 | 0.034519 | Signal transduction, G protein-coupled receptor signaling pathway, and detection of chemical stimulus involved in sensory perception of smell. |
| *A1BG-AS1* | A1BG antisense RNA 1 | 4.83235 | 0.002671 | Unknown function. |
| *IL4I1* | Interleukin 4 induced 1 | 4.822522 | 0.034519 | Adaptive immune response, regulation of adaptive immune response, negative regulation of T cell mediated immune response to tumor cell, L-phenylalanine catabolic process, tryptophan catabolic process, tyrosine catabolic process, amino acid catabolic process, tryptophan catabolic process to indole-3-acetate, negative regulation of T cell proliferation, regulation of B cell differentiation, positive regulation of regulatory T cell differentiation, positive regulation of transcription by RNA polymerase II, and negative regulation of T cell activation. |
| *TLE6* | TLE family member 6, subcortical maternal complex member | 4.202398 | 0.043949 | Negative regulation of transcription by RNA polymerase II, regulation of DNA-templated transcription, actin filament organization, positive regulation of embryonic development, establishment of spindle localization, regulation of cell division, endoplasmic reticulum localization, mitochondrion localization, embryonic process involved in female pregnancy, and negative regulation of canonical Wnt signaling pathway. |
| *RSAD2* | Radical S-adenosyl methionine domain containing 2 | 4.168445 | 0.010969 | Response to virus, positive regulation of toll-like receptor 7 signaling pathway, positive regulation of toll-like receptor 9 signaling pathway, CD4-positive, alpha-beta T cell activation, CD4-positive, alpha-beta T cell differentiation, negative regulation of viral genome replication, innate immune response, negative regulation of protein secretion, positive regulation of immune response, defense response to virus, and positive regulation of T-helper 2 cell cytokine production. |
| *ABHD17AP6* | ABHD17A pseudogene 6 | 4.012437 | 0.010969 | Unknown function. |
| *TRAJ37* | T cell receptor alpha joining 37 | 3.988853 | 0.034519 | Unknown function. |
| *REM1* | RRAD and GEM like GTPase 1 | 3.910138 | 0.025395 | Regulation of skeletal muscle contraction by calcium ion signaling, negative regulation of calcium ion transmembrane transport via high voltage-gated calcium channel, |
| *NTN3* | Netrin 3 | 3.811308 | 0.048514 | Regulation of transcription by RNA polymerase II, and axon guidance. |
| *SPTY2D1OS* | Mitochondrial sheath formation associated | 3.731458 | 0.034519 | Unknown function. |

### Supplementary Materials

| | | | | |
|---|---|---|---|---|
| *LOXHD1* | Lipoxygenase homology PLAT domains 1 | 3.721866 | 0.037611 | Sensory perception of sound. |
| *RPGRIP1* | RPGR interacting protein 1 | 3.679216 | 0.034519 | Visual perception, retinal rod cell development, neural precursor cell proliferation, non-motile cilium assembly. |
| *SIGLEC15* | Sialic acid binding Ig like lectin 15 | 3.675846 | 0.049167 | Regulation of actin cytoskeleton organization, regulation of bone resorption, regulation of osteoclast development. |
| *TNFRSF4* | TNF receptor superfamily member 4 | 3.638565 | 0.030527 | Positive regulation of immunoglobulin production, inflammatory response, immune response, positive regulation of B cell proliferation, tumor necrosis factor-mediated signaling pathway, T cell proliferation, negative regulation of DNA-binding transcription factor activity, negative regulation of DNA-templated transcription, and symbiont entry into host cell. |
| *INHBA* | Inhibin subunit beta A | 3.296578 | 0.036202 | Ovarian follicle development, positive regulation of protein phosphorylation, hair follicle development, hematopoietic progenitor cell differentiation, regulation of transcription by RNA polymerase II, transcription by RNA polymerase II, autophagy, defense response, cell surface receptor signaling pathway, cell-cell signaling, nervous system development, androgen metabolic process, negative regulation of cell population proliferation, male gonad development, positive regulation of gene expression, cytokine-mediated signaling pathway, striatal medium spiny neuron differentiation, cell differentiation, erythrocyte differentiation, negative regulation of cell growth, negative regulation of type II interferon production, activin receptor signaling pathway, positive regulation of collagen biosynthetic process, endodermal cell differentiation, negative regulation of phosphorylation, odontogenesis, hemoglobin biosynthetic process, progesterone secretion, negative regulation of B cell differentiation, positive regulation of erythrocyte differentiation, negative regulation of macrophage differentiation, positive regulation of DNA-templated transcription, positive regulation of transcription by RNA polymerase II, positive regulation of transcription by RNA polymerase III, regulation of follicle-stimulating hormone secretion, positive regulation of follicle-stimulating hormone secretion, negative regulation of follicle-stimulating hormone secretion, mesodermal cell differentiation, positive regulation of protein metabolic process, negative regulation of hair follicle development, Sertoli cell differentiation, roof of mouth development, positive regulation of ovulation, positive regulation of SMAD protein signal transduction, SMAD protein signal transduction, cardiac fibroblast cell development, eyelid development in camera-type eye, testosterone biosynthetic process, positive regulation of ERK1 and ERK2 cascade, cellular response to follicle-stimulating hormone stimulus, cellular response to cholesterol, cellular response to hypoxia, cellular response to oxygen-glucose deprivation, GABAergic neuron differentiation, extrinsic apoptotic signaling pathway, androst-4-ene-3,17-dione biosynthetic process, response to aldosterone, cellular response to angiotensin, negative regulation of G1/S transition of mitotic cell cycle, and positive regulation of extrinsic apoptotic signaling pathway in absence of ligand, |
| *PCDH17* | Protocadherin 17 | 3.263631 | 0.017783 | Cell adhesion, homophilic cell adhesion via plasma membrane adhesion molecules, adult behavior, negative regulation of synaptic transmission, synaptic membrane adhesion, presynaptic active zone assembly, and regulation of synaptic vesicle clustering. |
| *ACTN2* | Actinin alpha 2 | 3.215432 | 0.03891 | Cell adhesion, microspike assembly, actin cytoskeleton organization, regulation of membrane potential, regulation of apoptotic process, negative regulation of potassium ion transport, positive regulation of potassium ion transport, sarcomere organization, positive regulation of DNA-templated transcription, focal adhesion assembly, actin filament uncapping, muscle cell development, cardiac muscle cell development, protein localization to plasma membrane, phospholipase C-activating angiotensin-activated signaling pathway, postsynaptic actin cytoskeleton organization, negative regulation of potassium ion transmembrane transporter activity, positive regulation of potassium ion transmembrane transporter activity, negative regulation of protein localization to cell surface, positive regulation of endocytic recycling, and positive regulation of cation channel activity. |
| *ENSG00000224611* | TBPL1 pseudogene 1 | 3.11397 | 0.045705 | *Pseudogene.* |

#### Supplementary Materials

| | | | | |
|---|---|---|---|---|
| *FAM78A* | Family with sequence similarity 78 member A | 3.086257 | 0.043949 | Unknown function. |
| *HAVCR2* | Hepatitis A virus cellular receptor 2 | 2.887053 | 0.045084 | Adaptive immune response, macrophage activation involved in immune response, natural killer cell tolerance induction, regulation of tolerance induction dependent upon immune response, negative regulation of T-helper 1 type immune response, negative regulation of natural killer cell mediated cytotoxicity directed against tumor cell target, regulation of transcription by RNA polymerase II, inflammatory response, negative regulation of gene expression, negative regulation of myeloid dendritic cell activation, negative regulation of NF-kappaB transcription factor activity, negative regulation of interferon-alpha production, negative regulation of type II interferon production, negative regulation of interleukin-2 production, negative regulation of interleukin-3 production, negative regulation of interleukin-6 production, negative regulation of tumor necrosis factor production, positive regulation of chemokine production, positive regulation of type II interferon production, positive regulation of interleukin-1 production, positive regulation of interleukin-4 production, positive regulation of tumor necrosis factor production, negative regulation of natural killer cell activation, toll-like receptor 3 signaling pathway, toll-like receptor 7 signaling pathway, toll-like receptor 9 signaling pathway, positive regulation of T cell proliferation, negative regulation of T cell proliferation, positive regulation of macrophage activation, innate immune response, defense response to Gram-positive bacterium, maternal process involved in female pregnancy, positive regulation of ERK1 and ERK2 cascade, cellular response to lipopolysaccharide, negative regulation of granulocyte colony-stimulating factor production, negative regulation of defense response to bacterium, positive regulation of defense response to bacterium, positive regulation of non-canonical NF-kappaB signal transduction, negative regulation of immunological synapse formation, negative regulation of T cell activation via T cell receptor contact with antigen bound to MHC molecule on antigen presenting cell. |
| *SCN1B* | Sodium voltage-gated channel beta subunit 1 | 2.852842 | 0.043949 | Sodium ion transport, cell adhesion, axon guidance, positive regulation of sodium ion transport, positive regulation of neuron projection development, neuronal action potential propagation, corticospinal neuron axon guidance, monoatomic ion transmembrane transport, sodium ion transmembrane transport, locomotion, membrane depolarization, cardiac muscle contraction, regulation of ventricular cardiac muscle cell membrane repolarization, regulation of atrial cardiac muscle cell membrane depolarization, cardiac conduction, cardiac muscle cell action potential involved in contraction, membrane depolarization during action potential, membrane depolarization during cardiac muscle cell action potential, membrane depolarization during Purkinje myocyte cell action potential, regulation of heart rate by cardiac conduction, regulation of sodium ion transmembrane transport, and positive regulation of voltage-gated sodium channel activity. |
| *VAMP5* | Vesicle associated membrane protein 5 | 2.505949 | 0.034519 | Muscle organ development, skeletal muscle tissue development, cell differentiation, Golgi to plasma membrane protein transport. |
| *IL15* | Interleukin 15 | 2.481169 | 0.049167 | Natural killer cell differentiation, natural killer cell proliferation, positive regulation of cytokine production, NK T cell proliferation, immune response, signal transduction, tyrosine phosphorylation of STAT protein, cell-cell signaling, positive regulation of cell population proliferation, macrophage differentiation, positive regulation of interleukin-17 production, positive regulation of natural killer cell proliferation, positive regulation of natural killer cell differentiation, positive regulation of tissue remodeling, interleukin-15-mediated signaling pathway, positive regulation of T cell proliferation, neutrophil activation, positive regulation of tyrosine phosphorylation of STAT protein, extrathymic T cell selection, regulation of T cell differentiation, cell maturation, lymph node development, regulation of defense response to virus by host, positive regulation of inflammatory response, positive regulation of peptidyl-tyrosine phosphorylation, positive regulation of phagocytosis, positive regulation of immune response, negative regulation of cold-induced thermogenesis, and positive regulation of protein O-linked glycosylation. |
| *RAB29* | RAB29, member RAS oncogene family | 2.338901 | 0.034519 | Positive regulation of receptor recycling, intracellular protein transport, mitochondrion organization, Golgi organization, synapse assembly, response to bacterium, negative regulation of neuron projection development, vesicle-mediated transport, cell differentiation, melanosome organization, T cell activation, retrograde transport, endosome to |

| | | | | |
|---|---|---|---|---|
| | | | | Golgi, modulation by host of viral process, positive regulation of T cell receptor signaling pathway, protein localization to membrane, positive regulation of intracellular protein transport, protein localization to ciliary membrane, regulation of retrograde transport, endosome to Golgi, and cellular detoxification. |
| *ENSG00000 257027* | Novel transcript | 2.258975 | 0.020608 | *Pseudogene*. |
| *ENSG00000 287263* | Novel transcript | 2.19124 | 0.029719 | *Pseudogene*. |
| *SEPTIN6* | Septin 6 | 2.072231 | 0.032211 | Mitotic cytokinesis, spermatogenesis, protein localization, cell differentiation, cilium assembly, and cytoskeleton-dependent cytokinesis. |
| *TBX2* | T-box transcription factor 2 | 2.053467 | 0.047669 | Negative regulation of transcription by RNA polymerase II, cell fate specification, heart looping, outflow tract septum morphogenesis, outflow tract morphogenesis, endocardial cushion morphogenesis, endocardial cushion formation, regulation of transcription by RNA polymerase II, apoptotic process, Notch signaling pathway, muscle cell fate determination, regulation of heart contraction, fibroblast growth factor receptor signaling pathway, neurogenesis, response to retinoic acid, embryonic heart tube development, aorta morphogenesis, atrioventricular canal development, embryonic digit morphogenesis, pigment metabolic process involved in pigmentation, negative regulation of DNA-templated transcription, positive regulation of DNA-templated transcription, positive regulation of transcription by RNA polymerase II, embryonic camera-type eye morphogenesis, cardiac muscle tissue development, smooth muscle cell differentiation, roof of mouth development, pharyngeal system development, positive regulation of cardiac muscle cell proliferation, cardiac muscle cell myoblast differentiation, epithelial tube branching involved in lung morphogenesis, developmental growth involved in morphogenesis, mammary placode formation, mesenchymal cell proliferation involved in lung development, cellular response to dexamethasone stimulus, ureteric peristalsis, cochlea morphogenesis, cellular senescence, melanocyte proliferation, negative regulation of heart looping, negative regulation of cardiac chamber formation, positive regulation of cell cycle G1/S phase transition, cardiac jelly development, atrioventricular canal morphogenesis, and negative regulation of cellular senescence. |
| *SLC38A7* | Solute carrier family 38 member 7 | 1.941876 | 0.043949 | Amino acid transmembrane transport, sodium ion transport, asparagine transport, and glutamine transport. |
| *DCBLD1* | Discoidin, CUB and LCCL domain containing 1 | 1.674635 | 0.047669 | Plasma membrane. |
| *FMNL3* | Formin like 3 | 1.671389 | 0.021927 | Angiogenesis, cytoskeleton organization, regulation of cell shape, cell migration, regulation of cell morphogenesis, actin cytoskeleton organization, cortical actin cytoskeleton organization, filopodium assembly. |
| *PRSS23* | Serine protease 23 | 1.571684 | 0.035407 | Proteolysis. |
| *STING1* | Stimulator of interferon response cGAMP interactor 1 | 1.528112 | 0.008756 | Autophagosome assembly, activation of innate immune response, pattern recognition receptor signaling pathway, positive regulation of defense response to virus by host, cytoplasmic pattern recognition receptor signaling pathway, positive regulation of macroautophagy, positive regulation of protein binding, positive regulation of type I interferon production, positive regulation of interferon-beta production, monoatomic ion transmembrane transport, cellular response to interferon-beta, innate immune response, positive regulation of transcription by RNA polymerase II, regulation of inflammatory response, positive regulation of DNA-binding transcription factor activity, protein complex oligomerization, defense response to virus, positive regulation of type I interferon-mediated signaling pathway, reticulophagy, protein localization to endoplasmic reticulum, cellular response to exogenous dsRNA, cellular response to organic cyclic compound, antiviral innate immune response, cGAS/STING signaling pathway, and proton transmembrane transport. |
| *LOXL1* | Lysyl oxidase like 1 | 1.514447 | 0.03891 | Protein deamination, collagen fibril organization, response to lipopolysaccharide, and aorta development. |
| *RUNX1* | RUNX family transcription factor 1 | 1.509627 | 0.008756 | Negative regulation of transcription by RNA polymerase II, ossification, chondrocyte differentiation, myeloid leukocyte differentiation, regulation of transcription by RNA polymerase II, regulation of plasminogen activation, hemopoiesis, myeloid cell differentiation, neuron differentiation, negative regulation of granulocyte differentiation, positive regulation of granulocyte differentiation, positive regulation of interleukin-2 |

#### Supplementary Materials

| | | | | |
|---|---|---|---|---|
| | | | | production, positive regulation of collagen biosynthetic process, negative regulation of CD4-positive, alpha-beta T cell differentiation, positive regulation of CD8-positive, alpha-beta T cell differentiation, regulation of cell differentiation, positive regulation of angiogenesis, positive regulation of DNA-templated transcription, positive regulation of transcription by RNA polymerase II, peripheral nervous system neuron development, positive regulation of developmental process, regulation of cardiac muscle cell proliferation, cardiac muscle tissue regeneration, hematopoietic stem cell proliferation, positive regulation of extracellular matrix organization, and regulation of connective tissue replacement. |
| *SMG9* | SMG9 nonsense mediated mRNA decay factor | 1.441519 | 0.006806 | Nuclear-transcribed mRNA catabolic process, nonsense-mediated decay, eye development, in utero embryonic development, brain development, heart development, and negative regulation of apoptotic process. |
| *CYP4V2* | Cytochrome P450 family 4 subfamily V member 2 | 1.391549 | 0.047669 | Retinoid metabolic process, visual perception, fatty acid omega-oxidation, and sterol metabolic process. |
| *MYD88* | MYD88 innate immune signal transduction adaptor | 1.355573 | 0.030628 | Response to molecule of fungal origin, leukocyte activation involved in inflammatory response, neutrophil activation involved in immune response, MyD88-dependent toll-like receptor signaling pathway, chromatin remodeling, phagocytosis, apoptotic process, inflammatory response, signal transduction, cell surface receptor signaling pathway, JNK cascade, Toll signaling pathway, induced systemic resistance, gene expression, positive regulation of gene expression, microglia differentiation, response to organic cyclic compound, response to amine, immunoglobulin mediated immune response, lipopolysaccharide-mediated signaling pathway, positive regulation of type I interferon production, response to peptidoglycan, positive regulation of chemokine production, positive regulation of interleukin-1 beta production, positive regulation of interleukin-17 production, positive regulation of interleukin-23 production, positive regulation of interleukin-6 production, positive regulation of interleukin-8 production, positive regulation of tumor necrosis factor production, toll-like receptor 4 signaling pathway, toll-like receptor 5 signaling pathway, toll-like receptor 8 signaling pathway, toll-like receptor TLR6:TLR2 signaling pathway, interleukin-33-mediated signaling pathway, defense response to bacterium, defense response to protozoan, positive regulation of canonical NF-kappaB signal transduction, response to amino acid, skin development, innate immune response, response to ethanol, positive regulation of transcription by RNA polymerase II, positive regulation of JNK cascade, positive regulation of smooth muscle cell proliferation, positive regulation of lymphocyte proliferation, regulation of inflammatory response, defense response to Gram-positive bacterium, positive regulation of NF-kappaB transcription factor activity, defense response to virus, type I interferon-mediated signaling pathway, positive regulation of macrophage cytokine production, interleukin-1-mediated signaling pathway, response to interleukin-1, 3'-UTR-mediated mRNA stabilization, neutrophil-mediated killing of bacterium, cellular response to lipopolysaccharide, cellular response to mechanical stimulus, establishment of endothelial intestinal barrier, cellular response to oxidised low-density lipoprotein particle stimulus, positive regulation of cytokine production involved in inflammatory response, positive regulation of NLRP3 inflammasome complex assembly, regulation of neutrophil migration, regulation of chemokine (C-X-C motif) ligand 1 production, and regulation of chemokine (C-X-C motif) ligand 2 production. |
| *TTYH3* | Tweety family member 3 | 1.270052 | 0.034519 | Chloride transport, L-glutamate transmembrane transport, monoatomic ion transmembrane transport, and chloride transmembrane transport. |
| *RFX5* | Regulatory factor X5 | 1.243338 | 0.022466 | Negative regulation of transcription by RNA polymerase II, regulation of transcription by RNA polymerase II, positive regulation of MHC class II biosynthetic process, and positive regulation of transcription by RNA polymerase II. |
| *GRAMD1A* | GRAM domain containing 1A | 1.228632 | 0.004462 | Autophagy, intracellular sterol transport, and cellular response to cholesterol. |
| *TNS3* | Tensin 3 | 1.21326 | 0.040848 | Positive regulation of Rac protein signal transduction. |
| *PARP12* | Poly(ADP-ribose) polymerase family member 12 | 1.193966 | 0.049167 | Protein auto-ADP-ribosylation. |

*P-values were corrected by multiple hypothesis testing to give the Q-value by the Benjamini-Hochberg method. **Functional Annotation is adapted from Database for Annotation, Visualization and Integrated Discovery (DAVID).

#### Supplementary Materials

**Supplemental Table 10.** Top 44 Downregulated Genes in Generalized LS *vs.* HC.

| Gene | Protein Encoded | logFC | Q-Value* | Functional Annotation** |
|---|---|---|---|---|
| *WIF1* | WNT inhibitory factor 1 | -4.55749 | 0.043949 | Blood vessel development, signal transduction, determination of left/right symmetry, heart development, anterior/posterior pattern specification, Wnt signaling pathway, negative regulation of Wnt signaling pathway, nodal signaling pathway, and positive regulation of fat cell differentiation. |
| *CLDN11* | Claudin 11 | -4.41521 | 0.003473 | Cell adhesion, spermatogenesis, axon ensheathment, calcium-independent cell-cell adhesion via plasma membrane cell-adhesion molecules, bicellular tight junction assembly, tight junction assembly. |
| *HAS1* | Hyaluronan synthase 1 | -4.39139 | 0.021416 | Polysaccharide biosynthetic process, glycosaminoglycan biosynthetic process, cell adhesion, negative regulation of fibroblast migration, hyaluronan biosynthetic process, cellular response to platelet-derived growth factor stimulus, extracellular matrix assembly. |
| *SERPINA5* | Serpin family A member 5 | -4.17743 | 0.023595 | Lipid transport, spermatogenesis, fusion of sperm to egg plasma membrane involved in single fertilization, negative regulation of hydrolase activity. |
| *GREM2* | Gremlin 2, DAN family BMP antagonist | -3.86729 | 0.004462 | Animal organ morphogenesis, embryonic body morphogenesis, cytokine-mediated signaling pathway, sequestering of BMP from receptor via BMP binding, determination of dorsal identity, regulation of cytokine activity. |
| *FOS* | Fos proto-oncogene, AP-1 transcription factor subunit | -3.7269 | 0.034519 | Conditioned taste aversion, neural retina development, regulation of transcription by RNA polymerase II, transcription by RNA polymerase II, inflammatory response, transforming growth factor beta receptor signaling pathway, nervous system development, female pregnancy, response to xenobiotic stimulus, response to light stimulus, response to gravity, response to toxic substance, response to activity, skeletal muscle cell proliferation, cerebral cortex development, neuron differentiation, osteoclast differentiation, response to lipopolysaccharide, response to progesterone, response to insulin, cellular response to zinc ion starvation, cellular response to reactive oxygen species, response to immobilization stress, skeletal muscle cell differentiation, response to muscle stretch, response to ethanol, positive regulation of osteoclast differentiation, positive regulation of DNA-templated transcription, positive regulation of transcription by RNA polymerase II, response to corticosterone, myoblast proliferation, response to cAMP, SMAD protein signal transduction, cellular response to cadmium ion, cellular response to calcium ion, cellular response to tumor necrosis factor, cellular response to epidermal growth factor stimulus, cellular response to parathyroid hormone stimulus, cellular response to hypoxia, cellular response to transforming growth factor beta stimulus, medium-term memory, integrated stress response signaling, positive regulation of miRNA transcription, mononuclear cell differentiation, cellular response to phorbol 13-acetate 12-myristate, cellular response to prolactin. |
| *TNNI2* | Troponin I2, fast skeletal type | -3.63702 | 0.049167 | Skeletal muscle contraction, positive regulation of DNA-templated transcription, cardiac muscle contraction, relaxation of skeletal muscle. |
| *PBOV1* | Prostate and breast cancer overexpressed 1 | -3.58171 | 0.040365 | Nucleoplasm, cytosol, and centriolar satellite. |
| *LY6K* | Lymphocyte antigen 6 family member K | -3.54087 | 0.019681 | Binding of sperm to zona pellucida. |
| *LYVE1* | Lmphatic vessel endothelial hyaluronan receptor 1 | -3.04804 | 0.043949 | Positive regulation of cellular extravasation, receptor-mediated endocytosis, cell adhesion, cell-matrix adhesion, response to wounding, anatomical structure morphogenesis, hyaluronan catabolic process. |
| *BDNF* | Brain derived neurotrophic factor | -2.86887 | 0.048514 | Cell surface receptor protein tyrosine kinase signaling pathway, nervous system development, axon guidance, synapse assembly, peripheral nervous system development, memory, negative regulation of myotube differentiation, positive regulation of neuron projection development, nerve development, brain-derived neurotrophic factor receptor signaling pathway, positive regulation of brain-derived neurotrophic factor receptor signaling pathway, positive regulation of peptidyl-serine phosphorylation, nerve growth factor signaling pathway, negative regulation of neuron apoptotic process, regulation of neuron differentiation, collateral sprouting, positive regulation of collateral sprouting, neuron projection morphogenesis, modulation of chemical synaptic transmission, positive regulation of synapse assembly, positive regulation of receptor |

#### Supplementary Materials

| | | | | |
|---|---|---|---|---|
| | | | | binding, regulation of protein localization to cell surface, negative regulation of apoptotic signaling pathway. |
| *RPLP1P6* | Ribosomal protein lateral stalk subunit P1 pseudogene 6 | -2.83328 | 0.033919 | Unknown function. |
| *SLPI* | Secretory leukocyte peptidase inhibitor | -2.79552 | 0.033919 | Immune response, antibacterial humoral response, negative regulation of protein binding, response to lipopolysaccharide, modulation of process of another organism, negative regulation of viral genome replication, and innate immune response. |
| *DUSP1* | Dual specificity phosphatase 1 | -2.76631 | 0.032954 | Endoderm formation, chromatin remodeling, regulation of transcription by RNA polymerase II, negative regulation of cell adhesion, signal transduction, negative regulation of cell population proliferation, peptidyl-tyrosine dephosphorylation, peptidyl-threonine dephosphorylation, negative regulation of MAP kinase activity, negative regulation of MAPK cascade, negative regulation of meiotic cell cycle, peptidyl-serine dephosphorylation, negative regulation of monocyte chemotaxis, regulation of mitotic cell cycle spindle assembly checkpoint, negative regulation of p38MAPK cascade, and cellular response to chemokine. |
| *PCOLCE2* | Procollagen C-endopeptidase enhancer 2 | -2.74598 | 0.043551 | Cellular response to leukemia inhibitory factor. |
| *EGR1* | Early growth response 1 | -2.70552 | 0.019236 | Negative regulation of transcription by RNA polymerase II, response to hypoxia, response to ischemia, regulation of transcription by RNA polymerase II, response to glucose, positive regulation of gene expression, T cell differentiation, BMP signaling pathway, positive regulation of chemokine production, positive regulation of interleukin-1 beta production, response to insulin, circadian regulation of gene expression, regulation of protein sumoylation, skeletal muscle cell differentiation, regulation of neuron apoptotic process, positive regulation of gene expression via chromosomal CpG island demethylation, estrous cycle, locomotor rhythm, positive regulation of DNA-templated transcription, positive regulation of transcription by RNA polymerase II, positive regulation of hormone biosynthetic process, rhythmic process, circadian temperature homeostasis, interleukin-1-mediated signaling pathway, cellular response to gamma radiation, cellular response to heparin, cellular response to mycophenolic acid, glomerular mesangial cell proliferation, positive regulation of glomerular metanephric mesangial cell proliferation, negative regulation of canonical Wnt signaling pathway, cellular response to interleukin-8, positive regulation of post-translational protein modification, positive regulation of miRNA transcription, and regulation of progesterone biosynthetic process. |
| *RASD1* | Ras related dexamethasone induced 1 | -2.3483 | 0.010246 | Signal transduction, G protein-coupled receptor signaling pathway, nitric oxide mediated signal transduction, and negative regulation of DNA-templated transcription. |
| *NMNAT3* | Nicotinamide nucleotide adenylyltransferase 3 | -2.11484 | 0.043949 | tRNA processing, biosynthetic process, nucleotide biosynthetic process, NAD biosynthetic process, response to wounding, and response to tumor necrosis factor. |
| *MFSD2A* | MFSD2 lysolipid transporter A, lysophospholipid | -2.03954 | 0.048961 | Retinal pigment epithelium development, phosphatidylcholine biosynthetic process, brain development, photoreceptor cell morphogenesis, carbohydrate transport, cellular response to starvation, positive regulation of triglyceride biosynthetic process, fatty acid transport, long-chain fatty acid transport, hippocampus development, positive regulation of cell growth, negative regulation of fatty acid beta-oxidation, very-low-density lipoprotein particle assembly, maintenance of blood-brain barrier, photoreceptor cell outer segment organization, regulation of multicellular organism growth, transcytosis, regulation of dendrite development, cognition, lysophospholipid transport, transmembrane transport, retina morphogenesis in camera-type eye, establishment of blood-brain barrier, motor behavior, energy homeostasis, lysophospholipid translocation, regulation of neuron projection arborization, transport across blood-brain barrier, regulation of phosphatidylcholine metabolic process, regulation of phosphatidylethanolamine metabolic process, regulation of phosphatidylserine metabolic process, and lipid transport across blood-brain barrier. |
| *ERRFI1* | ERBB receptor feedback inhibitor 1 | -1.92224 | $9.04 \times 10^{-5}$ | Liver development, tissue homeostasis, glucose metabolic process, bile acid biosynthetic process, apoptotic process, epidermal growth factor receptor signaling pathway, negative regulation of epidermal growth factor-activated receptor activity, embryo implantation, cholesterol metabolic |

| | | | | |
|---|---|---|---|---|
| | | | | process, response to xenobiotic stimulus, gene expression, cell migration, negative regulation of protein autophosphorylation, response to estradiol, response to progesterone, response to insulin, uterine epithelium development, chondrocyte proliferation, limb joint morphogenesis, negative regulation of epidermal growth factor receptor signaling pathway, cholesterol homeostasis, phosphatidylinositol 3-kinase/protein kinase B signal transduction, skin morphogenesis, regulation of keratinocyte differentiation, lung alveolus development, epithelial cell proliferation, progesterone receptor signaling pathway, cartilage development, lung vasculature development, lung epithelium development, fat pad development, protein localization to plasma membrane, response to 1-oleoyl-sn-glycerol 3-phosphate. |
| *TSC22D3* | TSC22 domain family member 3 | -1.89375 | 0.003473 | Regulation of transcription by RNA polymerase II, response to osmotic stress, negative regulation of activation-induced cell death of T cells. |
| *ATOH8* | Atonal bHLH transcription factor 8 | -1.86684 | 0.048514 | Formation of primary germ layer, negative regulation of endothelial cell proliferation, nervous system development, anatomical structure morphogenesis, positive regulation of endothelial cell migration, negative regulation of gene expression, cell differentiation, tube formation, positive regulation of endothelial cell differentiation, negative regulation of DNA-templated transcription, positive regulation of DNA-templated transcription, positive regulation of transcription by RNA polymerase II, myoblast proliferation, positive regulation of SMAD protein signal transduction, positive regulation of miRNA transcription, |
| *ZFP36* | ZFP36 ring finger protein | -1.85278 | 0.019681 | Negative regulation of transcription by RNA polymerase II, MAPK cascade, nuclear-transcribed mRNA catabolic process, deadenylation-dependent decay, nuclear-transcribed mRNA poly(A) tail shortening, mRNA catabolic process, response to wounding, regulation of keratinocyte proliferation, myeloid cell differentiation, nuclear-transcribed mRNA catabolic process, deadenylation-independent decay, regulation of tumor necrosis factor production, negative regulation of interleukin-2 production, negative regulation of viral transcription, miRNA-mediated gene silencing by inhibition of translation, p38MAPK cascade, response to starvation, regulation of mRNA stability, cellular response to fibroblast growth factor stimulus, positive regulation of fat cell differentiation, regulation of keratinocyte differentiation, negative regulation of erythrocyte differentiation, negative regulation of inflammatory response, mRNA transport, positive regulation of nuclear-transcribed mRNA poly(A) tail shortening, hematopoietic stem cell differentiation, 3'-UTR-mediated mRNA destabilization, 3'-UTR-mediated mRNA stabilization, cellular response to lipopolysaccharide, cellular response to tumor necrosis factor, cellular response to epidermal growth factor stimulus, cellular response to glucocorticoid stimulus, cellular response to granulocyte macrophage colony-stimulating factor stimulus, positive regulation of nuclear-transcribed mRNA catabolic process, deadenylation-dependent decay, positive regulation of deadenylation-independent decapping of nuclear-transcribed mRNA, negative regulation of hematopoietic stem cell differentiation, regulation of keratinocyte apoptotic process, negative regulation of polynucleotide adenylyltransferase activity, positive regulation of intracellular mRNA localization, positive regulation of miRNA-mediated gene silencing. |
| *GSTM3* | Glutathione S-transferase mu 3 | -1.84384 | 0.019681 | Glutathione metabolic process, establishment of blood-nerve barrier, nitrobenzene metabolic process, xenobiotic catabolic process, response to estrogen, cellular detoxification of nitrogen compound. |
| *MYC* | MYC proto-oncogene, bHLH transcription factor | -1.81519 | 0.023806 | G1/S transition of mitotic cell cycle, negative regulation of transcription by RNA polymerase II, MAPK cascade, branching involved in ureteric bud morphogenesis, positive regulation of mesenchymal cell proliferation, chromatin remodeling, regulation of DNA-templated transcription, regulation of transcription by RNA polymerase II, intracellular iron ion homeostasis, DNA damage response, positive regulation of cell population proliferation, response to xenobiotic stimulus, response to gamma radiation, regulation of gene expression, regulation of cell cycle process, positive regulation of gene expression, rRNA metabolic process, regulation of telomere maintenance, positive regulation of telomere maintenance, negative regulation of stress-activated MAPK cascade, protein-DNA complex disassembly, cellular response to UV, negative regulation of apoptotic process, negative regulation of gene expression via chromosomal CpG island methylation, fibroblast apoptotic process, negative regulation of monocyte differentiation, positive regulation |

| | | | | |
|---|---|---|---|---|
| | | | | of translation, positive regulation of DNA-templated transcription, positive regulation of transcription by RNA polymerase II, positive regulation of fibroblast proliferation, negative regulation of fibroblast proliferation, positive regulation of epithelial cell proliferation, chromosome organization, negative regulation of cell division, negative regulation of transcription initiation by RNA polymerase II, ERK1 and ERK2 cascade, response to growth factor, cellular response to hypoxia, cellular response to xenobiotic stimulus, positive regulation of metanephric cap mesenchymal cell proliferation, positive regulation of intrinsic apoptotic signaling pathway by p53 class mediator, positive regulation of miRNA transcription, regulation of somatic stem cell population maintenance, positive regulation of cysteine-type endopeptidase activity. |
| *MT1E* | Metallothionein 1E | -1.70916 | 0.027419 | Intracellular zinc ion homeostasis, detoxification of copper ion, negative regulation of growth, cellular response to cadmium ion, cellular response to copper ion, cellular response to zinc ion. |
| *ADRB2* | Adrenoceptor beta 2 | -1.67936 | 0.043949 | Diet induced thermogenesis, norepinephrine-epinephrine-mediated vasodilation involved in regulation of systemic arterial blood pressure, regulation of sodium ion transport, transcription by RNA polymerase II, receptor-mediated endocytosis, smooth muscle contraction, cell surface receptor signaling pathway, activation of transmembrane receptor protein tyrosine kinase activity, adenylate cyclase-modulating G protein-coupled receptor signaling pathway, endosome to lysosome transport, response to cold, positive regulation of protein kinase A signaling, positive regulation of bone mineralization, heat generation, negative regulation of multicellular organism growth, positive regulation of MAPK cascade, bone resorption, negative regulation of G protein-coupled receptor signaling pathway, positive regulation of transcription by RNA polymerase II, negative regulation of smooth muscle contraction, brown fat cell differentiation, positive regulation of mini excitatory postsynaptic potential, adrenergic receptor signaling pathway, adenylate cyclase-activating adrenergic receptor signaling pathway, positive regulation of protein serine/threonine kinase activity, AMPA selective glutamate receptor signaling pathway, positive regulation of cold-induced thermogenesis, positive regulation of cAMP/PKA signal transduction, positive regulation of autophagosome maturation, positive regulation of lipophagy, cellular response to amyloid-beta, response to psychosocial stress, positive regulation of cAMP-dependent protein kinase activity, and positive regulation of AMPA receptor activity. |
| *OCLN* | Occludin | -1.60932 | 0.047669 | Negative regulation of protein phosphorylation, positive regulation of lamellipodium assembly, positive regulation of gene expression, negative regulation of gene expression, regulation of D-glucose transmembrane transport, positive regulation of microtubule polymerization, maintenance of blood-brain barrier, cell-cell junction organization, positive regulation of D-glucose import, protein-containing complex assembly, bicellular tight junction assembly, positive regulation of wound healing, protein localization to cell leading edge, and positive regulation of blood-brain barrier permeability. |
| *AMOT* | Angiomotin | -1.5496 | 0.038635 | Negative regulation of transcription by RNA polymerase II, angiogenesis, vasculogenesis, in utero embryonic development, gastrulation with mouth forming second, establishment of cell polarity involved in ameboidal cell migration, chemotaxis, cell-cell junction assembly, protein localization, negative regulation of angiogenesis, actin cytoskeleton organization, regulation of cell migration, hippo signaling, positive regulation of embryonic development, cell migration involved in gastrulation, negative regulation of vascular permeability, blood vessel endothelial cell migration, positive regulation of blood vessel endothelial cell migration, positive regulation of cell size, regulation of small GTPase mediated signal transduction, positive regulation of stress fiber assembly, establishment of epithelial cell polarity, and regulation of modification of postsynaptic actin cytoskeleton. |
| *TIPARP* | TCDD inducible poly(ADP-ribose) polymerase | -1.52428 | 0.00257 | Vasculogenesis, kidney development, androgen metabolic process, estrogen metabolic process, female gonad development, post-embryonic development, negative regulation of gene expression, hemopoiesis, positive regulation of protein catabolic process, platelet-derived growth factor receptor signaling pathway, skeletal system morphogenesis, smooth muscle tissue development, roof of mouth development, face morphogenesis, cellular response to organic cyclic compound, response to 2,3,7,8-tetrachlorodibenzodioxine. |

### Supplementary Materials

| | | | | |
|---|---|---|---|---|
| *SLC19A2* | Solute carrier family 19 member 2 | -1.51217 | 0.034519 | Spermatogenesis, thiamine diphosphate biosynthetic process, folic acid transport, thiamine transport, pyridoxine transport, thiamine-containing compound metabolic process, transmembrane transport, thiamine transmembrane transport. |
| *RGS2* | Regulator of G protein signaling 2 | -1.46653 | 0.034519 | Response to amphetamine, G protein-coupled receptor signaling pathway, spermatogenesis, regulation of G protein-coupled receptor signaling pathway, negative regulation of phospholipase activity, negative regulation of cardiac muscle hypertrophy, positive regulation of neuron projection development, negative regulation of translation, negative regulation of MAP kinase activity, response to ethanol, negative regulation of G protein-coupled receptor signaling pathway, brown fat cell differentiation, relaxation of cardiac muscle, relaxation of vascular associated smooth muscle, maternal process involved in female pregnancy, positive regulation of cardiac muscle contraction, negative regulation of cell growth involved in cardiac muscle cell development, regulation of adenylate cyclase-inhibiting adrenergic receptor signaling pathway, and negative regulation of glycine import across plasma membrane. |
| *KLF4* | KLF transcription factor 4 | -1.45891 | 0.043949 | Negative regulation of transcription by RNA polymerase II, glandular epithelial cell differentiation, defense response to tumor cell, regulation of transcription by RNA polymerase II, transcription by RNA polymerase II, mesodermal cell fate determination, negative regulation of cell population proliferation, epidermal cell differentiation, positive regulation of gene expression, negative regulation of gene expression, negative regulation of angiogenesis, stem cell population maintenance, post-embryonic camera-type eye development, negative regulation of NF-kappaB transcription factor activity, positive regulation of telomere maintenance, response to retinoic acid, negative regulation of interleukin-8 production, negative regulation of heterotypic cell-cell adhesion, somatic stem cell population maintenance, post-embryonic hemopoiesis, negative regulation of canonical NF-kappaB signal transduction, positive regulation of nitric oxide biosynthetic process, fat cell differentiation, regulation of cell differentiation, negative regulation of DNA-templated transcription, positive regulation of DNA-templated transcription, positive regulation of transcription by RNA polymerase II, positive regulation of hemoglobin biosynthetic process, negative regulation of smooth muscle cell proliferation, regulation of axon regeneration, epidermis morphogenesis, negative regulation of inflammatory response, positive regulation of protein metabolic process, canonical Wnt signaling pathway, negative regulation of response to cytokine stimulus, establishment of skin barrier, cellular response to hydrogen peroxide, cellular response to retinoic acid, cellular response to growth factor stimulus, cellular response to cycloheximide, cellular response to laminar fluid shear stress, negative regulation of cell migration involved in sprouting angiogenesis, regulation of blastocyst development, cellular response to peptide, positive regulation of miRNA transcription, positive regulation of sprouting angiogenesis, negative regulation of leukocyte adhesion to arterial endothelial cell, cellular response to leukemia inhibitory factor, negative regulation of G1/S transition of mitotic cell cycle, negative regulation of chemokine (C-X-C motif) ligand 2 production, and negative regulation of extrinsic apoptotic signaling pathway in absence of ligand. |
| *RHOB* | Ras homolog family member B | -1.40678 | 0.019681 | Mitotic cytokinesis, angiogenesis, intracellular protein transport, apoptotic process, actin filament organization, cell adhesion, signal transduction, Rho protein signal transduction, endosome to lysosome transport, positive regulation of endothelial cell migration, cell differentiation, regulation of cell migration, negative regulation of cell migration, positive regulation of apoptotic process, positive regulation of angiogenesis, negative regulation of cell cycle, endothelial tube morphogenesis, cellular response to hydrogen peroxide, cellular response to ionizing radiation, and regulation of modification of postsynaptic structure. |
| *KLF6* | KLF transcription factor 6 | -1.3926 | 0.00257 | Regulation of transcription by RNA polymerase II, B cell differentiation, positive regulation of DNA-templated transcription, and positive regulation of transcription by RNA polymerase II. |
| *TUBB2A* | Tubulin beta 2A class IIa | -1.37101 | 0.040365 | Microtubule cytoskeleton organization, mitotic cell cycle, neuron migration, and cerebral cortex development. |
| *TMEM185B* | Transmembrane protein 185B | -1.36718 | 0.047832 | Membrane. |

#### Supplementary Materials

| | | | | |
|---|---|---|---|---|
| *CD55* | CD55 molecule (Cromer blood group) | -1.27971 | 0.049167 | Positive regulation of T cell cytokine production, complement activation, classical pathway, positive regulation of cytosolic calcium ion concentration, regulation of complement activation, regulation of lipopolysaccharide-mediated signaling pathway, innate immune response, respiratory burst, negative regulation of complement activation, symbiont entry into host cell, regulation of complement-dependent cytotoxicity, positive regulation of CD4-positive, alpha-beta T cell activation, positive regulation of CD4-positive, and alpha-beta T cell proliferation. |
| *KLF2* | KLF transcription factor 2 | -1.22954 | 0.027419 | Negative regulation of transcription by RNA polymerase II, cell morphogenesis, in utero embryonic development, regulation of transcription by RNA polymerase II, negative regulation of interleukin-6 production, multicellular organism growth, epigenetic regulation of gene expression, vasodilation, erythrocyte maturation, positive regulation of nitric oxide biosynthetic process, positive regulation of DNA-templated transcription, positive regulation of transcription by RNA polymerase II, positive regulation of retinoic acid receptor signaling pathway, positive regulation of protein metabolic process, type I pneumocyte differentiation, cellular response to hydrogen peroxide, cellular response to interleukin-1, cellular response to tumor necrosis factor, cellular response to cycloheximide, cellular response to fluid shear stress, cellular response to laminar fluid shear stress, cellular stress response to acid chemical, cellular response to peptide, negative regulation of sprouting angiogenesis. |
| *TXNL4A* | Thioredoxin like 4A | -1.19852 | 0.01528 | Spliceosomal complex assembly, RNA splicing, via transesterification reactions, mRNA splicing, via spliceosome, and cell division. |
| *TXN* | Thioredoxin | -1.14567 | 0.008756 | Negative regulation of transcription by RNA polymerase II, response to radiation, activation of protein kinase B activity, positive regulation of peptidyl-serine phosphorylation, positive regulation of DNA binding, cell redox homeostasis, negative regulation of protein export from nucleus, positive regulation of phosphatidylinositol 3-kinase/protein kinase B signal transduction, cellular detoxification of hydrogen peroxide, response to nitric oxide, and positive regulation of peptidyl-cysteine S-nitrosylation. |
| *SESN1* | Sestrin 1 | -1.10457 | 0.034519 | Positive regulation of macroautophagy, cellular response to amino acid starvation, cellular response to glucose starvation, cellular response to L-leucine, reactive oxygen species metabolic process, cellular oxidant detoxification, regulation of response to reactive oxygen species, negative regulation of TORC1 signaling, and cellular response to leucine starvation. |
| *SKP1* | S-phase kinase associated protein 1 | -1.06035 | $5.29 \times 10^{-6}$ | Protein polyubiquitination, chromatin remodeling, ubiquitin-dependent protein catabolic process, protein monoubiquitination, protein ubiquitination, SCF-dependent proteasomal ubiquitin-dependent protein catabolic process, proteasome-mediated ubiquitin-dependent protein catabolic process, maintenance of protein location in nucleus, protein K48-linked ubiquitination, and positive regulation of ubiquitin protein ligase activity. |
| *MYADM* | Myeloid associated differentiation marker | -1.04172 | 0.047669 | Negative regulation of protein phosphorylation, negative regulation of gene expression, positive regulation of cell migration, negative regulation of actin filament polymerization, membrane raft organization, negative regulation of heterotypic cell-cell adhesion, intracellular signal transduction, protein localization to plasma membrane raft, establishment of endothelial barrier, negative regulation of protein kinase C signaling, and positive regulation of substrate adhesion-dependent cell spreading. |

*P-values were corrected by multiple hypothesis testing to give the Q-value by the Benjamini-Hochberg method. **Functional Annotation is adapted from Database for Annotation, Visualization and Integrated Discovery (DAVID).

#### Supplementary Materials

**Supplemental Table 11.** Top 50 Upregulated Genes in Linear LS *vs.* HC.

| Gene | Protein Encoded | logFC | Q-Value* | Functional Annotation** |
|---|---|---|---|---|
| *IER5L* | Immediate early response 5 like | 2.643747 | 0.037881 | Unknown function. |

*P-values were corrected by multiple hypothesis testing to give the Q-value by the Benjamini-Hochberg method. **Functional Annotation is adapted from Database for Annotation, Visualization and Integrated Discovery (DAVID).

#### Supplementary Materials

#### Supplementary Materials

**Supplemental Table 12.** Top 50 Upregulated Genes in Inflammatory LS *versus* HC.

| Gen | Protein Encoded | logFC | Q-Value* | Functional Annotation** |
|---|---|---|---|---|
| *PLA2G2D* | Phospholipase A2 group IID | 7.895575 | 0.006136 | CD4-positive, CD25-positive, alpha-beta regulatory T cell differentiation, regulation of acute inflammatory response to antigenic stimulus, phospholipid metabolic process, inflammatory response, lipid catabolic process, negative regulation of T cell proliferation, phosphatidylethanolamine metabolic process, phosphatidylcholine metabolic process, phosphatidylglycerol metabolic process, and arachidonate secretion. |
| *CXCL10* | C-X-C motif chemokine ligand 10 | 7.263021 | 0.044945 | Chemotaxis, inflammatory response, signal transduction, cell surface receptor signaling pathway, G protein-coupled receptor signaling pathway, adenylate cyclase-activating G protein-coupled receptor signaling pathway, cell-cell signaling, muscle organ development, blood circulation, positive regulation of cell population proliferation, response to gamma radiation, T cell chemotaxis, regulation of T cell chemotaxis, response to auditory stimulus, negative regulation of angiogenesis, neutrophil chemotaxis, killing of cells of another organism, response to vitamin D, cellular response to heat, endothelial cell activation, regulation of cell population proliferation, regulation of apoptotic process, negative regulation of myoblast differentiation, positive regulation of transcription by RNA polymerase II, positive chemotaxis, positive regulation of release of sequestered calcium ion into cytosol, antimicrobial humoral immune response mediated by antimicrobial peptide, chemokine-mediated signaling pathway, cellular response to lipopolysaccharide, positive regulation of monocyte chemotaxis, cellular response to interleukin-17, cellular response to virus, antiviral innate immune response, regulation of endothelial tube morphogenesis, negative regulation of myoblast fusion, and positive regulation of T cell migration. |
| *CCL3L1* | C-C motif chemokine ligand 3 like 3 | 6.981265 | 0.003968 | Inflammatory response, immune response, negative regulation of cell population proliferation, positive regulation of cell migration, eosinophil chemotaxis, cell chemotaxis, antimicrobial humoral immune response mediated by antimicrobial peptide, and chemokine-mediated signaling pathway. |
| *FCRL3* | Fc receptor like 3 | 6.879306 | 0.014386 | Negative regulation of immunoglobulin production, immune response, cell surface receptor signaling pathway, single fertilization, positive regulation of B cell proliferation, regulation of toll-like receptor 9 signaling pathway, positive regulation of MAPK cascade, regulation of B cell differentiation, negative regulation of B cell receptor signaling pathway, regulation of B cell activation, and regulation of calcium ion import. |
| *ADAMDEC1* | ADAM like decysin 1 | 6.869514 | 0.014628 | Proteolysis, immune response, negative regulation of cell adhesion, and integrin-mediated signaling pathway. |
| *CXCL11* | C-X-C motif chemokine ligand 11 | 6.838779 | 0.00368 | Chemotaxis, inflammatory response, signal transduction, adenylate cyclase-activating G protein-coupled receptor signaling pathway, cell-cell signaling, T cell chemotaxis, neutrophil chemotaxis, killing of cells of another organism, regulation of cell population proliferation, positive regulation of release of sequestered calcium ion into cytosol, antimicrobial humoral immune response mediated by antimicrobial peptide, chemokine-mediated signaling pathway, and cellular response to lipopolysaccharide. |
| *LAG3* | Lymphocyte activating 3 | 6.669933 | $5.92 \times 10^{-5}$ | Adaptive immune response, plasmacytoid dendritic cell activation, cell surface receptor signaling pathway, negative regulation of interleukin-2 production, T cell activation, natural killer cell mediated cytotoxicity, innate immune response, negative regulation of regulatory T cell differentiation, positive regulation of natural killer cell mediated cytotoxicity, and regulation of immune response. |
| *TTC24* | Tetratricopeptide repeat domain 24 | 6.517082 | 0.001489 | Unknown function. |
| *GZMA* | granzyme A | 6.496906 | 0.01084 | Apoptotic process, immune response, response to bacterium, killing of cells of another organism, negative regulation of endodeoxyribonuclease activity, positive regulation of apoptotic process, negative regulation of DNA binding, negative regulation of oxidoreductase activity, proteolysis involved in protein catabolic process, protein maturation, pyroptotic inflammatory response, granzyme-mediated programmed cell death signaling pathway, and cytotoxic T cell pyroptotic cell death. |
| *SLC12A3* | Solute carrier family 12 member 3 | 6.412847 | 0.00214 | Response to dietary excess, monoatomic ion transport, sodium ion transport, cell volume homeostasis, signal transduction, sodium ion transmembrane transport, chloride ion homeostasis, potassium ion homeostasis, sodium ion homeostasis, renal sodium ion absorption, chloride |

| | | | | |
|---|---|---|---|---|
| | | | | transmembrane transport, response to aldosterone, and potassium ion import across plasma membrane. |
| *IGHV3-30* | Immunoglobulin heavy variable 3-30 | 6.403192 | 0.027084 | Immune response and immunoglobulin mediated immune response. |
| *IDO1* | Indoleamine 2,3-dioxygenase 1 | 6.282943 | 0.009929 | Immune system process, positive regulation of T cell tolerance induction, positive regulation of chronic inflammatory response, positive regulation of type 2 immune response, tryptophan catabolic process, inflammatory response, female pregnancy, tryptophan catabolic process to kynurenine, quinolinate biosynthetic process, response to lipopolysaccharide, negative regulation of interleukin-10 production, positive regulation of interleukin-12 production, multicellular organismal response to stress, kynurenic acid biosynthetic process, 'de novo' NAD biosynthetic process from tryptophan, swimming behavior, T cell proliferation, negative regulation of T cell proliferation, positive regulation of apoptotic process, negative regulation of activated T cell proliferation, negative regulation of T cell apoptotic process, and positive regulation of T cell apoptotic process. |
| *PACSIN1* | Protein kinase C and casein kinase substrate in neurons 1 | 6.259739 | 0.000368 | Cytoskeleton organization, actin filament organization, regulation of endocytosis, negative regulation of endocytosis, synaptic vesicle endocytosis, neuron projection morphogenesis, protein localization to membrane, protein localization to plasma membrane, plasma membrane tubulation, and positive regulation of dendrite development. |
| *GZMK* | Granzyme K | 6.241519 | 0.015403 | Proteolysis, protein maturation, granzyme-mediated programmed cell death signaling pathway. |
| *GZMB* | Granzyme B | 6.176459 | 0.01723 | Proteolysis, apoptotic process, negative regulation of translation, killing of cells of another organism, natural killer cell mediated cytotoxicity, proteolysis involved in protein catabolic process, protein maturation, pyroptotic inflammatory response, granzyme-mediated programmed cell death signaling pathway, and positive regulation of establishment of protein localization to mitochondrion. |
| *TIGIT* | T cell immunoreceptor with Ig and ITIM domains | 6.060183 | 0.004504 | Negative regulation of interleukin-12 production, positive regulation of interleukin-10 production, negative regulation of natural killer cell mediated cytotoxicity, and negative regulation of T cell activation. |
| *TRAJ6* | T cell receptor alpha joining 6 | 6.058536 | 0.006312 | Unknown function. |
| *FCRL6* | Fc receptor like 6 | 6.039736 | 0.001181 | Immune response and cell surface receptor signaling pathway. |
| *UBASH3A* | Ubiquitin associated and SH3 domain containing A | 5.880184 | 0.016781 | Regulation of cytokine production, and negative regulation of T cell receptor signaling pathway. |
| *PLA1A* | Phospholipase A1 member A | 5.863125 | 0.009863 | Lipid metabolic process, phosphatidylserine metabolic process, lipid catabolic process, and phosphatidylserine acyl-chain remodeling. |
| *EOMES* | Eomesodermin | 5.761123 | 0.026262 | Negative regulation of transcription by RNA polymerase II, endoderm formation, mesoderm formation, endodermal cell fate specification, neuron migration, trophectodermal cell differentiation, adaptive immune response, CD8-positive, alpha-beta T cell differentiation involved in immune response, chromatin remodeling, regulation of transcription by RNA polymerase II, brain development, cardioblast differentiation, gene expression, stem cell population maintenance, olfactory bulb development, cerebral cortex regionalization, cerebral cortex neuron differentiation, skeletal muscle cell differentiation, negative regulation of DNA-binding transcription factor activity, positive regulation of DNA-templated transcription, positive regulation of transcription by RNA polymerase II, mesendoderm development, astrocyte differentiation, cell differentiation involved in embryonic placenta development, and mesodermal to mesenchymal transition involved in gastrulation. |
| *TDO2* | Tryptophan 2,3-dioxygenase | 5.742643 | 0.001913 | Tryptophan metabolic process, tryptophan catabolic process to kynurenine, tryptophan catabolic process to acetyl-CoA, protein homotetramerization, and response to nitroglycerin. |
| *TRAJ47* | T cell receptor alpha joining 47 | 5.646062 | 0.001128 | Unknown function. |
| *CRTAM* | Cytotoxic and regulatory T cell molecule | 5.638861 | 0.011894 | Establishment of T cell polarity, positive regulation of cytokine production, adaptive immune response, detection of tumor cell, positive regulation of natural killer cell mediated cytotoxicity directed against tumor cell target, heterophilic cell-cell adhesion via plasma membrane cell adhesion molecules, cell recognition, positive regulation of type II interferon production, regulation of T cell differentiation, positive regulation of natural killer cell mediated cytotoxicity, negative regulation of activated T cell proliferation, regulation of T cell activation, detection of |

| | | | | |
|---|---|---|---|---|
| | | | | stimulus, lymphocyte migration into lymphoid organs, regulation of CD8-positive, and alpha-beta T cell activation. |
| *NUGGC* | Nuclear GTPase, germinal center associated | 5.614293 | 0.013431 | Regulation of nuclear cell cycle DNA replication, negative regulation of apoptotic process, and cellular response to lipopolysaccharide. |
| *CNGB1* | Cyclic nucleotide gated channel subunit beta 1 | 5.595467 | 0.008022 | Retina homeostasis, monoatomic cation transport, potassium ion transport, sodium ion transport, calcium ion transport, visual perception, phototransduction, sensory perception of smell, positive regulation of gene expression, olfactory nerve maturation, protein localization to organelle, monoatomic ion transmembrane transport, sodium ion transmembrane transport, photoreceptor cell outer segment organization, photoreceptor cell maintenance, detection of light stimulus involved in visual perception, detection of chemical stimulus involved in sensory perception of smell, regulation of cytosolic calcium ion concentration, membrane depolarization, calcium ion transmembrane transport, monoatomic cation transmembrane transport, ion channel modulating, G protein-coupled receptor signaling pathway, and response to odorant. |
| *C11orf42* | Chromosome 11 open reading frame 42 | 5.595181 | 0.000845 | Unknown function. |
| *IGHJ5* | Immunoglobulin heavy joining 5 | 5.580898 | 0.017772 | Unknown function. |
| *TRAJ17* | T cell receptor alpha joining 17 | 5.524647 | 0.002044 | Unknown function. |
| *CETP* | Cholesteryl ester transfer protein | 5.518989 | 0.00107 | Triglyceride metabolic process, lipid transport, cholesterol metabolic process, negative regulation of macrophage derived foam cell differentiation, regulation of cholesterol efflux, phospholipid transport, cholesterol transport, positive regulation of cholesterol transport, triglyceride transport, very-low-density lipoprotein particle remodeling, low-density lipoprotein particle remodeling, high-density lipoprotein particle remodeling, cholesterol homeostasis, reverse cholesterol transport, phosphatidylcholine metabolic process, lipid homeostasis, phospholipid homeostasis, triglyceride homeostasis, and positive regulation of phospholipid transport. |
| *TRAJ35* | T cell receptor alpha joining 35 (non-functional) | 5.514488 | 0.010977 | Unknown function. |
| *IL4I1* | Interleukin 4 induced 1 | 5.495 | 0.003679 | Adaptive immune response, regulation of adaptive immune response, negative regulation of T cell mediated immune response to tumor cell, L-phenylalanine catabolic process, tryptophan catabolic process, tyrosine catabolic process, amino acid catabolic process, tryptophan catabolic process to indole-3-acetate, negative regulation of T cell proliferation, regulation of B cell differentiation, positive regulation of regulatory T cell differentiation, and positive regulation of transcription by RNA polymerase II, and negative regulation of T cell activation. |
| *F2* | Coagulation factor II, thrombin | 5.489594 | 0.004424 | Positive regulation of protein phosphorylation, proteolysis, acute-phase response, cell surface receptor signaling pathway, G protein-coupled receptor signaling pathway, blood coagulation, positive regulation of cell population proliferation, regulation of cell shape, response to wounding, negative regulation of platelet activation, platelet activation, regulation of blood coagulation, positive regulation of blood coagulation, negative regulation of blood coagulation, positive regulation of cell growth, positive regulation of insulin secretion, positive regulation of collagen biosynthetic process, fibrinolysis, negative regulation of proteolysis, positive regulation of receptor signaling pathway via JAK-STAT, negative regulation of astrocyte differentiation, tissue remodeling, positive regulation of release of sequestered calcium ion into cytosol, regulation of cytosolic calcium ion concentration, cytolysis by host of symbiont cells, positive regulation of phosphatidylinositol 3-kinase/protein kinase B signal transduction, negative regulation of fibrinolysis, antimicrobial humoral immune response mediated by antimicrobial peptide, thrombin-activated receptor signaling pathway, neutrophil-mediated killing of gram-negative bacterium, positive regulation of lipid kinase activity, negative regulation of cytokine production involved in inflammatory response, positive regulation of protein localization to nucleus, positive regulation of phospholipase C-activating G protein-coupled receptor signaling pathway, ligand-gated ion channel signaling pathway, and positive regulation of reactive oxygen species metabolic process. |

| | | | | |
|---|---|---|---|---|
| *GZMH* | Granzyme H | 5.460792 | 0.019417 | Proteolysis, apoptotic process, killing of cells of another organism, and protein maturation. |
| *SI* | Sucrase-isomaltase | 5.380332 | 0.018882 | Sucrose catabolic process, and polysaccharide digestion. |
| *CD27* | CD27 molecule | 5.360276 | 0.016781 | Cell surface receptor signaling pathway, immunoglobulin mediated immune response, T cell activation, negative regulation of apoptotic process, response to ethanol, positive regulation of B cell differentiation, positive regulation of T cell differentiation, positive regulation of JNK cascade, negative regulation of T cell apoptotic process, adaptive immune memory response involving T cells and B cells, extrinsic apoptotic signaling pathway, CD27 signaling pathway, and positive regulation of non-canonical NF-kappaB signal transduction. |
| *MYH7* | Myosin heavy chain 7 | 5.319966 | 0.041248 | Regulation of the force of heart contraction, regulation of heart rate, skeletal muscle contraction, muscle contraction, striated muscle contraction, adult heart development, regulation of the force of skeletal muscle contraction, transition between fast and slow fiber, cardiac muscle hypertrophy in response to stress, muscle filament sliding, regulation of slow-twitch skeletal muscle fiber contraction, sarcomere organization, ATP metabolic process, ventricular cardiac muscle tissue morphogenesis, and cardiac muscle contraction. |
| *ARSL* | Arylsulfatase L | 5.318337 | 0.001809 | Skeletal system development. |
| *CERKL* | Ceramide kinase like | 5.308302 | 0.014313 | Sphingolipid metabolic process, sphingolipid biosynthetic process, and negative regulation of apoptotic process. |
| *FHAD1* | Forkhead associated phosphopeptide binding domain 1 | 5.180048 | 0.017544 | Unknown function. |
| *RIMBP3* | RIMS binding protein 3 | 5.173931 | 0.01694 | Spermatid development and fertilization. |
| *TRAJ7* | T cell receptor alpha joining 7 | 5.118298 | 0.003849 | Unknown function. |
| *ERN2* | endoplasmic reticulum to nucleus signaling 2 | 5.110908 | 0.012569 | Chromatin remodeling, mRNA processing, protein phosphorylation, regulation of gene expression, rRNA catabolic process, apoptotic chromosome condensation, endoplasmic reticulum unfolded protein response, response to endoplasmic reticulum stress, IRE1-mediated unfolded protein response, negative regulation of DNA-templated transcription, intrinsic apoptotic signaling pathway in response to endoplasmic reticulum stress, and regulation of primary metabolic process. |
| *NEFM* | Neurofilament medium chain | 5.109341 | 0.021833 | Neurofilament bundle assembly. |
| *AIM2* | Absent in melanoma 2 | 5.072051 | 0.010187 | Activation of innate immune response, pattern recognition receptor signaling pathway, positive regulation of defense response to virus by host, immune response, DNA damage response, brain development, negative regulation of NF-kappaB transcription factor activity, positive regulation of interleukin-1 beta production, tumor necrosis factor-mediated signaling pathway, cellular response to interferon-beta, T cell homeostasis, innate immune response, positive regulation of inflammatory response, regulation of behavior, positive regulation of NF-kappaB transcription factor activity, defense response to virus, negative regulation of phosphatidylinositol 3-kinase/protein kinase B signal transduction, neuron cellular homeostasis, pyroptotic inflammatory response, cellular response to xenobiotic stimulus, AIM2 inflammasome complex assembly, pyroptosome complex assembly, and positive regulation of cysteine-type endopeptidase activity. |
| *SLCO5A1* | Solute carrier organic anion transporter family member 5A1 | 5.069789 | 0.009285 | Sodium-independent organic anion transport, and transmembrane transport. |
| *KLRC2* | Killer cell lectin like receptor C2 | 5.048911 | 0.006162 | Stimulatory C-type lectin receptor signaling pathway, natural killer cell mediated immunity, adaptive immune response, cellular defense response, signal transduction, regulation of natural killer cell activation, positive regulation of natural killer cell degranulation, positive regulation of natural killer cell mediated cytotoxicity. |
| *TRAJ41* | T cell receptor alpha joining 41 | 5.042022 | 0.007509 | Unknown function. |
| *HSF5* | Heat shock transcription factor 5 | 5.022644 | 0.011834 | Regulation of transcription by RNA polymerase II, male meiotic nuclear division, spermatogenesis, cell differentiation, negative regulation of DNA-templated transcription, and positive regulation of DNA-templated transcription. |

#### Supplementary Materials

| | | | | |
|---|---|---|---|---|
| *MZB1* | Marginal zone B and B1 cell specific protein | 5.016251 | 0.02579 | Positive regulation of immunoglobulin production, apoptotic process, positive regulation of cell population proliferation, regulation of B cell proliferation, integrin activation, regulation of cell population proliferation, and regulation of insulin receptor signaling pathway. |

*P-values were corrected by multiple hypothesis testing to give the Q-value by the Benjamini-Hochberg method. **Functional Annotation** is adapted from Database for Annotation, Visualization and Integrated Discovery (DAVID).

#### Supplementary Materials

**Supplemental Table 13.** Top 50 Downregulated Genes in Inflammatory LS *versus* HC.

| Gen | Protein Encoded | logFC | Q-Value* | Functional Annotation** |
|---|---|---|---|---|
| *FOSB* | FosB proto-oncogene, AP-1 transcription factor subunit | -4.63952 | 0.012696 | Negative regulation of transcription by RNA polymerase II, response to amphetamine, regulation of transcription by RNA polymerase II, transcription by RNA polymerase II, female pregnancy, response to xenobiotic stimulus, response to mechanical stimulus, response to progesterone, cellular response to hormone stimulus, response to morphine, response to ethanol, positive regulation of transcription by RNA polymerase II, behavioral response to cocaine, response to corticosterone, response to cAMP, and cellular response to calcium ion. |
| *MATN4* | Matrilin 4 | -4.43286 | 0.006142 | Extracellular matrix organization. |
| *SERTM1* | Serine rich and transmembrane domain containing 1 | -4.39376 | 0.002618 | Membrane and intracellular membrane-bounded organelle. |
| *TNMD* | Tenomodulin | -4.20675 | 0.007601 | Endothelial cell morphogenesis, negative regulation of endothelial cell proliferation, and negative regulation of angiogenesis. |
| *FOS* | Fos proto-oncogene, AP-1 transcription factor subunit | -4.16386 | 0.000792 | Conditioned taste aversion, neural retina development, regulation of transcription by RNA polymerase II, transcription by RNA polymerase II, inflammatory response, transforming growth factor beta receptor signaling pathway, nervous system development, female pregnancy, response to xenobiotic stimulus, response to light stimulus, response to gravity, response to toxic substance, response to activity, skeletal muscle cell proliferation, cerebral cortex development, neuron differentiation, osteoclast differentiation, response to lipopolysaccharide, response to progesterone, response to insulin, cellular response to zinc ion starvation, cellular response to reactive oxygen species, response to immobilization stress, skeletal muscle cell differentiation, response to muscle stretch, response to ethanol, positive regulation of osteoclast differentiation, positive regulation of DNA-templated transcription, positive regulation of transcription by RNA polymerase II, response to corticosterone, myoblast proliferation, response to cAMP, SMAD protein signal transduction, cellular response to cadmium ion, cellular response to calcium ion, cellular response to tumor necrosis factor, cellular response to epidermal growth factor stimulus, cellular response to parathyroid hormone stimulus, cellular response to hypoxia, cellular response to transforming growth factor beta stimulus, medium-term memory, integrated stress response signaling, positive regulation of miRNA transcription, mononuclear cell differentiation, cellular response to phorbol 13-acetate 12-myristate, and cellular response to prolactin. |
| *CLDN11* | Claudin 11 | -4.12274 | 0.000792 | Cell adhesion, spermatogenesis, axon ensheathment, calcium-independent cell-cell adhesion via plasma membrane cell-adhesion molecules, bicellular tight junction assembly, and tight junction assembly. |
| *AHSP* | Alpha hemoglobin stabilizing protein | -4.02282 | 0.019958 | Protein folding, hemoglobin metabolic process, hemopoiesis, erythrocyte differentiation, and protein stabilization. |
| *SERPINA5* | Serpin family A member 5 | -3.94162 | 0.004726 | Acrosomal membrane, extracellular region, extracellular space, external side of plasma membrane, membrane, platelet alpha granule, platelet dense tubular network, protein-containing complex, protein C inhibitor-TMPRSS7 complex, protein C inhibitor-TMPRSS11E complex, protein C inhibitor-PLAT complex, protein C inhibitor-PLAU complex, protein C inhibitor-thrombin complex, protein C inhibitor-KLK3 complex, protein C inhibitor-plasma kallikrein complex, collagen-containing extracellular matrix, extracellular exosome, protein C inhibitor-coagulation factor V complex, protein C inhibitor-coagulation factor Xa complex, and protein C inhibitor-coagulation factor XI complex, |
| *WIF1* | WNT inhibitory factor 1 | -3.93192 | 0.016926 | Blood vessel development, signal transduction, determination of left/right symmetry, heart development, anterior/posterior pattern specification, Wnt signaling pathway, negative regulation of Wnt signaling pathway, nodal signaling pathway, and positive regulation of fat cell differentiation. |
| *SMIM24* | Small integral membrane protein 24 | -3.87699 | 0.015436 | Biological process. |
| *LY6K* | Lymphocyte antigen 6 family member K | -3.71016 | 0.002422 | Binding of sperm to zona pellucida. |
| *CDH4* | Cadherin 4 | -3.64389 | 0.007028 | Cell morphogenesis, cell-cell junction assembly, cell adhesion, homophilic cell adhesion via plasma membrane adhesion molecules, heterophilic cell-cell adhesion via plasma membrane cell adhesion molecules, axon guidance, calcium-dependent cell-cell adhesion via plasma membrane cell |

| | | | | |
|---|---|---|---|---|
| | | | | adhesion molecules, cell migration, adherens junction organization, cell-cell adhesion mediated by cadherin, positive regulation of axon extension, and axon extension. |
| *BEX5* | Brain expressed X-linked 5 | -3.61348 | 0.006136 | Signal transduction. |
| *RPLP1P6* | Ribosomal protein lateral stalk subunit P1 pseudogene 6 [ | -3.59322 | 6.39X 10-5 | Unknown function. |
| *MIR494* | MicroRNA 494 | -3.52041 | 0.033773 | Negative regulation of gene expression, miRNA-mediated post-transcriptional gene silencing, negative regulation of fibroblast proliferation, negative regulation of cell migration involved in sprouting angiogenesis, negative regulation of blood vessel endothelial cell proliferation involved in sprouting angiogenesis, and negative regulation of artery morphogenesis. |
| *VEGFD* | Vascular endothelial growth factor D | -3.51692 | 0.038701 | Response to hypoxia, positive regulation of protein phosphorylation, positive regulation of endothelial cell proliferation, sprouting angiogenesis, positive regulation of cell population proliferation, response to bacterium, vascular endothelial growth factor signaling pathway, positive regulation of angiogenesis, vascular endothelial growth factor receptor signaling pathway, fibroblast proliferation, positive chemotaxis, induction of positive chemotaxis, positive regulation of cell division, positive regulation of mast cell chemotaxis, and dopaminergic neuron differentiation. |
| *CES1P1* | Carboxylesterase 1 pseudogene 1 | -3.51408 | 0.037683 | Anatomical structure morphogenesis and lipid catabolic process. |
| *ENSG00000 286555* | Novel transcript, antisense to ARAP1 | -3.45297 | 0.007057 | *Pseudogene.* |
| *OR51B6* | Olfactory receptor family 51 subfamily B member 6 | -3.42701 | 0.013666 | G protein-coupled receptor signaling pathway, sensory perception of smell, detection of chemical stimulus involved in sensory perception of smell. |
| *ENSG00000 278001* | Family with sequence similarity 27, member D1 (FAM27D1) pseudogene | -3.33585 | 0.010489 | *Pseudogene.* |
| *ENSG00000 250770* | Tetraspanin 11 (TSPAN11) pseudogene | -3.33023 | 0.017989 | *Pseudogene.* |
| *LYVE1* | Lymphatic vessel endothelial hyaluronan receptor 1 | -3.32458 | 0.00864 | Positive regulation of cellular extravasation, receptor-mediated endocytosis, cell adhesion, cell-matrix adhesion, response to wounding, anatomical structure morphogenesis, hyaluronan catabolic process. |
| *ENSG00000 249593* | Novel transcript | -3.30891 | 0.016781 | *Pseudogene.* |
| *EGR1* | Early growth response 1 | -3.20557 | $9.06 \times 10^{-5}$ | Negative regulation of transcription by RNA polymerase II, response to hypoxia, response to ischemia, regulation of transcription by RNA polymerase II, response to glucose, positive regulation of gene expression, T cell differentiation, BMP signaling pathway, positive regulation of chemokine production, positive regulation of interleukin-1 beta production, response to insulin, circadian regulation of gene expression, regulation of protein sumoylation, skeletal muscle cell differentiation, regulation of neuron apoptotic process, positive regulation of gene expression via chromosomal CpG island demethylation, estrous cycle, locomotor rhythm, positive regulation of DNA-templated transcription, positive regulation of transcription by RNA polymerase II, positive regulation of hormone biosynthetic process, rhythmic process, circadian temperature homeostasis, interleukin-1-mediated signaling pathway, cellular response to gamma radiation, cellular response to heparin, cellular response to mycophenolic acid, glomerular mesangial cell proliferation, positive regulation of glomerular metanephric mesangial cell proliferation, negative regulation of canonical Wnt signaling pathway, cellular response to interleukin-8, positive regulation of post-translational protein modification, positive regulation of miRNA transcription, and regulation of progesterone biosynthetic process. |
| *ENSG00000 262202* | Novel transcript | -3.19739 | 0.018015 | *Pseudogene.* |
| *WBP11P3* | WBP11 pseudogene 3 | -3.17323 | 0.014928 | Unknown function. |

#### Supplementary Materials

| | | | | |
|---|---|---|---|---|
| *GREM2* | Gremlin 2, DAN family BMP antagonist | -3.13571 | 0.005025 | Animal organ morphogenesis, embryonic body morphogenesis, cytokine-mediated signaling pathway, sequestering of BMP from receptor via BMP binding, determination of dorsal identity, regulation of cytokine activity, |
| *PCOLCE2* | Procollagen C-endopeptidase enhancer 2 | -3.09553 | 0.002023 | Cellular response to leukemia inhibitory factor. |
| *BMP3* | Bone morphogenetic protein 3 | -3.07488 | 0.027025 | Skeletal system development, osteoblast differentiation, signal transduction, cell-cell signaling, negative regulation of ossification, cartilage development. |
| *SCARA5* | Scavenger receptor class A member 5 | -3.04973 | 0.014775 | Intracellular iron ion homeostasis, endocytosis, cellular response to heat, iron ion transmembrane transport, and protein homotrimerization. |
| *ASS1P9* | Argininosuccinate synthetase 1 pseudogene 9 | -3.02976 | 0.021833 | Unknown function. |
| *SCUBE1* | Signal peptide, CUB domain and EGF like domain containing 1 | -3.02151 | 0.010977 | Inflammatory response, signal transduction, adult heart development, blood coagulation, post-embryonic development, endothelial cell differentiation, positive regulation of smoothened signaling pathway, |
| *AQP7* | Aquaporin 7 | -3.01047 | 0.003968 | Water transport, polyol transmembrane transport, glycerol transmembrane transport, urea transmembrane transport. |
| *DUSP1* | Dual specificity phosphatase 1 | -3.00625 | 0.000783 | Endoderm formation, chromatin remodeling, regulation of transcription by RNA polymerase II, negative regulation of cell adhesion, signal transduction, negative regulation of cell population proliferation, peptidyl-tyrosine dephosphorylation, peptidyl-threonine dephosphorylation, negative regulation of MAP kinase activity, negative regulation of MAPK cascade, negative regulation of meiotic cell cycle, peptidyl-serine dephosphorylation, negative regulation of monocyte chemotaxis, regulation of mitotic cell cycle spindle assembly checkpoint, negative regulation of p38MAPK cascade, and cellular response to chemokine. |
| *ENSG00000 167046* | Novel transcript, antisense to SLCO4A1 | -2.93004 | 0.046959 | *Pseudogene*. |
| *OR5P1P* | Olfactory receptor family 5 subfamily P member 1 pseudogene | -2.92295 | 0.036897 | G protein-coupled receptor signaling pathway. |
| *CLEC3B* | C-type lectin domain family 3 member B | -2.91996 | 0.001128 | Ossification, positive regulation of plasminogen activation, and bone mineralization. |
| *ATP5MC1P6* | ATP synthase membrane subunit c locus 1 pseudogene 6 | -2.88631 | 0.026327 | Unknown function. |
| *PRRT4* | Proline rich transmembrane protein 4 | -2.88129 | 0.017989 | Membrane. |
| *HAS1* | hyaluronan synthase 1 | -2.85536 | 0.037683 | Polysaccharide biosynthetic process, glycosaminoglycan biosynthetic process, cell adhesion, negative regulation of fibroblast migration, hyaluronan biosynthetic process, cellular response to platelet-derived growth factor stimulus, and extracellular matrix assembly. |
| *PNMA2* | PNMA family member 2 | -2.83847 | 0.033331 | Positive regulation of apoptotic process. |
| *PBOV1* | Prostate and breast cancer overexpressed 1 | -2.81915 | 0.038712 | Nucleoplasm, cytosol and centriolar satellite. |
| *PDGFRL* | Platelet derived growth factor receptor like | -2.73158 | 0.001181 | G protein-coupled receptor signaling pathway and platelet-derived growth factor receptor-beta signaling pathway. |
| *BUB3P1* | BUB3 mitotic checkpoint protein pseudogene 1 | -2.71631 | 0.042752 | Unknown function. |
| *RPL32P10* | Ribosomal protein L32 pseudogene 10 | -2.70023 | 0.028425 | Unknown function. |
| *CES1* | Carboxylesterase 1 | -2.67816 | 0.002382 | Anatomical structure morphogenesis and lipid catabolic process. |
| *DPPA2P4* | Developmental pluripotency associated 2 pseudogene 4 | -2.64047 | 0.030065 | Unknown function. |

#### Supplementary Materials

| | | | | |
|---|---|---|---|---|
| *MT1G* | Metallothionein 1G | -2.63974 | 0.026241 | Intracellular zinc ion homeostasis, detoxification of copper ion, monocyte differentiation, cellular response to vascular endothelial growth factor stimulus, monocyte activation, negative regulation of growth, cellular response to cadmium ion, cellular response to copper ion, and cellular response to zinc ion. |
| *CYP4Z2P* | Cytochrome P450 family 4 subfamily Z member 2, pseudogene | -2.62887 | 0.025844 | Membrane. |
| *CYP4B1* | Cytochrome P450 family 4 subfamily B member 1 | -2.62576 | 0.040109 | Fatty acid metabolic process, and biphenyl metabolic process. |

*P-values were corrected by multiple hypothesis testing to give the Q-value by the Benjamini-Hochberg method. **Functional Annotation is adapted from Database for Annotation, Visualization and Integrated Discovery (DAVID).

#### Supplementary Materials

**Supplemental Table 14.** Top 50 Upregulated Genes in Mixed LS *versus* HC.

| Gen | Protein Encoded | logFC | Q-Value* | Functional Annotation** |
|---|---|---|---|---|
| *PLA2G2D* | Phospholipase A2 group IID | 9.859328 | 0.002914 | CD4-positive, CD25-positive, alpha-beta regulatory T cell differentiation, regulation of acute inflammatory response to antigenic stimulus, phospholipid metabolic process, inflammatory response, lipid catabolic process, negative regulation of T cell proliferation, phosphatidylethanolamine metabolic process, phosphatidylcholine metabolic process, phosphatidylglycerol metabolic process, and arachidonate secretion. |
| *CCL3L1* | C-C motif chemokine ligand 3 like 3 | 8.885192 | 0.001661 | Inflammatory response, immune response, negative regulation of cell population proliferation, positive regulation of cell migration, eosinophil chemotaxis, cell chemotaxis, antimicrobial humoral immune response mediated by antimicrobial peptide, and chemokine-mediated signaling pathway. |
| *ADAMDEC1* | ADAM like decysin 1 | 8.50145 | 0.00819 | Proteolysis, immune response, negative regulation of cell adhesion, and integrin-mediated signaling pathway. |
| *GZMK* | Granzyme K | 8.414446 | 0.004406 | Proteolysis, protein maturation, and granzyme-mediated programmed cell death signaling pathway. |
| *CXCL10* | C-X-C motif chemokine ligand 10 | 8.208479 | 0.046377 | Chemotaxis, inflammatory response, signal transduction, cell surface receptor signaling pathway, G protein-coupled receptor signaling pathway, adenylate cyclase-activating G protein-coupled receptor signaling pathway, cell-cell signaling, muscle organ development, blood circulation, positive regulation of cell population proliferation, response to gamma radiation, T cell chemotaxis, regulation of T cell chemotaxis, response to auditory stimulus, negative regulation of angiogenesis, neutrophil chemotaxis, killing of cells of another organism, response to vitamin D, cellular response to heat, endothelial cell activation, regulation of cell population proliferation, regulation of apoptotic process, negative regulation of myoblast differentiation, positive regulation of transcription by RNA polymerase II, positive chemotaxis, positive regulation of release of sequestered calcium ion into cytosol, antimicrobial humoral immune response mediated by antimicrobial peptide, chemokine-mediated signaling pathway, cellular response to lipopolysaccharide, positive regulation of monocyte chemotaxis, cellular response to interleukin-17, cellular response to virus, antiviral innate immune response, regulation of endothelial tube morphogenesis, negative regulation of myoblast fusion, and positive regulation of T cell migration. |
| *MS4A1* | Membrane spanning 4-domains A1 | 8.161711 | 0.031671 | Store-operated calcium entry, humoral immune response, cell surface receptor signaling pathway, response to bacterium, B cell differentiation, B cell proliferation, B cell activation, B cell receptor signaling pathway, protein tetramerization, calcium ion import into cytosol, and positive regulation of calcium ion import across plasma membrane. |
| *GZMA* | Granzyme A | 8.13217 | 0.005367 | Apoptotic process, immune response, response to bacterium, killing of cells of another organism, negative regulation of endodeoxyribonuclease activity, positive regulation of apoptotic process, negative regulation of DNA binding, negative regulation of oxidoreductase activity, proteolysis involved in protein catabolic process, protein maturation, pyroptotic inflammatory response, granzyme-mediated programmed cell death signaling pathway, and cytotoxic T cell pyroptotic cell death. |
| *CXCL11* | C-X-C motif chemokine ligand 11 | 8.083087 | 0.003375 | Chemotaxis, inflammatory response, signal transduction, adenylate cyclase-activating G protein-coupled receptor signaling pathway, cell-cell signaling, T cell chemotaxis, neutrophil chemotaxis, killing of cells of another organism, regulation of cell population proliferation, positive regulation of release of sequestered calcium ion into cytosol, antimicrobial humoral immune response mediated by antimicrobial peptide, chemokine-mediated signaling pathway, and cellular response to lipopolysaccharide. |
| *TTC24* | Tetratricopeptide repeat domain 24 | 8.053313 | 0.001143 | Unknown function. |
| *FCRL3* | Fc receptor like 3 | 7.904898 | 0.014335 | Negative regulation of immunoglobulin production, immune response, cell surface receptor signaling pathway, single fertilization, positive regulation of B cell proliferation, regulation of toll-like receptor 9 signaling pathway, positive regulation of MAPK cascade, regulation of B cell differentiation, negative regulation of B cell receptor signaling pathway, regulation of B cell activation, and regulation of calcium ion import. |

#### Supplementary Materials

| | | | | |
|---|---|---|---|---|
| *IGHGP* | Immunoglobulin heavy constant gamma P (non-functional) | 7.782268 | 0.049154 | Unknown function. |
| *CRTAM* | Cytotoxic and regulatory T cell molecule | 7.658448 | 0.002969 | Establishment of T cell polarity, positive regulation of cytokine production, adaptive immune response, detection of tumor cell, positive regulation of natural killer cell mediated cytotoxicity directed against tumor cell target, heterophilic cell-cell adhesion via plasma membrane cell adhesion molecules, cell recognition, positive regulation of type II interferon production, regulation of T cell differentiation, positive regulation of natural killer cell mediated cytotoxicity, negative regulation of activated T cell proliferation, regulation of T cell activation, detection of stimulus, lymphocyte migration into lymphoid organs, regulation of CD8-positive, and alpha-beta T cell activation. |
| *FCRL6* | Fc receptor like 6 | 7.566648 | 0.000682 | Immune response, cell surface receptor signaling pathway. |
| *IDO1* | Indoleamine 2,3-dioxygenase 1 | 7.566347 | 0.006666 | Immune system process, positive regulation of T cell tolerance induction, positive regulation of chronic inflammatory response, positive regulation of type 2 immune response, tryptophan catabolic process, inflammatory response, female pregnancy, tryptophan catabolic process to kynurenine, quinolinate biosynthetic process, response to lipopolysaccharide, negative regulation of interleukin-10 production, positive regulation of interleukin-12 production, multicellular organismal response to stress, kynurenic acid biosynthetic process, 'de novo' NAD biosynthetic process from tryptophan, swimming behavior, T cell proliferation, negative regulation of T cell proliferation, positive regulation of apoptotic process, negative regulation of activated T cell proliferation, negative regulation of T cell apoptotic process, positive regulation of T cell apoptotic process. |
| *IGLV6-57* | Immunoglobulin lambda variable 6-57 | 7.456583 | 0.011585 | Adaptive immune response and immune response. |
| *LAG3* | Lymphocyte activating 3 | 7.452066 | 0.000109 | Adaptive immune response, plasmacytoid dendritic cell activation, cell surface receptor signaling pathway, negative regulation of interleukin-2 production, T cell activation, natural killer cell mediated cytotoxicity, innate immune response, negative regulation of regulatory T cell differentiation, positive regulation of natural killer cell mediated cytotoxicity, and regulation of immune response. |
| *IGHV5-51* | Immunoglobulin heavy variable 5-51 | 7.28631 | 0.043656 | Immunoglobulin mediated immune response. |
| *MMP1* | matrix metallopeptidase 1 | 7.200133 | 0.003031 | Proteolysis, extracellular matrix disassembly, extracellular matrix organization, collagen catabolic process, positive regulation of protein-containing complex assembly, and cellular response to UV-A. |
| *SLC12A3* | Solute carrier family 12 member 3 | 7.145034 | 0.003464 | Response to dietary excess, monoatomic ion transport, sodium ion transport, cell volume homeostasis, signal transduction, sodium ion transmembrane transport, chloride ion homeostasis, potassium ion homeostasis, sodium ion homeostasis, renal sodium ion absorption, chloride transmembrane transport, response to aldosterone, and potassium ion import across plasma membrane. |
| *TRAJ27* | T cell receptor alpha joining 27 | 7.109609 | 0.002165 | Unknown function. |
| *GZMB* | Granzyme B | 7.054876 | 0.017302 | Proteolysis, apoptotic process, negative regulation of translation, killing of cells of another organism, natural killer cell mediated cytotoxicity, proteolysis involved in protein catabolic process, protein maturation, pyroptotic inflammatory response, granzyme-mediated programmed cell death signaling pathway, and positive regulation of establishment of protein localization to mitochondrion. |
| *CD27* | CD27 molecule | 6.998513 | 0.006359 | Cell surface receptor signaling pathway, immunoglobulin mediated immune response, T cell activation, negative regulation of apoptotic process, response to ethanol, positive regulation of B cell differentiation, positive regulation of T cell differentiation, positive regulation of JNK cascade, negative regulation of T cell apoptotic process, adaptive immune memory response involving T cells and B cells, extrinsic apoptotic signaling pathway, CD27 signaling pathway, and positive regulation of non-canonical NF-kappaB signal transduction. |
| *EOMES* | Eomesodermin | 6.948721 | 0.018417 | Negative regulation of transcription by RNA polymerase II, endoderm formation, mesoderm formation, endodermal cell fate specification, neuron migration, trophectodermal cell differentiation, adaptive immune response, CD8-positive, alpha-beta T cell differentiation involved in immune response, chromatin remodeling, regulation of transcription by |

| | | | | |
|---|---|---|---|---|
| | | | | RNA polymerase II, brain development, cardioblast differentiation, gene expression, stem cell population maintenance, olfactory bulb development, cerebral cortex regionalization, cerebral cortex neuron differentiation, skeletal muscle cell differentiation, negative regulation of DNA-binding transcription factor activity, positive regulation of DNA-templated transcription, positive regulation of transcription by RNA polymerase II, mesendoderm development, astrocyte differentiation, cell differentiation involved in embryonic placenta development, and mesodermal to mesenchymal transition involved in gastrulation. |
| *TIGIT* | T cell immunoreceptor with Ig and ITIM domains | 6.844114 | 0.005762 | Negative regulation of interleukin-12 production, positive regulation of interleukin-10 production, negative regulation of natural killer cell mediated cytotoxicity, and negative regulation of T cell activation. |
| *TRGV8* | T cell receptor gamma variable 8 | 6.836902 | 0.001194 | Adaptive immune response and innate immune response. |
| *CCL4L2* | C-C motif chemokine ligand 4 like 2 | 6.798009 | 0.016713 | Immune response. |
| *GBP5* | Guanylate binding protein 5 | 6.761607 | 0.024128 | Activation of innate immune response, protein targeting, inflammatory response, positive regulation of interleukin-1 beta production, positive regulation of interleukin-18 production, protein localization to Golgi apparatus, defense response to bacterium, innate immune response, positive regulation of innate immune response, protein homotetramerization, defense response to virus, cytolysis in another organism, cellular response to lipopolysaccharide, cellular response to type II interferon, positive regulation of pyroptotic inflammatory response, positive regulation of AIM2 inflammasome complex assembly, positive regulation of cytokine production involved in inflammatory response, and positive regulation of NLRP3 inflammasome complex assembly. |
| *IGHV3-20* | Immunoglobulin heavy variable 3-20 | 6.748827 | 0.015275 | Adaptive immune response and immunoglobulin mediated immune response. |
| *IGHV4-39* | Immunoglobulin heavy variable 4-39 | 6.733877 | 0.038687 | Adaptive immune response, immune response and immunoglobulin mediated immune response. |
| *IGHJ5* | Immunoglobulin heavy joining 5 | 6.728236 | 0.017197 | Unknown function. |
| *FCGR1CP* | Fc gamma receptor Ic, pseudogene | 6.699464 | 0.008158 | Unknown function. |
| *GZMH* | Granzyme H | 6.653307 | 0.014293 | Proteolysis, apoptotic process, killing of cells of another organism, and protein maturation. |
| *UBASH3A* | Ubiquitin associated and SH3 domain containing A | 6.646908 | 0.018167 | Regulation of cytokine production and negative regulation of T cell receptor signaling pathway. |
| *HSF5* | Heat shock transcription factor 5 | 6.604906 | 0.005927 | Regulation of transcription by RNA polymerase II, male meiotic nuclear division, spermatogenesis, cell differentiation, negative regulation of DNA-templated transcription, and positive regulation of DNA-templated transcription. |
| *TRAJ6* | T cell receptor alpha joining 6 | 6.571817 | 0.014883 | Unknown function. |
| *TRGV10* | T cell receptor gamma variable 10 (non-functional) | 6.557693 | 0.003105 | Adaptive immune response. |
| *NKG7* | Natural killer cell granule protein 7 | 6.554512 | 0.005317 | Natural killer cell mediated cytotoxicity directed against tumor cell target, inflammatory response, killing of cells of another organism, CD4-positive, alpha-beta T cell activation, natural killer cell mediated cytotoxicity, defense response to protozoan, natural killer cell degranulation, positive regulation of inflammatory response, and granzyme-mediated programmed cell death signaling pathway. |
| *SIRPG* | Signal regulatory protein gamma | 6.537991 | 0.004085 | Cell adhesion, cell-cell signaling, positive regulation of cell population proliferation, negative regulation of cell population proliferation, positive regulation of cell-cell adhesion, intracellular signal transduction, positive regulation of phagocytosis, and positive regulation of T cell activation. |
| *KLRC2* | Killer cell lectin like receptor C2 | 6.535524 | 0.00298 | Stimulatory C-type lectin receptor signaling pathway, natural killer cell mediated immunity, adaptive immune response, cellular defense response, signal transduction, regulation of natural killer cell activation, positive regulation of natural killer cell degranulation, and positive regulation of natural killer cell mediated cytotoxicity. |
| *RTP5* | Receptor transporter protein 5 (putative) | 6.515603 | 0.001882 | Detection of chemical stimulus involved in sensory perception of bitter taste, protein targeting to membrane, and protein insertion into membrane. |

#### Supplementary Materials

| | | | | |
|---|---|---|---|---|
| *ANTXRLP1* | ANTXR like pseudogene 1 | 6.467276 | 0.001079 | Unknown function. |
| *IGHV1-3* | Immunoglobulin heavy variable 1-3 | 6.46436 | 0.035361 | Immunoglobulin mediated immune response. |
| *IL4I1* | Interleukin 4 induced 1 | 6.440281 | 0.002316 | Adaptive immune response, regulation of adaptive immune response, negative regulation of T cell mediated immune response to tumor cell, L-phenylalanine catabolic process, tryptophan catabolic process, tyrosine catabolic process, amino acid catabolic process, tryptophan catabolic process to indole-3-acetate, negative regulation of T cell proliferation, regulation of B cell differentiation, positive regulation of regulatory T cell differentiation, positive regulation of transcription by RNA polymerase II, and negative regulation of T cell activation. |
| *TNR* | Tenascin R | 6.402399 | 0.007672 | Cell adhesion, neuron cell-cell adhesion, nervous system development, axon guidance, associative learning, negative regulation of neuron projection development, telencephalon cell migration, negative regulation of cell-cell adhesion, regulation of cell adhesion, extracellular matrix organization, regulation of cell migration, synaptic transmission, glutamatergic, locomotory exploration behavior, regulation of cell differentiation, axon extension involved in regeneration, negative regulation of axon extension involved in regeneration, negative regulation of synaptic transmission, synapse organization, neuromuscular process controlling balance, positive regulation of synaptic transmission, glutamatergic, positive regulation of transmission of nerve impulse, long-term synaptic potentiation, and neuroblast migration. |
| *FCRL5* | Fc receptor like 5 | 6.386703 | 0.0316 | Immune response, cell surface receptor signaling pathway, single fertilization, and B cell activation. |
| *IGKV3D-15* | Immunoglobulin kappa variable 3D-15 | 6.375818 | 0.034592 | Adaptive immune response and immune response. |
| *NUGGC* | Nuclear GTPase, germinal center associated | 6.347175 | 0.016453 | Regulation of nuclear cell cycle DNA replication, negative regulation of apoptotic process, cellular response to lipopolysaccharide. |
| *TRAJ7* | T cell receptor alpha joining 7 | 6.326581 | 0.002526 | Unknown function. |
| *IL21R* | Interleukin 21 receptor | 6.325731 | 0.034394 | Cell surface receptor signaling pathway, immunoglobulin mediated immune response, cytokine-mediated signaling pathway, natural killer cell activation, and interleukin-21-mediated signaling pathway. |
| *SH2D1A* | SH2 domain containing 1A | 6.316964 | 0.006784 | Adaptive immune response, immune system process, humoral immune response, cellular defense response, cell-cell signaling, natural killer cell activation, natural killer cell mediated cytotoxicity, positive regulation of natural killer cell mediated cytotoxicity, regulation of immune response, and negative regulation of T cell receptor signaling pathway. |

*P-values were corrected by multiple hypothesis testing to give the Q-value by the Benjamini-Hochberg method. **Functional Annotation is adapted from Database for Annotation, Visualization and Integrated Discovery (DAVID).

#### Supplementary Materials

**Supplemental Table 15** Top 50 Downregulated Genes in Mixed LS *versus* HC.

| Gen | Protein Encoded | logFC | Q-Value* | Functional Annotation** |
|---|---|---|---|---|
| *TIMP4* | TIMP metallopeptidase inhibitor 4 | -6.95639 | 0.003031 | Notch signaling pathway, response to hormone, and response to cytokine, and negative regulation of membrane protein ectodomain proteolysis. |
| *MT1G* | Metallothionein 1G | -6.60475 | 0.000328 | Intracellular zinc ion homeostasis, detoxification of copper ion, monocyte differentiation, cellular response to vascular endothelial growth factor stimulus, monocyte activation, negative regulation of growth, cellular response to cadmium ion, cellular response to copper ion, and cellular response to zinc ion. |
| *CES1P1* | Carboxylesterase 1 pseudogene 1 | -6.32189 | 0.007154 | Anatomical structure morphogenesis and lipid catabolic process. |
| *ENSG00000 222007* | Novel transcript, antisense to AGAP1 | -6.21662 | 0.000895 | Pseudogene. |
| *TNXA* | Tenascin XA (pseudogene) | -5.89225 | 0.004886 | Extracellular exosome. |
| *CLDN8* | Claudin 8 | -5.88965 | 0.003898 | Cell adhesion, calcium-independent cell-cell adhesion via plasma membrane cell-adhesion molecules, and bicellular tight junction assembly, |
| *LY6K* | Lymphocyte antigen 6 family member K | -5.71552 | 0.002132 | Binding of sperm to zona pellucida. |
| *PCP4* | Purkinje cell protein 4 | -5.59271 | 0.021587 | Positive regulation of neuron differentiation, positive regulation of CAMKK-AMPK signaling cascade. |
| *DIRAS3* | DIRAS family GTPase 3 | -5.43167 | 0.015473 | Regulation of cyclin-dependent protein serine/threonine kinase activity, small GTPase-mediated signal transduction, and genomic imprinting. |
| *BMP3* | Bone morphogenetic protein 3 | -5.15291 | 0.006022 | Skeletal system development, osteoblast differentiation, signal transduction, cell-cell signaling, negative regulation of ossification, and cartilage development. |
| *GPRC5A* | G protein-coupled receptor class C group 5 member A | -5.05791 | 0.027464 | Signal transduction, negative regulation of epidermal growth factor-activated receptor activity, and G protein-coupled receptor signaling pathway. |
| *EDN3* | Endothelin 3 | -5.01118 | 0.034394 | Neural crest cell migration, positive regulation of leukocyte chemotaxis, regulation of systemic arterial blood pressure by endothelin, intracellular calcium ion homeostasis, signal transduction, cell surface receptor signaling pathway, cell-cell signaling, axon guidance, blood circulation, cell population proliferation, positive regulation of cell population proliferation, positive regulation of heart rate, regulation of gene expression, intracellular magnesium ion homeostasis, vein smooth muscle contraction, regulation of vasoconstriction, peptide hormone secretion, melanocyte differentiation, neutrophil chemotaxis, vasoconstriction, positive regulation of MAP kinase activity, positive regulation of cell differentiation, positive regulation of mitotic nuclear division, positive regulation of smooth muscle contraction, positive regulation of hormone secretion, regulation of developmental pigmentation, axon extension, establishment of localization in cell, potassium ion transmembrane transport, and positive regulation of potassium ion transmembrane transport. |
| *MIR28* | MicroRNA 28 | -5.00975 | 0.012968 | Positive regulation of smooth muscle cell apoptotic process, miRNA-mediated post-transcriptional gene silencing, miRNA-mediated gene silencing by inhibition of translation, and negative regulation of interleukin-34 production. |
| *SLPI* | Secretory leukocyte peptidase inhibitor | -4.994 | 0.013187 | Immune response, antibacterial humoral response, negative regulation of protein binding, response to lipopolysaccharide, modulation of process of another organism, negative regulation of viral genome replication, and innate immune response. |
| *GSTT2B* | Glutathione S-transferase theta 2B | -4.91136 | 0.049988 | Glutathione metabolic process. |
| *SLC7A14* | Solute carrier family 7 member 14 | -4.90818 | 0.022105 | Amino acid transmembrane transport, amino acid transport, gamma-aminobutyric acid import. |
| *TECRP2* | Trans-2,3-enoyl-CoA reductase pseudogene 2 | -4.82389 | 0.00234 | Unknown function. |
| *GREM2* | Gremlin 2, DAN family BMP antagonist | -4.81708 | 0.006249 | Animal organ morphogenesis, embryonic body morphogenesis, cytokine-mediated signaling pathway, sequestering of BMP from receptor via BMP binding, determination of dorsal identity, regulation of cytokine activity. |

#### Supplementary Materials

| | | | | |
|---|---|---|---|---|
| *HOXA5* | Homeobox A5 | -4.78609 | 0.033276 | Respiratory system process, regulation of transcription by RNA polymerase II, anterior/posterior pattern specification, positive regulation of gene expression, cell migration, negative regulation of angiogenesis, thyroid gland development, regulation of mammary gland epithelial cell proliferation, multicellular organism growth, positive regulation of apoptotic process, positive regulation of myeloid cell differentiation, negative regulation of erythrocyte differentiation, positive regulation of transcription by RNA polymerase II, lung alveolus development, embryonic skeletal system morphogenesis, bronchiole development, epithelial tube branching involved in lung morphogenesis, lung goblet cell differentiation, lung-associated mesenchyme development, trachea cartilage morphogenesis, intestinal epithelial cell maturation, mesenchymal-epithelial cell signaling, mammary gland epithelial cell differentiation, mammary gland alveolus development, and cell-cell signaling involved in mammary gland development. |
| *NTF4* | Neurotrophin 4 | -4.73619 | 0.014293 | Cell surface receptor protein tyrosine kinase signaling pathway, ganglion mother cell fate determination, long-term memory, sensory organ boundary specification, adult locomotory behavior, epidermis development, nerve development, positive regulation of peptidyl-serine phosphorylation, ameloblast differentiation, nerve growth factor signaling pathway, mechanoreceptor differentiation, negative regulation of neuron apoptotic process, neuron projection morphogenesis, modulation of chemical synaptic transmission, innervation, and taste bud development. |
| *ENSG00000 283541* | Contactin associated protein-like 3B (CNTNAP3B) pseudogene | -4.73065 | 0.032451 | *Paseudogene*. |
| *LEXM* | Lymphocyte expansion molecule | -4.72219 | 0.026757 | Unknown function. |
| *SNORD53* | Small nucleolar RNA, C/D box 53 | -4.69753 | 0.043274 | RNA processing. |
| *MIR99A* | MicroRNA 99a | -4.68817 | 0.040802 | miRNA-mediated post-transcriptional gene silencing, negative regulation of interleukin-6-mediated signaling pathway, negative regulation of receptor signaling pathway via STAT. |
| *NPTX2* | Neuronal pentraxin 2 | -4.63672 | 0.020102 | Chemical synaptic transmission, and associative learning. |
| *SLX1A-SULT 1A3* | SLX1A-SULT1A3 readthrough (NMD candidate) | -4.61035 | 0.041324 | Unknown function. |
| *ATP5MC1P6* | ATP synthase membrane subunit c locus 1 pseudogene 6 | -4.60084 | 0.007987 | Unknown function. |
| *ODF3L1* | Outer dense fiber of sperm tails 3 like 1 | -4.59943 | 0.026918 | Unknown function. |
| *FAM181B* | Family with sequence similarity 181 member B | -4.5888 | 0.006509 | Unknown function. |
| *FGFBP2* | Fibroblast growth factor binding protein 2 | -4.54733 | 0.049195 | Cell-cell signaling. |
| *PKIA* | cAMP-dependent protein kinase inhibitor alpha | -4.53734 | 0.012985 | Negative regulation of transcription by RNA polymerase II, regulation of G2/M transition of mitotic cell cycle, negative regulation of protein import into nucleus, negative regulation of cAMP/PKA signal transduction, and negative regulation of cAMP-dependent protein kinase activity. |
| *CES1* | Carboxylesterase 1 | -4.52621 | 0.018183 | Anatomical structure morphogenesis, and lipid catabolic process. |
| *CYP2W1* | Cytochrome P450 family 2 subfamily W member 1 | -4.51443 | 0.01531 | Organic acid metabolic process, phospholipid metabolic process, xenobiotic metabolic process, retinoic acid catabolic process, aflatoxin metabolic process, and cytochrome metabolic process. |
| *SERTM1* | Serine rich and transmembrane domain containing 1 | -4.51383 | 0.017581 | Membrane and intracellular membrane-bounded organelle. |
| *AKR7A3* | Aldo-keto reductase family 7 member A3 | -4.51087 | 0.011309 | Aldehyde metabolic process. |
| *ENSG00000 286430* | Novel transcript, antisense to PEMT | -4.47929 | 0.016453 | *Pseudogene*. |
| *MDFI* | MyoD family inhibitor | -4.43187 | 0.013286 | Negative regulation of transcription by RNA polymerase II, dorsal/ventral axis specification, regulation of Wnt signaling pathway, negative regulation of Wnt signaling pathway, cytoplasmic sequestering of transcription |

| | | | | |
|---|---|---|---|---|
| | | | | factor, negative regulation of DNA binding, negative regulation of DNA-templated transcription, regulation of JNK cascade, embryonic skeletal system morphogenesis, and trophoblast giant cell differentiation. |
| *RPL7P60* | Ribosomal protein L7 pseudogene 60 | -4.29405 | 0.015102 | Unknown function. |
| *PNMA8B* | PNMA family member 8B | -4.26722 | 0.003518 | Unknown function. |
| *ENSG00000 286488* | Novel transcript, antisense to STARD5 | -4.26415 | 0.012915 | *Pseudogene*. |
| *CLEC3B* | C-type lectin domain family 3 member B | -4.25133 | 0.017903 | Ossification, positive regulation of plasminogen activation, and bone mineralization. |
| *LYVE1* | Lymphatic vessel endothelial hyaluronan receptor 1 | -4.22923 | 0.028246 | Positive regulation of cellular extravasation, receptor-mediated endocytosis, cell adhesion, cell-matrix adhesion, response to wounding, anatomical structure morphogenesis, and hyaluronan catabolic process. |
| *KLF15* | KLF transcription factor 15 | -4.21315 | 0.02298 | Intracellular glucose homeostasis, regulation of transcription by RNA polymerase II, glial cell differentiation, cardiac muscle hypertrophy in response to stress, regulation of Wnt signaling pathway, response to insulin, positive regulation of transcription by RNA polymerase II, positive regulation of D-glucose import, podocyte differentiation, cellular response to peptide, and negative regulation of peptidyl-lysine acetylation. |
| *DUSP1* | Dual specificity phosphatase 1 | -4.20656 | 0.045239 | Endoderm formation, chromatin remodeling, regulation of transcription by RNA polymerase II, negative regulation of cell adhesion, signal transduction, negative regulation of cell population proliferation, peptidyl-tyrosine dephosphorylation, peptidyl-threonine dephosphorylation, negative regulation of MAP kinase activity, negative regulation of MAPK cascade, negative regulation of meiotic cell cycle, peptidyl-serine dephosphorylation, negative regulation of monocyte chemotaxis, regulation of mitotic cell cycle spindle assembly checkpoint, negative regulation of p38MAPK cascade, and cellular response to chemokine. |
| *ENSG00000 276663* | Novel transcript, antisense to ARL2BP | -4.16197 | 0.029347 | *Pseudogene*. |
| *SLITRK6* | SLIT and NTRK like family member 6 | -4.14 | 0.013903 | Startle response, lens development in camera-type eye, auditory receptor cell morphogenesis, axonogenesis, synapse assembly, visual perception, sensory perception of sound, adult locomotory behavior, vestibulocochlear nerve development, auditory behavior, multicellular organism growth, positive regulation of synapse assembly, linear vestibuloocular reflex, innervation, cochlea development, and regulation of presynapse assembly. |
| *DSCAS* | DSC1/DSC2 antisense RNA | -4.11322 | 0.029814 | Unknown function. |
| *SLC25A4* | Solute carrier family 25 member 4 | -4.11215 | 0.0019 | Mitochondrial genome maintenance, generation of precursor metabolites and energy, apoptotic mitochondrial changes, adenine transport, ADP transport, regulation of mitochondrial membrane permeability, negative regulation of necroptotic process, mitochondrial ADP transmembrane transport, negative regulation of mitochondrial outer membrane permeabilization involved in apoptotic signaling pathway, positive regulation of mitophagy, proton transmembrane transport, mitochondrial ATP transmembrane transport, and adaptive thermogenesis. |
| *CYP51A1* | Cytochrome P450 family 51 subfamily A member 1 | -4.1106 | 0.011785 | Lipid metabolic process, steroid biosynthetic process, cholesterol biosynthetic process, cholesterol metabolic process, sterol metabolic process, cholesterol biosynthetic process via 24,25-dihydrolanosterol, negative regulation of protein catabolic process, small molecule biosynthetic process, negative regulation of protein secretion, negative regulation of amyloid-beta clearance, organic hydroxy compound biosynthetic process. |
| *RAB40A* | RAB40A, member RAS oncogene family | -4.07631 | 0.041032 | Exocytosis, protein secretion, protein ubiquitination, and intracellular signal transduction. |

*P-values were corrected by multiple hypothesis testing to give the Q-value by the Benjamini-Hochberg method. **Functional Annotation is adapted from Database for Annotation, Visualization and Integrated Discovery (DAVID).

#### Supplementary Materials

**Supplemental Table 16.** Top 50 Upregulated Genes in Inactive LS *versus* HC.

| Gen | Protein Encoded | logFC | Q-Value* | Functional Annotation** |
|---|---|---|---|---|
| *PCDH10* | Protocadherin 10 | 6.345631 | 0.035905 | Cell adhesion, homophilic cell adhesion via plasma membrane adhesion molecules, nervous system development. |
| *ENSG00000 255532* | NADPH oxidase 4 pseudogene | 6.199091 | 0.010551 | *Pseudogene.* |
| *ASB4* | Ankyrin repeat and SOCS box containing 4 | 6.127844 | 0.035905 | Intracellular signal transduction, protein autoubiquitination, positive regulation of vasculogenesis. |
| *SMLR1* | Small leucine rich protein 1 | 6.051307 | 0.049745 | Membrane. |
| *TLE6* | TLE family member 6, subcortical maternal complex member | 5.968794 | 0.023459 | Negative regulation of transcription by RNA polymerase II, regulation of DNA-templated transcription, actin filament organization, positive regulation of embryonic development, establishment of spindle localization, regulation of cell division, endoplasmic reticulum localization, mitochondrion localization, embryonic process involved in female pregnancy, negative regulation of canonical Wnt signaling pathway. |
| *FOXE1* | Forkhead box E1 | 5.932883 | 0.023459 | Negative regulation of transcription by RNA polymerase II, regulation of transcription by RNA polymerase II, thyroid hormone generation, anatomical structure morphogenesis, cell migration, cell differentiation, thyroid gland development, hair follicle morphogenesis, negative regulation of DNA-templated transcription, positive regulation of DNA-templated transcription, thymus development, embryonic organ morphogenesis, hard palate development, soft palate development, chordate pharynx development, and cranial skeletal system development. |
| *OR1Q1* | Olfactory receptor family 1 subfamily Q member 1 | 5.879129 | 0.035905 | Signal transduction, G protein-coupled receptor signaling pathway, sensory perception of smell, and detection of chemical stimulus involved in sensory perception of smell. |
| *STEAP1B* | STEAP family member 1B | 5.493165 | 0.035905 | Endosome, plasma membrane and membrane. |
| *KIF19* | Kinesin family member 19 | 5.419611 | 0.023459 | Microtubule-based movement, axonemal microtubule depolymerization and plus-end specific microtubule depolymerization. |
| *OR3A4P* | Olfactory receptor family 3 subfamily A member 4 pseudogene | 5.33793 | 0.035905 | Signal transduction and sensory perception of smell. |
| *VWCE* | Von Willebrand factor C and EGF domains | 5.274359 | 0.023459 | Cellular response to virus. |
| *OR1E2* | Olfactory receptor family 1 subfamily E member 2 | 4.976998 | 0.04064 | Signal transduction, G protein-coupled receptor signaling pathway, sensory perception of chemical stimulus, detection of chemical stimulus involved in sensory perception of smell. |
| *OR56A5* | Olfactory receptor family 56 subfamily A member 5 | 4.808939 | 0.035905 | G protein-coupled receptor signaling pathway, detection of chemical stimulus involved in sensory perception of smell. |
| *RSAD2* | Radical S-adenosyl methionine domain containing 2 | 4.691546 | 0.049745 | Response to virus, positive regulation of toll-like receptor 7 signaling pathway, positive regulation of toll-like receptor 9 signaling pathway, CD4-positive, alpha-beta T cell activation, CD4-positive, alpha-beta T cell differentiation, negative regulation of viral genome replication, innate immune response, negative regulation of protein secretion, positive regulation of immune response, defense response to virus, positive regulation of T-helper 2 cell cytokine production. |
| *ENSG00000 231305* | IFT122 pseudogene 3 | 4.674466 | 0.023459 | *Pseudogene.* |
| *TBX2* | T-box transcription factor 2 | 2.695704 | 0.049745 | Negative regulation of transcription by RNA polymerase II, cell fate specification, heart looping, outflow tract septum morphogenesis, outflow tract morphogenesis, endocardial cushion morphogenesis, endocardial cushion formation, regulation of transcription by RNA polymerase II, apoptotic process, Notch signaling pathway, muscle cell fate determination, regulation of heart contraction, fibroblast growth factor receptor signaling pathway, neurogenesis, response to retinoic acid, embryonic heart tube development, aorta |

#### Supplementary Materials

| | | | | |
|---|---|---|---|---|
| | | | | Morphogenesis, atrioventricular canal development, embryonic digit morphogenesis, pigment metabolic process involved in pigmentation, negative regulation of DNA-templated transcription, positive regulation of DNA-templated transcription, positive regulation of transcription by RNA polymerase II, embryonic camera-type eye morphogenesis, cardiac muscle tissue development, smooth muscle cell differentiation, roof of mouth development, pharyngeal system development, positive regulation of cardiac muscle cell proliferation, cardiac muscle cell myoblast differentiation, epithelial tube branching involved in lung morphogenesis, developmental growth involved in morphogenesis, mammary placode formation, mesenchymal cell proliferation involved in lung development, cellular response to dexamethasone stimulus, ureteric peristalsis, cochlea morphogenesis, cellular senescence, melanocyte proliferation, negative regulation of heart looping, negative regulation of cardiac chamber formation, positive regulation of cell cycle G1/S phase transition, cardiac jelly development, atrioventricular canal morphogenesis, and negative regulation of cellular senescence. |
| *RUNX1* | RUNX family transcription factor 1 | 1.712447 | 0.036882 | Negative regulation of transcription by RNA polymerase II, ossification, chondrocyte differentiation, myeloid leukocyte differentiation, regulation of transcription by RNA polymerase II, regulation of plasminogen activation, hemopoiesis, myeloid cell differentiation, neuron differentiation, negative regulation of granulocyte differentiation, positive regulation of granulocyte differentiation, positive regulation of interleukin-2 production, positive regulation of collagen biosynthetic process, negative regulation of CD4-positive, alpha-beta T cell differentiation, positive regulation of CD8-positive, alpha-beta T cell differentiation, regulation of cell differentiation, positive regulation of angiogenesis, positive regulation of DNA-templated transcription, positive regulation of transcription by RNA polymerase II, peripheral nervous system neuron development, positive regulation of developmental process, regulation of cardiac muscle cell proliferation, cardiac muscle tissue regeneration, hematopoietic stem cell proliferation, positive regulation of extracellular matrix organization, and regulation of connective tissue replacement. |

*P-values were corrected by multiple hypothesis testing to give the Q-value by the Benjamini-Hochberg method. **Functional Annotation is adapted from Database for Annotation, Visualization and Integrated Discovery (DAVID).

#### Supplementary Materials

**Supplemental Table 17.** Top 50 Downregulated Genes in Inactive LS *versus* HC.

| Gen | Protein Encoded | logFC | Q-Value* | Functional Annotation** |
|---|---|---|---|---|
| *LY6K* | Lymphocyte antigen 6 family member K | -6.365 68 | 0.023459 | Binding of sperm to zona pellucida. |
| *RAB6C* | RAB6C, member RAS oncogene family | -5.094 02 | 0.037545 | Mitotic cell cycle, intracellular protein transport, retrograde vesicle-mediated transport, Golgi to endoplasmic reticulum, intra-Golgi vesicle-mediated transport, small GTPase-mediated signal transduction, response to xenobiotic stimulus, regulation of centrosome duplication, vesicle-mediated transport, retrograde transport, endosome to Golgi, protein localization to Golgi membrane. |
| *ENSG00000 283390* | PET100 pseudogene 1 | -5.075 57 | 0.023459 | *Pseudogene*. |
| *HEY1* | Hes related family bHLH transcription factor with YRPW motif 1 | -4.962 54 | 0.035905 | Negative regulation of transcription by RNA polymerase II, angiogenesis, cardiac conduction system development, aortic valve morphogenesis, pulmonary valve morphogenesis, atrioventricular valve formation, endocardial cushion morphogenesis, cardiac ventricle morphogenesis, regulation of DNA-templated transcription, regulation of transcription by RNA polymerase II, Notch signaling pathway, anterior/posterior pattern specification, dorsal aorta morphogenesis, umbilical cord morphogenesis, negative regulation of neuron differentiation, negative regulation of Notch signaling pathway, negative regulation of DNA-templated transcription, positive regulation of transcription by RNA polymerase II, regulation of neurogenesis, negative regulation of smooth muscle cell differentiation, cardiac epithelial to mesenchymal transition, heart trabecula formation, cardiac septum morphogenesis, ventricular septum morphogenesis, labyrinthine layer blood vessel development, arterial endothelial cell differentiation, negative regulation of biomineral tissue development, circulatory system development, regulation of vasculogenesis. |
| *C17orf58* | Chromosome 17 open reading frame 58 | -4.802 16 | 0.049745 | Collagen-containing extracellular matrix. |
| *CD55* | CD55 molecule (Cromer blood group) | -2.169 97 | 0.049745 | Positive regulation of T cell cytokine production, complement activation, classical pathway, positive regulation of cytosolic calcium ion concentration, regulation of complement activation, regulation of lipopolysaccharide-mediated signaling pathway, innate immune response, respiratory burst, negative regulation of complement activation, symbiont entry into host cell, regulation of complement-dependent cytotoxicity, positive regulation of CD4-positive, alpha-beta T cell activation, positive regulation of CD4-positive, and alpha-beta T cell proliferation. |
| *ERRFI1* | ERBB receptor feedback inhibitor 1 | -2.028 32 | 0.036882 | Liver development, tissue homeostasis, glucose metabolic process, bile acid biosynthetic process, apoptotic process, epidermal growth factor receptor signaling pathway, negative regulation of epidermal growth factor-activated receptor activity, embryo implantation, cholesterol metabolic process, response to xenobiotic stimulus, gene expression, cell migration, negative regulation of protein autophosphorylation, response to estradiol, response to progesterone, response to insulin, uterine epithelium development, chondrocyte proliferation, limb joint morphogenesis, negative regulation of epidermal growth factor receptor signaling pathway, cholesterol homeostasis, phosphatidylinositol 3-kinase/protein kinase B signal transduction, skin morphogenesis, regulation of keratinocyte differentiation, lung alveolus development, epithelial cell proliferation, progesterone receptor signaling pathway, cartilage development, lung vasculature development, lung epithelium development, fat pad development, protein localization to plasma membrane, and response to 1-oleoyl-sn-glycerol 3-phosphate. |
| *KLF6* | KLF transcription factor 6 | -1.747 76 | 0.015783 | Regulation of transcription by RNA polymerase II, B cell differentiation, positive regulation of DNA-templated transcription, positive regulation of transcription by RNA polymerase II. |
| *PTMA* | Prothymosin alpha | -1.214 05 | 0.00033 | DNA-templated transcription, negative regulation of apoptotic process, positive regulation of transcription by RNA polymerase II. |
| *SKP1* | S-phase kinase associated protein 1 | -1.077 44 | 0.023459 | Protein polyubiquitination, chromatin remodeling, ubiquitin-dependent protein catabolic process, protein monoubiquitination, protein ubiquitination, SCF-dependent proteasomal ubiquitin-dependent protein catabolic process, proteasome-mediated ubiquitin-dependent protein catabolic process, maintenance of protein location in nucleus, protein K48-linked ubiquitination, positive regulation of ubiquitin protein ligase activity. |

#### Supplementary Materials

*P-values were corrected by multiple hypothesis testing to give the Q-value by the Benjamini-Hochberg method. **Functional Annotation is adapted from Database for Annotation, Visualization and Integrated Discovery (DAVID).

**Supplemental Table 18.** Senescence gene set.

| |
|---|
| ATM;CBX2;CBX8;CCL2;CCNB1;CCNE1;CCNE2;CDK2;CDK4;CDK6;CDKN1A;CDKN1B;CDKN2A;CDKN2B;CDKN2C;CDKN2D;CXCL1;CXCL10;CXCL2;CXCL8;E2F1;E2F2;E2F3;GDF15;GDNF;H2AX;HMGA1;HMGA2;HMGB1;ICAM1;ID1;IGFBP2;IGFBP3;IGFBP4;IGFBP5;IGFBP6;IGFBP7;IL15;IL1A;IL6;LAMB1;LIF;LMNB1;MDM2;MDM4;MMP1;MMP10;MMP12;MMP13;MMP14;MMP2;MMP3;MMP9;PAPPA;SERPINE1;SERPINE2;TERF1;TERF2;TERF2IP;TFDP1;TFDP2;TGFB2;TIMP1;TIMP2;TNF;TP53;VCAM1;VEGFA |

**Supplemental Table 19.** Cell type proportion comparisons across disease state and sub-classifications.

Table is present as a separate csv file.

**Supplemental Table 20.** Cell type-specific DEGs for LS vs. HC.

Table is present as a separate csv file.
